# Heterologous SARS-CoV-2 Booster Vaccinations – Preliminary Report

**DOI:** 10.1101/2021.10.10.21264827

**Authors:** Robert L. Atmar, Kirsten E. Lyke, Meagan E. Deming, Lisa A. Jackson, Angela R. Branche, Hana M. El Sahly, Christina A. Rostad, Judith M. Martin, Christine Johnston, Richard E. Rupp, Mark J. Mulligan, Rebecca C. Brady, Robert W. Frenck, Martín Bäcker, Angelica C. Kottkamp, Tara M. Babu, Kumaravel Rajakumar, Srilatha Edupuganti, David Dobryzynski, Christine M. Posavad, Janet I. Archer, Sonja Crandon, Seema U. Nayak, Daniel Szydlo, Jillian Zemanek, Clara P. Dominguez Islas, Elizabeth R. Brown, Mehul S. Suthar, M. Juliana McElrath, Adrian B. McDermott, Sarah E. O’Connell, David C. Montefiori, Amanda Eaton, Kathleen M. Neuzil, David S. Stephens, Paul C. Roberts, John H. Beigel, the DMID 21-0012 Study Group

## Abstract

**Background:** While Coronavirus disease 2019 (Covid-19) vaccines are highly effective, breakthrough infections are occurring. Booster vaccinations have recently received emergency use authorization (EUA) for certain populations but are restricted to homologous mRNA vaccines. We evaluated homologous and heterologous booster vaccination in persons who had received an EUA Covid-19 vaccine regimen.

**Methods:** In this phase 1/2 open-label clinical trial conducted at ten U.S. sites, adults who received one of three EUA Covid-19 vaccines at least 12 weeks prior to enrollment and had no reported history of SARS-CoV-2 infection received a booster injection with one of three vaccines (Moderna mRNA-1273 100-μg, Janssen Ad26.COV2.S 5×10^10^ virus particles, or Pfizer-BioNTech BNT162b2 30-μg; nine combinations). The primary outcomes were safety, reactogenicity, and humoral immunogenicity on study days 15 and 29.

**Results:** 458 individuals were enrolled: 154 received mRNA-1273, 150 received Ad26.CoV2.S, and 153 received BNT162b2 booster vaccines. Reactogenicity was similar to that reported for the primary series. Injection site pain, malaise, headache, and myalgia occurred in more than half the participants. Booster vaccines increased the neutralizing activity against a D614G pseudovirus (4.2-76-fold) and binding antibody titers (4.6-56-fold) for all combinations; homologous boost increased neutralizing antibody titers 4.2-20-fold whereas heterologous boost increased titers 6.2-76-fold. Day 15 neutralizing and binding antibody titers varied by 28.7-fold and 20.9-fold, respectively, across the nine prime-boost combinations.

**Conclusion:** Homologous and heterologous booster vaccinations were well-tolerated and immunogenic in adults who completed a primary Covid-19 vaccine regimen at least 12 weeks earlier.

(Funded by National Institute of Allergy and Infectious Diseases; Clinical Trials.gov number, NCT04889209)

## INTRODUCTION

The successful development of safe and effective vaccines targeting the Severe Acute Respiratory Syndrome Coronavirus 2 (SARS-CoV-2) is a remarkable achievement. High vaccine efficacy against clinical outcomes, tolerability and safety resulted in Emergency Use Authorization (EUA) by the U.S. Food and Drug Administration (FDA) of three Covid-19 vaccines between December 2020 and February 2021.^1-3^ Widespread roll-out of these vaccines, as well as vaccines from other manufacturers worldwide, has resulted in more than 6.4 billion doses of vaccine administered, with 187.2 million U.S. individuals (65.3% of the population) being fully vaccinated as of October 8, 2021.^4^

Although the vaccines currently approved or available under EUA in the U.S. provide high levels of protection against severe illness and death, the increased transmission of the Delta variant resulted in increasing numbers of breakthrough infections in fully vaccinated individuals.^5-7^ This coincided with evidence of waning of immunity in some vaccinated populations.^6-8^ Booster vaccines may enhance waning immunity and expand the breadth of immunity against SARS-CoV-2 variants of concern. Levels of binding and neutralizing antibodies correlate with vaccine efficacy for both mRNA and adenovirus-vectored vaccines, and likely have utility in predicting efficacy after boosting.^9-12^ A homologous booster with 30μg BNT162b2 increased neutralizing antibody titers against wild-type virus and the Delta variant to more than 5 times as high as after dose 2.^13^ A booster injection of 50 µg of mRNA-1273 increased neutralizing antibody titers against wild-type virus and the Delta variant 3.8-fold and 2.1-fold higher, respectively, than after the primary series.^14^ Based on evidence of increased neutralizing titers after a third dose of vaccine, homologous BNT162b2 booster vaccination received EUA and is currently recommended for high-risk populations and select EUA applications for homologous boosting with mRNA-1273 and Ad26.CoV2.S are under review by FDA.

Heterologous prime boost strategies may offer immunological advantages to optimize the breadth and longevity of protection achieved with currently available vaccines. Administration of an mRNA vaccine after the initial ChAdOx1 dose as the second dose of a two-dose regimen was safe and had enhanced immunogenicity compared to the two-dose homologous ChAdOx1 vaccine.^15-17^ The use of heterologous booster vaccines, if similarly tolerable and immunogenic, could also simplify the logistics of administering booster vaccines.

We conducted a phase 1/2 clinical trial to assess the safety, reactogenicity, and immunogenicity of booster vaccination in persons who previously received a Covid-19 vaccine under EUA at least 12 weeks earlier to rapidly generate data about the safety and immunogenicity of booster doses, including heterologous boosting regimens. We report the initial results of this trial.

## METHODS

### Trial Design and Participants

This phase 1/2 adaptive design, open-label clinical trial was performed in sequential stages at ten clinical sites. Eligible participants were healthy adults who had received a Covid-19 vaccine available under EUA (Ad26.COV2.S [Janssen], mRNA-1273 [Moderna, Inc], or BNT162b2 [Pfizer/BioNTech]) at least 12 weeks earlier and reported no prior history of SARS-CoV-2 infection or monoclonal antibody infusion. We did not screen for past or current evidence of SARS-CoV-2 infection to facilitate rapid enrollment into the study. Full eligibility criteria are available in the protocol (available at).

The trial was reviewed and approved by a central institutional review board and overseen by an independent safety monitoring committee. All participants provided written informed consent prior to undergoing any study-related activities.

### Vaccine

The study vaccines used were mRNA-1273 100-μg (Stage 1), Ad26.COV2.S 5×10^10^ virus particles (Stage 2), and BNT162b2 30-μg (Stage 3),^1-3^ providing 9 different combinations of primary vaccination-boost (Stage 1, groups 1-3; Stage 2, groups 4-6; Stage 3, groups 7-9). Vaccines were administered as directed in their respective EUAs.

### Trial Procedures

Study participants were enrolled in two age groups, 18-55 years and ≥ 56 years, with approximately equal numbers in the two age strata and the three EUA-dosed vaccine groups (N = 50/group). After informed consent was obtained, participants were screened by medical history, targeted physical examination, and urine pregnancy test (if indicated), and eligible persons received the study booster vaccine. Participants were observed for 30 minutes after vaccination. Blood was collected for immunogenicity assessments at Days 1 (pre-vaccination), 15, and 29.

Participants used a memory aid to record local and systemic solicited adverse events (AEs) for 7 days following vaccination. Unsolicited AEs through 28 days following vaccination also were collected. Grading of AEs used the FDA’s Toxicity Grading Scale for Healthy Adult and Adolescent Volunteers Enrolled in Preventive Vaccine Clinical Trials.^18^ All serious adverse events (SAEs), new onset chronic medical conditions (NOCMCs), adverse events of special interest (AESIs), or related medically attended adverse events (MAAEs) are collected for the duration of the 12-month study and are reported through Study Day 29 in this report.

### Immunogenicity

Immunogenicity was assessed on Day 1 prior to booster vaccination and at Days 15 and 29 (unavailable for all groups) after boost. Serum binding antibody (bAb) levels against S-2P were evaluated with the MSD® 384-well Custom Serology Assay Electrochemiluminescence Immunoassay (4-plex ECLIA) version 2 (in validation) and using the 10-plex ECLIA for emerging SARS-CoV-2 variant spike proteins.^19^ SARS-CoV-2 neutralization titers expressed as the serum inhibitory dilution required to achieve 50% and 80% neutralization (ID50 and ID80, respectively) were determined by group, age group and timepoint, using pseudotyped lentiviruses presenting SARS-CoV-2 spike variants D614G, and for groups 1 to 3, B.1.617.2 (Delta) and B.1.351 (Beta), as described previously.^20^ For the Beta variant, only a random subset of samples (20 per EUA vaccine, distributed equally between age groups and sites) was analyzed. Neutralizing activity was expressed in International Units (IU50/mL and IU80/mL) for the D614G neutralization assay.

### Statistical Analysis

Summaries of baseline characteristics (including serological endpoints) are reported for all enrolled participants. Safety and (follow-up) immunogenicity analyses were restricted to participants who received the booster vaccines. No statistical comparisons between groups were planned and the analyses of safety and immunogenicity endpoints are only descriptive. The selected sample sizes of 50 per group and 25 per age stratum, allow for 99.5% and 92.8% probability of observing at least one an AE with a true event rate of 10%, respectively. Confidence intervals were not adjusted for multiplicity. The statistical analysis plan for this study is available.

## RESULTS

### Trial population

Four hundred fifty-eight participants were enrolled: 154, 150 and 154 in each of the three stages (Fig. S1). One participant (Group 7) did not receive booster vaccination. The demographic characteristics were similar across stages (Table 1). The interval between priming EUA vaccination and booster vaccination was shortest among those who were boosted with mRNA-1273, reflecting the temporal progression of enrollment across the sequential study stages (Table 1). Two participants (one each in groups 4 and 6 of Stage 2) who had serologic evidence (nucleocapsid protein antibody) of prior SARS-CoV-2 infection and one participant (in group 5 of Stage 2) who developed Covid-19 two days prior to Study Day 29 were included in the analyses. No more than 6% of samples were unavailable for analysis per group.

**Table 1.** Characteristics of the Participants at Enrollment.

| Group | 1 | 2 | 3 | 4 | 5 | 6 | 7 | 8 | 9 |
| --- | --- | --- | --- | --- | --- | --- | --- | --- | --- |
| **Primary**<br>**EUA Immunization**<br>**Vaccine** | **Janssen** | **Moderna** | **Pfizer/BioNTech** | **Janssen** | **Moderna** | **Pfizer/BioNTech** | **Janssen** | **Moderna** | **Pfizer/BioNTech** |
|  | **Ad26.COV2-S** | **mRNA-1273** | **BNT162b2** | **Ad26.COV2-S** | **mRNA-1273** | **BNT162b2** | **Ad26.COV2-S** | **mRNA-1273** | **BNT162b2** |
|  | **5x10<sup>10</sup> vp** | **100-mcg** | **30-mcg** | **5x10<sup>10</sup> vp** | **100-mcg** | **30-mcg** | **5x10<sup>10</sup> vp** | **100-mcg** | **30-mcg** |
| **Booster** | **Moderna mRNA-1273 100-mcg** |  |  | **Janssen Ad26.COV2-S 5x10<sup>10</sup> vp** |  |  | **Pfizer/BioNTech BNT162b2 30-mcg** |  |  |
| **Total Number** | 53 | 51 | 50 | 50 | 49 | 51 | 53* | 51 | 50 |
| **Sex – no. (%)** |  |  |  |  |  |  |  |  |  |
| **Female** | 26 (49.1) | 32 (62.7) | 29 (58.0) | 27 (46.0) | 16 (32.7) | 23 (45.1) | 29 (54.7) | 26 (51.0) | 23 (46.0) |
| **Male** | 27 (50.9) | 19 (37.3) | 21 (42.0) | 23 (54.0) | 33 (67.3) | 28 (54.9) | 24 (45.3) | 25 (49.0) | 27 (54.0) |
| **Age – years** |  |  |  |  |  |  |  |  |  |
| **Mean (s.d.)** | 56.8 (14.5) | 53.1 (16.2) | 54.8 (17.4) | 50.1 (13.9) | 49.9 (16.8) | 50.3 (15.4) | 47.7 (14.5) | 54.3 (16.8) | 50.4 (17.9) |
| **Range** | 24-81 | 24-76 | 22-85 | 24-77 | 20-75 | 20-76 | 22-74 | 23-75 | 19-80 |
| **Race – no. (%)** |  |  |  |  |  |  |  |  |  |
| **Asian** | 4 (7.5) | 5 (9.8) | 4 (8.0) | 3 (6.0) | 5 (10.2) | 6 (11.8) | 1 (1.9) | 2 (3.9) | 1 (2.0) |
| **Hawaiian**<br>**or Pacific**<br>**Islander** | 0 (0.0) | 0 (0.0) | 0 (0.0) | 0 (0.0) | 0 (0.0) | 0 (0.0) | 1 (1.9) | 0 (0.0) | 0 (0.0) |
| **Black/African**<br>**American** | 1 (1.9) | 2 (3.9) | 3 (6.0) | 0 (0.0) | 0 (0.0) | 2 (3.9) | 0 (0.0) | 2 (3.9) | 1 (2.0) |
| **White** | 46 (86.8) | 41 (80.4) | 43 (86.0) | 44 (88.0) | 43 (87.8) | 40 (78.4) | 50 (94.3) | 47 (92.2) | 43 (86.0) |
| **Multi-racial** | 1 (1.9) | 3 (5.9) | 0 (0.0) | 3 (6.0) | 1 (2.0) | 2 (3.9) | 1 (1.9) | 0 (0.0) | 4 (8.0) |
| **Other** | 1 (1.9%) | 0 (0.0) | 0 (0.0) | 0 (0.0) | 0 (0.0) | 1 (2.0%) | 0 (0.0) | 0 (0.0) | 1 (2.0%) |
| **Ethnicity – no (%)** |  |  |  |  |  |  |  |  |  |
| **Non-Hispanic** | 49 (92.5) | 46 (90.2) | 47 (94.0) | 47 (94.0) | 49 (100.0) | 48 (94.1) | 51 (96.2) | 49 (96.1) | 45 (90.0) |
| **Hispanic/Latino** | 4 (7.5) | 4 (7.8) | 3 (6.0) | 2 (4.0) | 0 (0.0) | 3 (5.9) | 2 (3.8) | 2 (3.9) | 5 (10.0) |
| **Unknown/Not**<br>**reported** | 0 (0.0) | 1 (2.0) | 0 (0.0) | 1 (2.0) | 0 (0.0) | 0 (0.0) | 0 (0.0) | 0 (0.0) | 0 (0.0) |
| **Boost Interval**<br>**weeks** |  |  |  |  |  |  |  |  |  |
| **Mean (s.d.)** | 13.7 (1.0) | 16.4 (1.9) | 16.8 (2.2) | 17.7 (2.0) | 19.3 (4.2) | 20.6 (5.8) | 19.9 (2.5) | 22.9 (4.6) | 24.1 (5.2) |
| **Range** | 12.0-15.9 | 12.4-20.0 | 12.0-20.9 | 13.9-21.0 | 12.6-26.0 | 12.3-41.3 | 10.9-23.0 | 12.6-28.7 | 14.3-31.9 |

\* Includes one participant with early termination from the Study at Day 1 visit and who did not receive a booster vaccination.

### Vaccine Safety

Two SAEs, unrelated to study vaccination, were reported. One (acute renal failure due to rhabdomyolysis from a fall) was reported 30 days after mRNA-1273 vaccination, and the other (acute cholecystitis) occurred 24 days after Ad26.COV2.S vaccination. No pre-specified study-halting rules were met, and no new onset chronic medical conditions occurred through study day 29. One related AESI (severe vomiting that led to a medically attended visit the day after vaccination) occurred in group 5 (Ad26.COV2.S boost). The number (and percentage) of boosted participants reporting unsolicited AEs, of any severity grade, that were deemed related to the study product was 24/154 (15.6%) in mRNA-1273 recipients, 18/150 (12.0%) in Ad26.COV2.S recipients and 22/153 (14.4%) in BNT162b2 recipients Table S1-S3). Most participants reporting related AEs ranked the AE as Grade 2 severity at most. There were four related Grade 3 AEs: one participant in the mRNA-1273 group (vomiting) and three in the Ad26.COV2.S group (one each of vomiting, fatigue/abnormal feeling, insomnia).

Injection site AEs were common, with local pain or tenderness being reported in 75-86%, 71-84%, and 72-92% of mRNA-1273, Ad26COV2.S and BNT162b2 recipients, respectively (Fig. 1, Tables S4-S6). Most injection site reactions were graded as mild, and only two (one mRNA-1273 and one Ad26COV2.S) reported as severe. Malaise, myalgias, and headaches were also commonly reported (Fig. 1, Table S7-S9). The proportion of participants reporting severe systemic solicited events/symptoms (out of the total 457 boosted in all three stages) are as follows: malaise and/or fatigue in 2.0-4.5%, myalgia in 0-3.3%, headache in 0.7-3.3%, nausea in 0-2.7%, chills in 0-3.3%, arthralgia in 0.6-2.0% and fever in 0.7-2.7%. Local and systemic reactions were most likely to occur in the 1-3 days after booster vaccination, and there were no clear patterns of reaction frequency for solicited or unsolicited AEs by primary EUA vaccine received or age group (Tables S1-S9).

**Figure 1.**
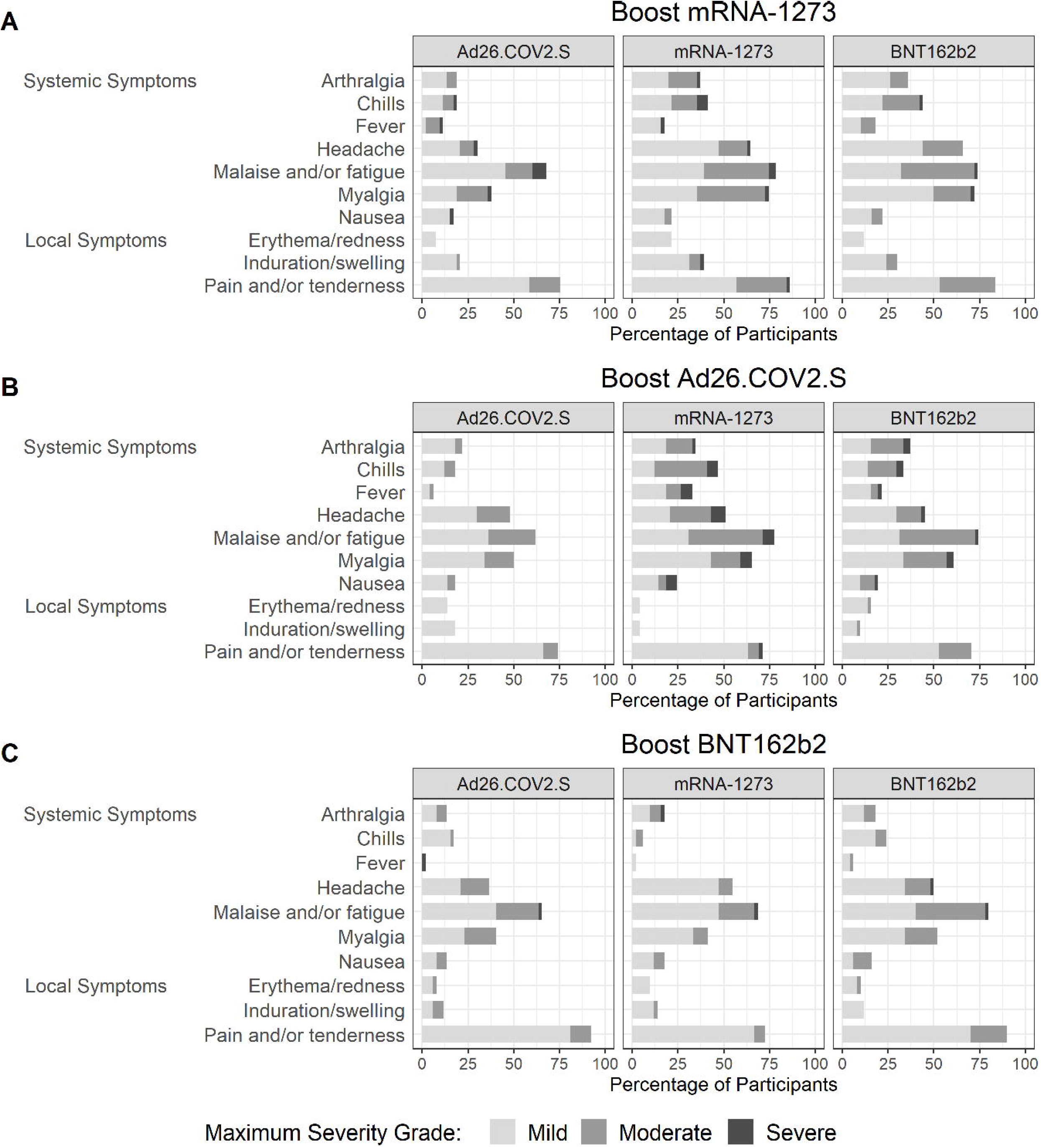
Reactogenicity. Injection site and systemic reactions reported within 7 days after administration of the Ad26.COV2.S, mRNA-1273, and BNT162b2 boosts are depicted by primary EUA immunization regimen. Local and systemic reactions following boost were graded as mild (does not interfere with activity), moderate (interferes with activity) or severe (prevents daily activity).

### Immunogenicity

#### SARS-CoV-2 Binding Antibody Responses

All participants but one (Ad26.COV2.S) had demonstrable bAb prior to vaccine administration (Table 2, Fig. 2). Baseline bAb to the WA-1 strain was 3-15-fold lower in those who received single-dose Ad26.COV2.S compared to the EUA-dosed mRNA vaccines (Table 2, Tables S10-S27). All groups demonstrated an increase in bAb following boost. A 2-fold or greater rise in bAb was noted in 98-100% of Ad26.COV2.S recipients, 96-100% of mRNA-1273 recipients and 98-100% of BNT162b2 recipients following mRNA booster dosing. The geometric mean fold rises in bAb titers ranged from 4.6-56 at Day 15, and were greatest for those who received BNT162b2 and mRNA-1273 boost after an Ad26.COV2.S primary vaccination (33- and 56-fold respectively). The Ad26.COV2.S boost increased bAb titers in all EUA dosed recipients but the Ad26.CoV.2-primed recipients achieved a level 7-10-fold lower compared to those who received an mRNA vaccine priming regimen. Antibody titers to the Delta variant were also evaluated in the 10-plex ECLIA (Tables S28-S33). At baseline, bAb levels to the Delta variant were 34%-45% lower compared with binding of the Wa-1 strain in the same assay. Following boost, all participants had detectable bAb to the Delta variant and the level was only 15-36% lower compared with the Wa-1 strain. Sera from the older and younger age groups had similar bAb levels. Serologic responses to Wa-1 and Beta strains for 4-plex ECLIA (arbitrary units/mL) (Tables S10-S21, S34-39), and WA-1 and Delta on the 10-plex ECLIA are reported in the Supplementary Appendix (Tables S22-S27, S34-S39).

**Table 2.**
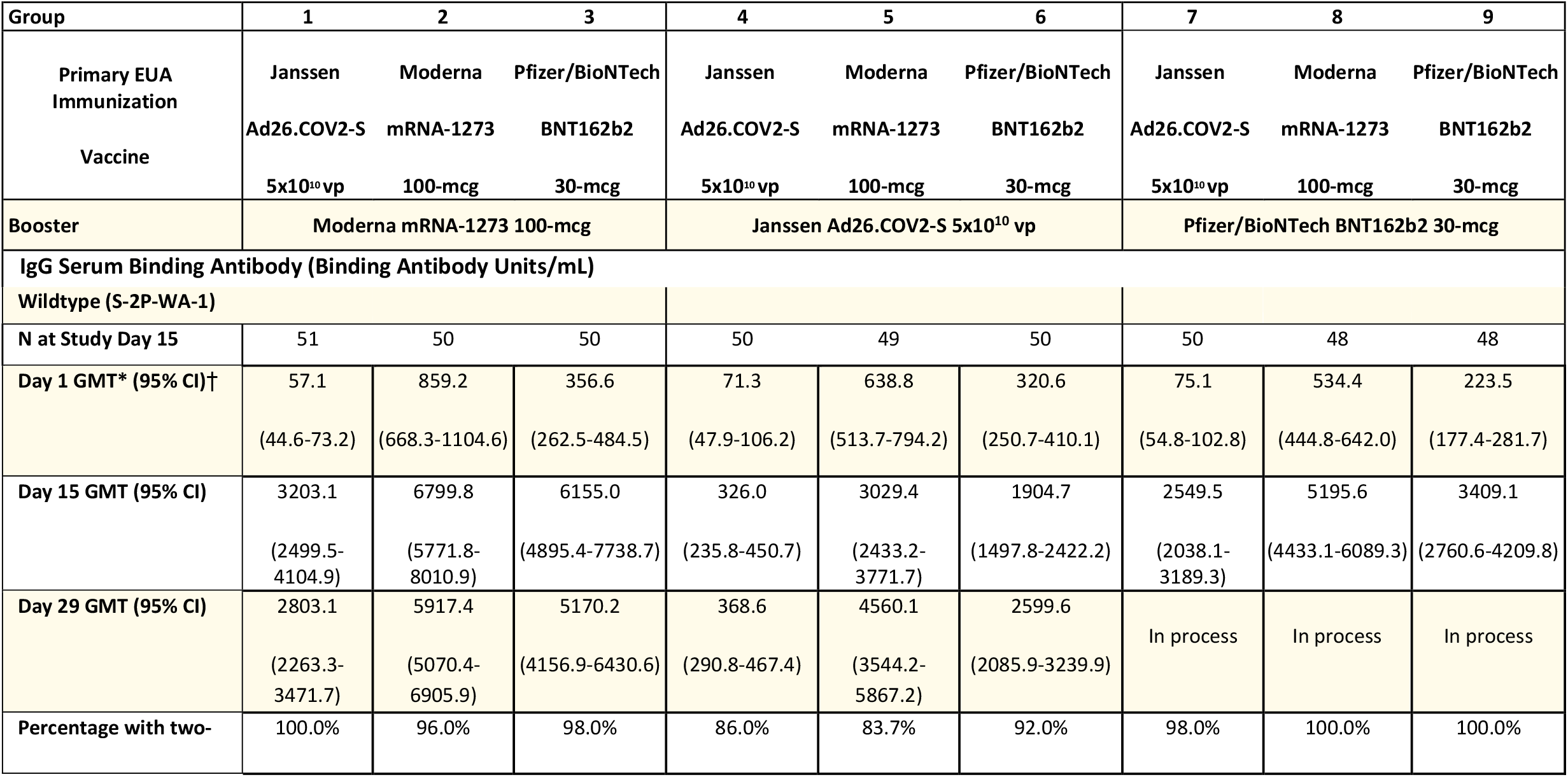
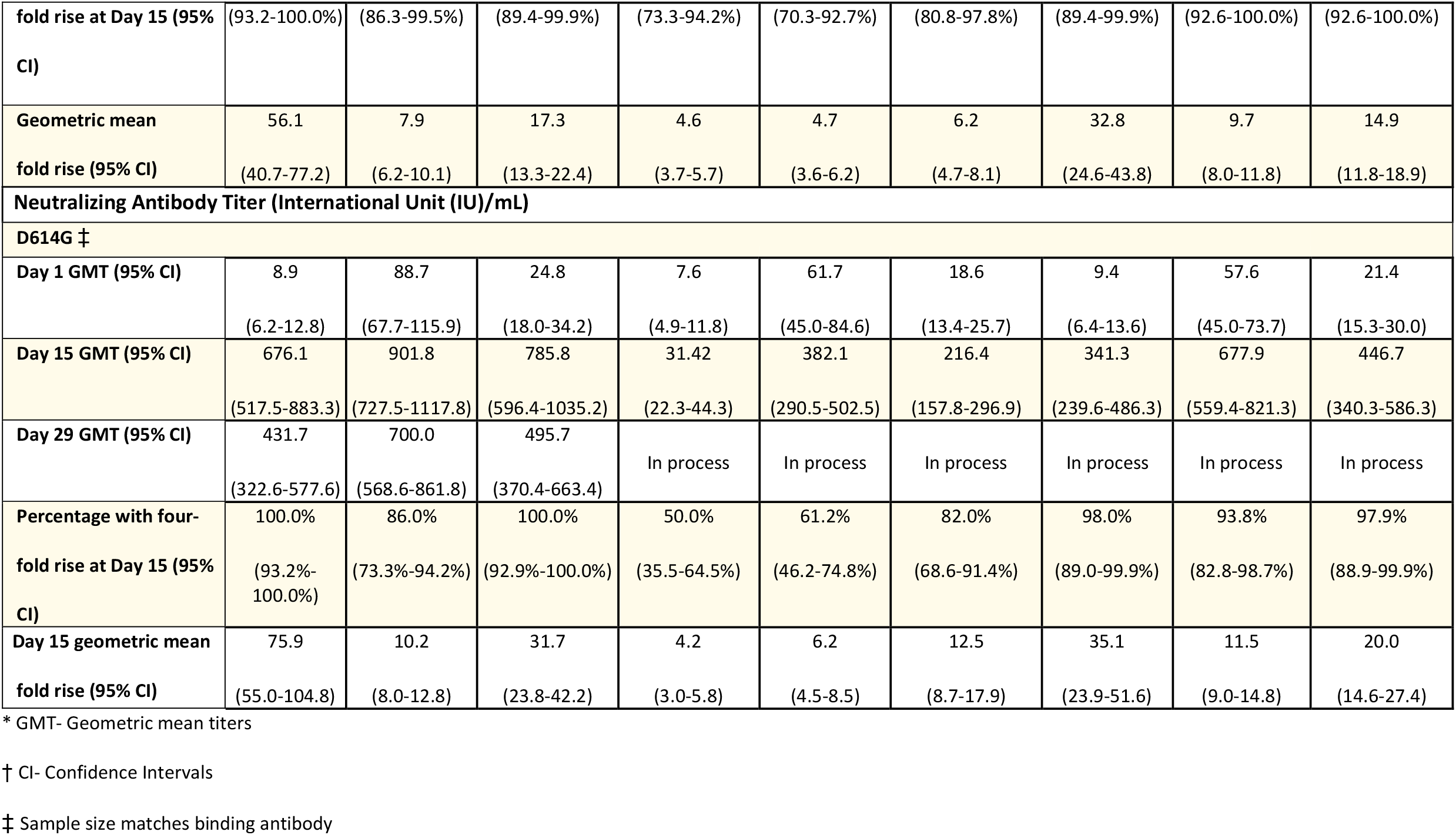
SARS-CoV-2 IgG Binding and Neutralizing Antibody Assays. IgG serum antibody responses to wildtype (S-2P-WA-1) and IU_50_ neutralizing antibody titers to pseudovirus D614G and bridged to international standards and reported as Binding Antibody Units/mL via a 4-plex ECLIA assay and international units ID50/mL (IU50/mL), respectively. Results are reported by Primary immunization EUA Vaccine, Booster vaccine and Timepoint.

**Figure 2.**
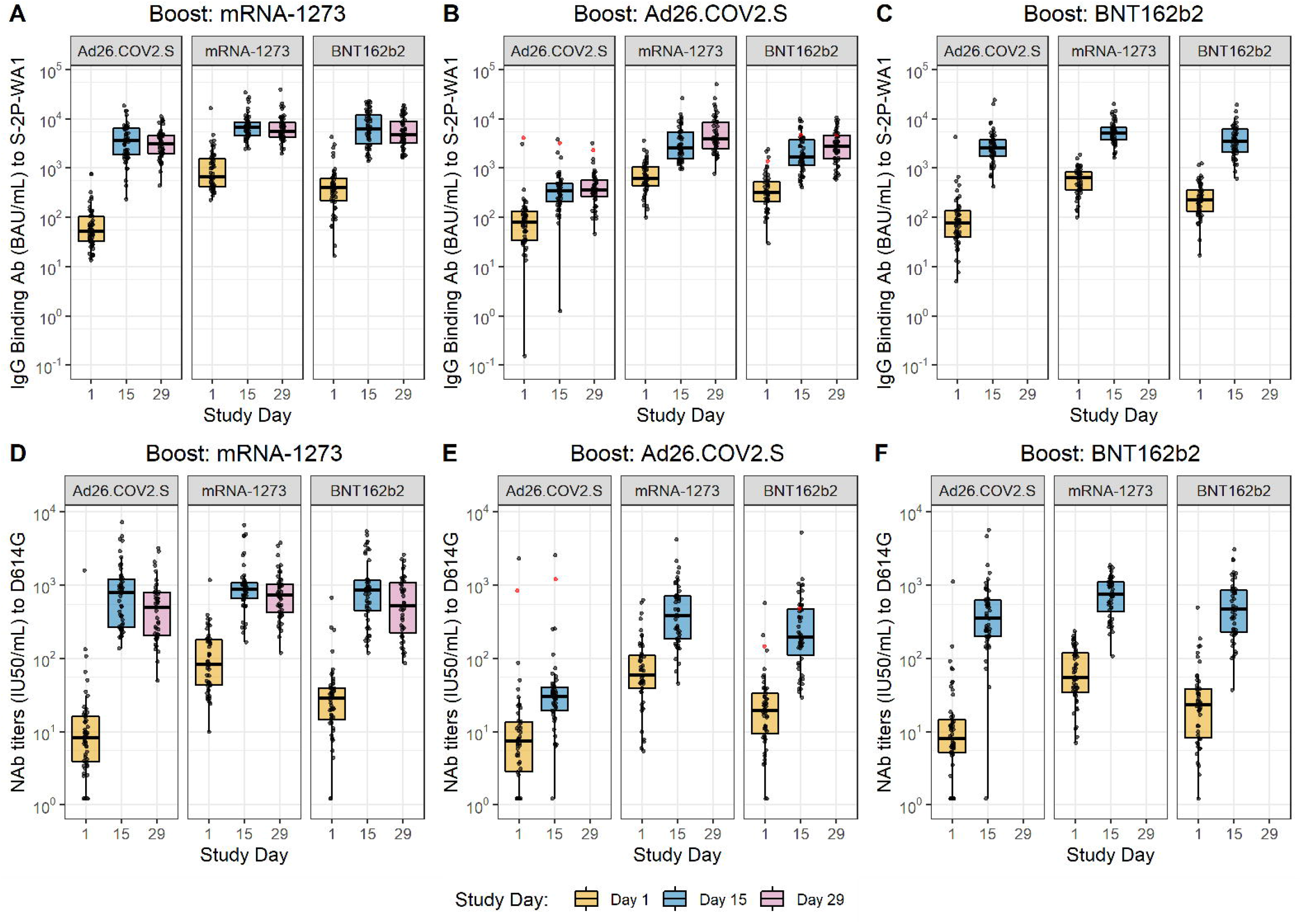
Binding Antibody and Neutralizing Antibody Titers. SARS-CoV-2 IgG binding antibody titers (A-C) and pseudovirus neutralization IU50/mL titers (D-F) at Study Day 1 (baseline), Study Day 15 (post-boost), and Study Day 29 (subset analysis; day 29 data not available for all groups). Binding antibody responses to wildtype (S-2P-WA-1 control), as measured by 4-plex ECLIA, and neutralizing antibody (NAb) levels to D614G, grouped by primary EUA immunization regimen (Ad26.COV2.S, mRNA-1273 and BNT162b2) and booster dosing are depicted. Titers were bridged to international standards and reported as Binding Antibody Units/mL and International Units (IU) 50% inhibitory dose/mL (IU50/mL). Individual values are shown as grey circles. Box plots represent median (horizontal line within the box) and 25^th^ and 75^th^ percentiles (lower and upper borders of the box), and the whiskers drawn to the value nearest to, but within, 1.5 times the interquartile range below and above the 25^th^ and 75^th^ percentile, respectively. The red dots represent participants possessing detectable antibody to the SARS-CoV-2 nucleocapsid protein at enrollment, indicative of a prior SARS-CoV-2 infection.

#### SARS-CoV-2 Neutralizing Antibody Responses

All sera from participants who had received mRNA-1273 as an EUA vaccine had pre-booster neutralizing activity to the D614G strain at study enrollment, while sera from 24 (15.8%) individuals who had received Ad26.COV2.S and five (3.3%) who had received BNT162b2 had no detectable D614G neutralizing activity. Serum neutralization (IU50/mL) levels prior to booster vaccination were approximately three-and ten-fold lower for BNT162b2 and Ad26.COV2.S recipients, respectively, compared to recipients of mRNA-1273, irrespective of interval between EUA vaccination or booster vaccination administered (Table 2, Tables S40-S45). The Study Day 15 post-boost neutralization titers ranged from 676.1-901.8 IU50/ml for participants boosted with mRNA-1273, 31.2-382.2 IU50/ml for those boosted with Ad26.COV2.S, and 341.3-677.9 IU50/mL for those boosted with BNT162b2. The geometric mean fold rises in neutralization titers were greatest for Ad26.COV2.S EUA vaccine recipients, followed by BNT162b2 and mRNA-1273 recipients. In general, Day 15 titers post-boost were highest in mRNA-1273-primed participants, followed by BNT162b2 and Ad26.COV2.S, irrespective of the booster vaccine administered. Persons who received an mRNA-based booster vaccination had a four-fold increase in their neutralization response more frequently than those who were boosted with Ad26.COV.S. Similar findings were observed when IU80/mL neutralization levels were assessed (Tables S46-S51).

At the time of this report, neutralization responses against the Delta and Beta variants were only available in those boosted with mRNA-1273. Overall, pre-booster serum neutralization levels were lower for the Delta and Beta variants than those measured against the D614G variant. All but four participants (two Ad26.COV2.S, two mRNA-1273) and two (one Ad26.COV2.S, one mRNA-1273) persons had four-fold or greater increases in ID50 titers to the Delta and Beta variants, respectively following the booster vaccination; similar findings were observed when ID80 neutralization levels were assessed (Tables S64-S71).

## DISCUSSION

We report interim findings from this clinical trial examining the safety, tolerability and immunogenicity of SARS-CoV-2 booster vaccination in healthy adults who had previously received a full EUA Covid-19 vaccine series. All booster vaccines were immunogenic in subjects irrespective of the primary EUA regimen. The fold increases from baseline in both binding and neutralizing antibody titers were similar or greater after heterologous boosts compared to homologous boosts. Reactogenicity was similar to that described in prior evaluations of Ad26.COV2.S, mRNA-1273 and BNT162b2 vaccines^1-3^ and did not differ between heterologous and homologous boosts. No safety concerns were identified.

Serum binding and neutralizing antibody activity correlates with protection from Covid-19 following mRNA and adenovirus-vectored vaccination.^9,11^ However, these correlates of protection were calculated based on data collected prior to widespread Delta variant circulation, although a recent analysis found that neutralizing antibody levels also correlate with protection from variants of concern, including Delta.^12^ A substantial increase in neutralizing antibody titers was observed in all study participants following booster vaccination irrespective of booster and primary vaccine series.

All groups, with the exception of the homologous Ad26.COV2.S prime-boost group, achieved post-boost neutralizing geometric mean IU50/mL levels of >100, which in a previous study correlated with 90.7% vaccine efficacy.^9^ This cutoff, however, predicts efficacy for preventing symptomatic disease (Covid-19). These data strongly suggest that homologous and heterologous booster vaccine will increase protective efficacy against symptomatic SARS-CoV-2 infection. Predicting vaccine efficacy against severe SARS-CoV-2 infection and death is significantly more challenging, and binding and neutralizing antibodies alone may not reflect measures of protection against these more important outcomes.^12^

The geometric mean neutralizing level of 901.8 IU50/mL after homologous boost of the mRNA-1273 EUA vaccine recipients was substantially higher than that achieved (GMT 247 IU50/mL) approximately four weeks after completion of the primary two-dose mRNA-1273 vaccine series among persons who did not develop Covid-19,^9^ indicating a robust anamnestic response following booster vaccination. Similar responses have been reported after homologous boosting with mRNA-1273, BNT162b2, and Ad26.COV2.S vaccines.^13,21,22^ Neutralizing activity against the Delta and Beta variants also increased substantially following booster vaccination among those evaluated in Stage 1. Many of these study participants did not have detectable neutralizing activity to these variants prior to boost, but all had detectable responses following booster vaccination. The neutralizing activity post-boost against the Delta variant was about 3-fold lower compared to D614G, and this decrement was similar regardless of primary vaccine series. This is similar to the lower neutralization values for Delta compared to D614G seen after primary vaccination with mRNA-1273.^19^ Despite the lower titer against Delta, mRNA-1273 has been shown to be protective against hospitalization and death from infection from the Delta variant.^23^ These data suggest that boosting with the original mRNA-1273 vaccine could maintain continued protection against variants of concern and comports with previous reports.^13,14,21^ Data for the neutralization titers against Delta and Beta after Stage 2 and 3 boosts will soon be available.

Our study has limitations. It was not designed to directly compare responses between different booster regimens. The sample size is insufficient for inter-group comparisons, and the demographics of those studied are not representative of the US population. Volunteers were not randomized into groups, nor stratified with respect to population characteristics or interval from last vaccination. Similarly, the sample size and interim follow-up period were not sufficient to identify rare or late adverse events following booster vaccination, and the interval between completion of the primary series and the booster vaccination evaluated is shorter than the 6 months authorized for BNT162b2. The immunogenicity data are limited to immune responses through Study Day 29, and only serologic responses are reported. The different homologous and heterologous vaccination regimens may vary in terms of cellular and humoral immune responses as well as longevity of the response.

In summary, this preliminary report demonstrates that boosting with any of the three vaccines currently licensed or authorized for emergency use in the US will stimulate an anamnestic response in persons who previously received of the primary series of any of these vaccines. Homologous boosts provided a wide range of immunogenicity responses, with heterologous boosts providing comparable or higher titers. Reactogenicity and adverse events were similar across booster groups. These data suggest that if a vaccine is approved or authorized as a booster, an immune response will be generated regardless of the primary Covid-19 vaccination regimen.

## Disclosure

The results presented here are interim data from an ongoing study and the database is not locked. Data have not been subjected to source verification or standard quality check procedures that would ordinarily occur at database lock. All sites are monitored by independent contractors.

## Data Availability

All available data produced in the present work are contained in the manuscript

## Acknowledgements

The trial was sponsored and primarily funded by the Infectious Diseases Clinical Research Consortium through the National Institute for Allergy and Infectious Diseases (NIAID) of the National Institutes of Health (NIH), under award numbers UM1AI48372, UM1AI148373, UM1AI148450, UM1AI148452, UM1AI148573, UM1AI148574, UM1AI148575, UM1AI148576, UM1AI148684, UM1 AI148689 and with support from the NIAID Collaborative Influenza Vaccine Innovation Centers (CIVICs) contract 75N93019C00050 and NIH Vaccine Research Center.

We would like to acknowledge the efforts of the Safety Monitoring Committee members - Lindsey R. Baden, M.D., Andrew T. Pavia, M.D., and Kawsar Talaat, M.D. Moreover, we would like to acknowledge the volunteers for their willingness to participate in this trial and to Dr. Gregory Deye who provided thoughtful discussions resulting in the early trial design. Additionally, we would like to acknowledge Moderna, Inc., Johnson & Johnson/Janssen, and Pfizer/BioNTech Pharmaceuticals for their collaboration, scientific input, and sharing of documents needed to implement this trial. All products were acquired through the government procurement process.

A complete list of members of the Mix and Match Study Group is provided in the Supplementary Appendix available at.

