## supplemental methods and data for "Heterologous SARS-CoV-2 Booster Vaccinations – Preliminary Report"

This appendix is submitted by the authors to provide additional information about their work.

Supplementary Appendix to Manuscript Entitled

### Heterologous SARS-CoV-2 Booster Vaccinations – Preliminary Report

Table S30: Ad26.COV2.S IgG Serum Antibody to S-2P-B.1.617.2 10-plex.................... .53

Table S31: Ad26.COV2.S IgG Serum Antibody to S-2P-B.1.617.2 10-plex by Age Group…………………………..54
Table S32: BNT162b2 IgG Serum Antibody to S-2P-B.1.617.2 10-plex………………………………………………...55
Table S33: BNT162b2 IgG Serum Antibody to S-2P-B.1.617.2 10-plex by Age Group……………………………...56
Table S34: mRNA-1273 IgG Serum Antibody to S-2P- B.1.351 4-plex ………………………………………………….57
Table S35: mRNA-1273 IgG Serum Antibody to S-2P- B.1.351 4-plex by Age Group ……………………………… 58
Table S36: Ad26.COV2.S IgG Serum Antibody to S-2P- B.1.351 4-plex ……………………………………………...59
Table S37: Ad26.COV2.S IgG Serum Antibody to S-2P- B.1.351 4-plex by Age Group…………………………...60
Table S38: BNT162b2 IgG Serum Antibody to S-2P- B.1.351 4-plex………………………………………………….. 61
Table S39: BNT162b2 IgG Serum Antibody to S-2P- B.1.351 4-plex by Age Group……………………………….62
Table S40: mRNA-1273 SARS-CoV-2 Pseudovirion Neutralization Antibody (I.U. 50) to D614G…………………63
Table S41: mRNA-1273 SARS-CoV-2 Pseudovirion Neutralization Antibody (I.U. 50) to D614G by Age…….64
Table S42: Ad26.COV2.S SARS-CoV-2 Pseudovirion Neutralization Antibody (I.U. 50) to D614G………………65
Table S43: Ad26.COV2.S SARS-CoV-2 Pseudovirion Neutralization Antibody (I.U. 50) to D614G by Age…..66
Table S44: BNT162b2 SARS-CoV-2 Pseudovirion Neutralization Antibody (I.U. 50) to D614G…………………..67
Table S45: BNT162b2 SARS-CoV-2 Pseudovirion Neutralization Antibody (I.U. 50) to D614G by Age……….68
Table S46: mRNA-1273 SARS-CoV-2 Pseudovirion Neutralization Antibody (I.U. 80) to D614G…………………69
Table S47: mRNA-1273 SARS-CoV-2 Pseudovirion Neutralization Antibody (I.U. 80) to D614G by Age…….70
Table S48: Ad26.COV2.S SARS-CoV-2 Pseudovirion Neutralization Antibody (I.U. 80) to D614G………………71
Table S49: Ad26.COV2.S SARS-CoV-2 Pseudovirion Neutralization Antibody (I.U. 80) to D614G by Age…..72
Table S50: BNT162b2 SARS-CoV-2 Pseudovirion Neutralization Antibody (I.U. 80) to D614G…………………..73
Table S51: BNT162b2 SARS-CoV-2 Pseudovirion Neutralization Antibody (I.U. 80) to D614G by Age……….74
Table S52: mRNA-1273 SARS-CoV-2 Pseudovirion Neutralization Antibody (ID50) to D614G……………………75
Table S53: mRNA-1273 SARS-CoV-2 Pseudovirion Neutralization Antibody (ID50) to D614G by Age……….76
Table S54: Ad26.COV2.S SARS-CoV-2 Pseudovirion Neutralization Antibody (ID50) to D614G…………………78
Table S55: Ad26.COV2.S SARS-CoV-2 Pseudovirion Neutralization Antibody (ID50) to D614G by Age……..79
Table S56: BNT162b2 SARS-CoV-2 Pseudovirion Neutralization Antibody (ID50) to D614G……………………..80
Table S57: BNT162b2 SARS-CoV-2 Pseudovirion Neutralization Antibody (ID50) to D614G by Age…………..81
Table S58: mRNA-1273 SARS-CoV-2 Pseudovirion Neutralization Antibody (ID80) to D614G……………………82
Table S59: mRNA-1273 SARS-CoV-2 Pseudovirion Neutralization Antibody (ID80) to D614G by Age……….83
Table S60: Ad26.COV2.S SARS-CoV-2 Pseudovirion Neutralization Antibody (ID80) to D614G…………………85
Table S61: Ad26.COV2.S SARS-CoV-2 Pseudovirion Neutralization Antibody (ID80) to D614G by Age……..86
Table S62: BNT162b2 SARS-CoV-2 Pseudovirion Neutralization Antibody (ID80) to D614G……………………..87
Table S63: BNT162b2 SARS-CoV-2 Pseudovirion Neutralization Antibody (ID80) to D614G by Age………….88
Table S64: mRNA-1273 SARS-CoV-2 Pseudovirion Neutralization Antibody (ID50) to B.1.617.2………………89
Table S65: mRNA-1273 SARS-CoV-2 Pseudovirion Neutralization Antibody (ID50) to B.1.617.2 by Age…. .90
Table S66: mRNA-1273 SARS-CoV-2 Pseudovirion Neutralization Antibody (ID80) to B.1.617.2………………91
Table S67: mRNA-1273 SARS-CoV-2 Pseudovirion Neutralization Antibody (ID80) to B.1.617.2 by Age…. .92
Table S68: mRNA-1273 SARS-CoV-2 Pseudovirion Neutralization Antibody (ID50) to B.1.351…………………93
Table S69: mRNA-1273 SARS-CoV-2 Pseudovirion Neutralization Antibody (ID50) to B.1.351 by Age……...94
Table S70: mRNA-1273 SARS-CoV-2 Pseudovirion Neutralization Antibody (ID80) to B.1.351………………...96
Table S71: mRNA-1273 SARS-CoV-2 Pseudovirion Neutralization Antibody (ID80) to B.1.351 by Age… … .97

**Supplemental Figures** ……………………………………………………………………………………………………………………98
Figure S1: Consort Diagram- Stage 1 mRNA-1273………………………………………………………………………98
Figure S2: Consort Diagram- Stage 2 Ad26.COV2.S……………………………………………………………………99
Figure S3: Consort Diagram- Stage 3 BNT162b2………………………………………………………………………100

**References:**………………………………………………………………………………………………………………………101

#### Mix and Match Study Group
#### (listed in PubMed and ordered by lead institutions and enrollment)

**Baylor College of Medicine (BCM), Houston TX, Vaccine Training and Evaluation Unit (VTEU)**
Robert L. Atmar, M.D., Hana El Sahly, M.D., Jennifer A. Whitaker, M.D., and Wendy A. Keitel, M.D.

**University of Maryland, Institute of Human Virology and Center for Vaccine Development and Global Health, Baltimore, MD, VTEU**
Kirsten E. Lyke, M.D., Meagan Deming, M.D., Ph.D., Jennifer S. Husson, M.D., M.P.H., Angie Price, D.N.P., M.S.N., C.R.N.P.

**Kaiser Permanente, Washington Health Research Institute, Seattle, Washington, VTEU**
Lisa A. Jackson, MD, MPH

**University of Rochester Medical Center, Rochester, NY, VTEU**
Angela Branche, M.D., David Dobryzynski, M.D., Ann R. Falsey, M.D., Ian Shannon, MS, RN,

**The Hope Clinic of Emory University (Hope), Atlanta, GA, VTEU**
Srilatha Edupuganti, M.D., M.P.H., Paulina A. Rebolledo, M.D., M.Sc., Nadine Rouphael, M.D., M.Sc.

**Emory Children's Center (ECC), Atlanta, GA, VTEU**
Christina Rostad, M.D., Andres F. Camacho-Gonzalez, M.D., M.Sc., Satoshi Kamidani, M.D.

**University of Pittsburg School of Medicine, Pittsburg, PA, subsidiary to Vanderbilt University VTEU, Nashville, TN**
Judith Martin, M.D., Kumaravel Rajakumar, M.D., M.S., Gysella B. Muniz M.D., Sonika Bhatnagar, M.D., M.P.H.

**University of Washington, Seattle, WA, VTEU**
Christine Johnston, M.D., M.P.H., Tara M. Babu, M.D., M.SCI., Anna Wald, M.D., M.P.H.

**University of Texas Medical Branch (UTMB), League City, TX, subsidiary to BCM VTEU**
Richard E. Rupp, M.D., Megan Berman, M.D., Laura Porterfield, M.D., Amber Stanford, PA-C

**New York University – Bellevue Vaccine Center, New York, NY, VTEU**
Mark J. Mulligan, M.D., FIDSA, Angelica Cifuentes Kottkamp, M.D., Jennifer Lee Dong, M.D.

**New York University- Long Island Vaccine Center, Mineola, NY, subsidiary to NYU VTEU**
Martín Bäcker, MD, Steven E. Carsons, MD, Diana Badillo, MD,

**Cincinnati Children's Hospital Medical Center (CCHMC), Cincinnati, OH, VTEU**
Rebecca Brady, M.D., Robert W. Frenck, Jr., M.D., Susan Parker, R.N., Michelle Dickey, A.P.R.N.

**Infectious Diseases Clinical Research Consortium (IDCRC) LOU**
Christine M. Posavad, Ph.D., John Hural, Ph.D

**IDCRC Leadership**
Kathleen M. Neuzil, M.D., David Stephens, M.D.

**IDCRC Statistical and Data Science Unit (SDSU):**
Elizabeth R. Brown, Sc.D., Clara Dominguez Islas, Ph.D.

**Statistical Center for HIV/AIDS Research and Prevention (SCHARP), Seattle, WA:**
Jillian Zemanek, M.P.H., Daniel Szydlo, M.S., Rahul Paul Choudhury, MS,

**IDCRC Laboratory Operations Unit (LOU):** Christine M. Posavad, PhD; John Hural, PhD; Michael Stirewalt; Megan Meagher

**Division of Microbiology and Infectious Diseases, National Institute of Allergy and Infectious Diseases, National Institutes of Health, Bethesda, MD.**
Sonja Crandon, B.S.N., Marina Lee, Ph.D., Seema Nayak, M.D., Paul C Roberts, Ph.D., John Beigel, M.D.

**FHI360, Durham, NC**
Janet I. Archer M.Sc.

**NIH Laboratory - Vaccine Research Center, Vaccine Immunology Program (VRC-VIP)**
Adrian McDermott, Ph.D., Sarah O’Connell, M.S., Bob Lin

**Duke University Laboratory – Durham, NC**
David Montefiori, Ph.D., Amanda Eaton, Ph.D.

**Emory University Laboratory (FRNT), Atlanta, GA**
Mehul S. Suthar, Ph.D., Lilin Lai, MD

**Fred Hutch Seattle, WA (ICS)**
M. Juliana McElrath, MD, PhD, Kristen W. Cohen, PhD, Stephen De Rosa, MD

#### Mix and Match Study Team Members

**Baylor College of Medicine (BCM), Houston TX, Vaccine Training and Evaluation Unit (VTEU)**
Robert L. Atmar, M.D.; Hana El Sahly, M.D.; Jennifer A. Whitaker, M.D.; Wendy A. Keitel, M.D.; Mary Healy, M.D.; Christine Akamine, M.D.; Pedro A. Piedra, M.D.; Chanei Henry, A.A.S.; Brandie Phillips, R.N.: Chianti Wade-Bowers, B.S.N, M.S.N.; Connie Rangel, R.N.; Logan Lee; Julia Guardado; Angelica Diaz, F.N.P.; Lisreina Toro; Tina Sierra; Janet Brown, J.D., R.Ph. : Cathy Faw, R.Ph.; Yvette Rugeley; Yolanda Rayford; Kayla Burrell; Jesus Banay; Tykel Eddy; Marinna Matta; Kathy Bosworth, B.A.

**University of Maryland, Institute of Human Virology and Center for Vaccine Development and Global Health, Baltimore, MD, VTEU**
Kirsten E. Lyke, M.D., Meagan E. Deming, M.D., Ph.D., Jennifer S. Husson, M.D., M.P.H., Angie Price, D.N.P., M.S.N., C.R.N.P., Karen Kotloff, M.D., Joel Chua, M.D., Myounghee Lee, Pharm.D., Ph.D., Lisa Anderson, R.N., B.S., Amy Nelson, R.N., M.S., Salma Sharaf, Young Chae Jessica Yoo, Lisa Langer, Pharm.D., Constance Thomas, R.N., Brenda Dorsey, R.N., B.S., M.S., CCRP., Alyson Kwon, M.S., CCRC, ACRP-PM., Sophie Harper, M.Sci., Paula Bernal, M.Sci, Ph.D., Eric Goldstein, Jeffrey Floyd, Regina Harley, M.S., Marcelo Sztein, M.D., Ming Bell, M.D., Leslie Howe, Erika Stiles, Andrew Chi, Pharm.D., Phuong Tran Nguyen, Pharm.D., Yogitha Pazhani, Pharm.D., Christine Aggabao, Pharm.D., Jeannie Murray.

**Kaiser Permanente, Washington Health Research Institute, Seattle, Washington, VTEU**
Lisa A. Jackson, MD, MPH, Lee Barr, R.N., Jesse Berg, B.S., Cassandra Bryant, B.S., Roger Calvert, PA-C, Barbara Carste, M.P.H., Joe Choe, B.S., Maya Dunstan, M.S., R.N., Jana ffitch, L.P.N., Colin Fields, M.D., Lynn Gross, PA-C, Erika Kiniry, M.P.H., Bonnie Lam, PharmD, De Vona Lang, Rebecca Lau, PharmD, Stella Lee, B.A., Paula Lins, PA-C, Amy Mohelnitzky, PA-C, Marilyn Nguyen, B.S., Matthew Nguyen, M.P.H., Hallie Phillips, M.Ed. Stephanie Pimienta, B.S., Melissa Resendiz Rivas, B.A., Melissa Boothe Scheer, PA-C, Janice Suyehira, M.D

**University of Rochester Medical Center, Rochester, NY, VTEU**
Angela R. Branche, MD, David Dobryzynski, MD, Ann R. Falsey, MD, Ian Shannon, MS, RN, Patrick Kingsley, Arthur Zemanek, RN, Sophia Wiltse, Erin Nowicki, Sharon Moorehead, Kari Steinmetz, CCRC, Doreen Francis, RN, CCRC, Cynthia MacDonald, RN, Jeanne Holden-Wiltse, MPH, MBA, Christopher Lane, Elizabeth Fladt, Kyle Richards, PharmD, Stephen Bean, PharmD, Nicole Dornbush, PharmD, Carol Cole, PharmD

**The Hope Clinic of Emory University (Hope), Atlanta, GA, VTEU**
Srilatha Edupuganti, M.D., M.P.H., Paulina A. Rebolledo, M.D., M.Sc., Nadine Rouphael, M.D., M.Sc.,
Alexis Ahonen, M.S.N., Alicarmen Alvarez, R.N., Amy Anderson, R.N., Mary Atha, M.S.N., Sarah Bechnak, R.N., Mary Bower, R.N., Ellie Butler, Laura Clegg, R.N., Carla N. Cooke, P.A., Sharon Curate-Ingram, R.N., Renata Lynn Dennis, M.P.H., R.N., Francine Dyer, R.N., Srilatha Edupuganti, M.D., M.P.H., Tigisty Girmary, R.N., Rebecca Gonzalez, Pharm.D., Daniel Graciaa, M.D., Natalie Gray, Cassie Grimsley-Ackerley, M.D., M.Sc., Lauren Hewitt, L.P.N., Christopher Huerta, Brandi Johnson, Colleen Kelley, M.D., M.P.H., Athena Koumanelis, Deborah Laryea, R.N., Cecilia Losada, Hollie Macenczak, R.N., Michelle Piane McCullough, Eileen Osinski, Bernadine Panganiban, Varun Phadke, M.D., Paulina A. Rebolledo, M.D., M.Sc., Nadine Rouphael, M.D., M.Sc., Erin Scherer, Ph.D., Jessica

Traenkner, P.A., Kristen Unterberger, P.A., Jacob Usher, Michelle Wiles, R.N., T. Jean Winter, Jianguo Xu, R.Ph., Ph.D., Yongxian Xu

**Emory Children's Center (ECC), Atlanta, GA, VTEU**
Christina Rostad, M.D., Andres F. Camacho-Gonzalez, M.D., M.Sc., Satoshi Kamidani, M.D., Evan J. Anderson, M.D., Amari Barrett, Julia Bartol, Leisa Bower, R.N., Jessica Bowman, R.N., Andres F. Camacho-Gonzalez, M.D., M.Sc., Victoria Curry, R.N., Theda Gibson, M.S., Felicia Glover, Hui-Mien Hsiao, M.S., Amberly Hunter, Laila Hussaini, M.P.H., Inara Jooma, Satoshi Kamidani, M.D., Peggy Kettle, R.N., Marcia Lewis, R.N., Wensheng Li, Cindy Lubbers, R.N., Lisa Macoy, R.N., M.S.N., Molly Morrison, Amy Muchinsky, Heather Nurse, R.N., Etza Peters, R.N., Susan Rogers, R.Ph., Christina A. Rostad, M.D., Amber Samuel, Maya Stagg, Kathy Stephens, R.N., M.S.N., Kathryn Zaks, M.S.

**University of Pittsburg School of Medicine, Pittsburg, PA, subsidiary to Vanderbilt University VTEU, Nashville, TN**
Judith M. Martin, M.D., Kumaravel Rajakumar, M.D., M.S., Gysella B. Muniz M.D., Sonika Bhatnagar, M.D., M.P.H., Alejandro Hoberman, MD, Timothy Shope MD, MPH, Nader Shaikh, MD, MPH, Spenser Kinsey BSN RN, Jennifer Opal RN, Melissa Andrasko RN, Shannon Mance MPH BSN RN, Lisa Vavro BSN RN, Lalicia Roman AS BS MS, Kimberly McMurtry PharmD, Emily Dougherty MPH, Kevin Lynch BS, Sabrina Catalano BS, Jamie Fries RN, MaryAnn Sieber RN, John F. Alcorn, Ph.D., Bo Zhai PhD

**University of Washington, Seattle, WA, VTEU**
Christine Johnston, M.D., M.P.H., Tara M. Babu, M.D., M.SCI., Anna Wald, M.D., M.P.H., Morissa Pertik, PA-C, T. Nui Pholsena, ARNP, Jina Taub, ARNP, Dana Varon, ARNP; Meredith Potochnic, PharmD; Matthew Dustrude, Kerry Laing, PhD, David M. Koelle, MD; Alyssa Braun, Anya Mathur, Dolly Singh, Jessica Heimonen, Jessica Moreno, Linsey McClellan, Maddie Humphreys, Ray Larsen, Taylor Krause, Mary Kirk, MPH, Matthew Seymour, MPH, Kirsten Hauge, MPH, Lawrence Hemingway, Christopher McClurkan, Max Krist, Victoria Campbell

**University of Texas Medical Branch (UTMB), League City, TX, subsidiary to BCM VTEU**
Richard E. Rupp, M.D., Megan Berman, M.D., Laura Porterfield, M.D., Amber Stanford, PA-C, Kristin Pollock, RN, Robert Cox, RN, Hala Ghoson, PharmD, Claire Marsh, BSN, RN, Amy McMahan, LVN, Esther Cox, Diane Barrett, MS;

**NYU VETU, New York, NY**
***New York University – Langone Vaccine Center, Manhattan, New York, NY, VTEU***
Mark J. Mulligan, MD, Mary Olson, NP, Marie Samanovic, PhD, Amber Cornelius, MS, Abdonnie Holder, James Wilson, Meron Tasissa, Hibah Khan, Sajjad Hussein, Doaa Ayoubi, Pharm.D., Ph.D., Mahnoor Ali

***NYU Langone Vaccine Center Research Clinic at Bellevue Hospital, New York, NY - subsidiary to NYU VTEU:***
Angelica Kottkamp, MD, Jennifer Dong, MD, Damian Inlall, Ellie Carmody, MD, Rebecca Boas, MD, Jennifer Knishinsky, MD, Reza Parungao, MD, Adam Schwartz, MD, Melinda Katz, MD, Natella Aronova, NP, Rita Mennuti, NP, Athina Agbayani, RN, Zeeshan Iqbal, Julia Wagner, Venissala Wongchai, Tiffany Salcito, Leeja Abraham, Denise Dong, Lance Goodman, Nadia Tadros, Danhong Jiang, Anna Jacobs, Pharm.D., Alina Neganova, RN

***New York University- Long Island Vaccine Center, Mineola, NY, subsidiary to NYU VTEU***
Martín Bäcker, M.D., Steven E. Carsons, M.D., Kimberly Byrnes, R.N.CCRC, Diana Badillo, M.D., Asif Noor, M.D., Sigridh Muñoz-Gómez, M.D., Claudia De La Matta Rodriguez, M.D., Urmee Saha, M.D., Sajumon Joseph, F.N.P., Reny Jose, F.N.P, Catherine Tsiakaros, R.N., Sarah J. Pastolero, R.N., Anita Farhi, R.N., Maung Aung, Alicia Vasile, B.S., R.Ph., April Correll, R.Ph., Louis Ragolia, Ph.D., Christopher Hall, Thomas Palaia, Fiona T. Fitzgerald, Madalyn Saporito, Lavern Harvey.

**Cincinnati Children's Hospital Medical Center (CCHMC), Cincinnati, OH, VTEU**
Rebecca C. Brady, M.D.; Robert W. Frenck, M.D.; Paul W. Spearman, M.D.; Grant C. Paulsen, M.D.;
Eleanor Widdice, M.D.; Felicia A. Scaggs Huang, M.D.; Michelle Dickey, A.P.R.N.; Kristen Buschle, A.P.R.N.; Susan Parker, R.N.; Stacy Ranz, L.P.N.; Tammy Lewis-McCauley, L.P.N.; Margery Huron, R.N.; Sally McCartney, R.N.; Jamie Kidd, R.N.; Jennifer Whitaker, R.N.; Kristie Price, Pharm.D.; Sarah Boland, R.Ph.; Jesse LePage, B.S.; Monica Malone McNeal, M.S.; Laura Pace, B.A.; Theresa Baker, M.S.; Robert Zoellner, B.S.; Mary Pat McKee, M.S.; Nicole Vollman, B.A., Jesse LePage, Kristie Price

**FHI360, Durham, NC**
Janet I. Archer M.Sc., Kuleni Abebe M.Sc. Heather Dickerson B.A., Sandra B. Brindis B.A., Linda McNeil MA PMP, Mary Briggs, Katlyn Hurst B.Sc.

**IDCRC Leadership and Administrative Team**
Kathleen M. Neuzil, M.D., David Stephens, M.D., Monica M. Farley M.D., Jeanne Marrazzo, M.D., Robert L. Atmar M.D., Jeffery Lennox, M.D., Sidnee Paschal Young, MALS; Barbara E. Walsh, Kayla Smith, MPH; and Bridget A. Wynn

**IDCRC Statistical and Data Science Unit (SDSU), Vaccine and Infectious Disease Division, Fred Hutchinson Cancer Research Center, Seattle, WA:**
Elizabeth R. Brown, Sc.D., Clara Dominguez Islas, Ph.D.

**Statistical Center for HIV/AIDS Research and Prevention (SCHARP), Fred Hutchinson Cancer Research Center, Seattle, WA:**
Jillian Zemanek, M.P.H., Daniel Szydlo, M.S., Rahul Paul Choudhury, MS, Brian Ingersoll, Wen-Min (Wendy) Hou, M.P.H., B.S.N, C.C.R.P., Kelly Maddox, Chloe Waters, Lauren Young, Karan A. Shah, M.S., Anisa C. Gravelle, Craig N Chin, Rudie Desravines, M.S., Mark Trumbull, M.S., Srikanth Nooney.

**IDCRC Laboratory Operations Unit (LOU):** Christine M. Posavad, PhD; John Hural, PhD; Michael Stirewalt; Megan Meagher

**Division of Microbiology and Infectious Diseases, National Institute of Allergy and Infectious Diseases, National Institutes of Health, Bethesda, MD.**

Marina Lee, Ph.D., Mohamed Elsafy, M.D., Rhonda Pikaart-Tautges, B.S. Janice Arega, M.S., Binh Hoang, R. Ph., Dan Curtin, Olivia Sparer, B.A., Ranjodh Gill M.P.H, Hyung Koo, B.S.N., Elisa Sindall, B.S.N., Sonja Crandon, B.S.N., Seema Nayak, M.D., Paul C Roberts, Ph.D., John Beigel, M.D.

**Thermo Fisher Scientific, Germantown, MD.**
Jim Dunn

**NIH Laboratory - Vaccine Research Center, Vaccine Immunology Program (VRC-VIP)**
Adrian McDermott, Ph.D., Sarah O'Connell, MS., Bob Lin, Sandeep Narpala, Obrimpong Amoa- Awua, Supra Gajjala, Robin Carroll, Mike Castro, Christopher Moore, Mursal Naisan, Muhammed Naqvi, Jennifer Wang

**Duke University Laboratory – Durham, NC**
David Montefiori, Ph.D., Amanda Eaton, Ph.D.

**Emory University Laboratory (FRNT), Atlanta, GA**
Mehul S. Suthar, Ph.D., Lilin Lai, M.D.

**Fred Hutch, Seattle, WA (ICS)**
M. Juliana McElrath, MD, PhD, Kristen W. Cohen, PhD, Stephen De Rosa, MD, Carol Marty, Todd Haight

#### Binding Assay Method Description

##### 10-plex ECLIA

Testing for serum IgG binding to variants of concern was performed using a fit-for-purpose 10-plex MSD method.[1] Multiplexed Plates (96 well) precoated with up to ten antigens per panel are supplied by the manufacturer. On the day of the assay, the plate is blocked for 60 minutes with MSD Blocker A (5% BSA). The blocking solution is washed off and test samples are applied to the wells at 4 dilution (1:100, 1:500, 1:2500 and 1:10,000) unless otherwise specified and allowed to incubate with shaking for two hours. Plates are washed and Sulfo-tag labeled anti IgG antibody is applied to the wells and allowed to associate with complexed coated antigen – sample antibody within the assay wells. Plates are washed to remove unbound detection antibody. A read solution containing ECL substrate is applied to the wells, and the plate is entered into the MSD Sector instrument. A current is applied to the plate and areas of well surface where sample antibody has complexed with coated antigen and labeled reporter will emit light in the presence of the ECL substrate. The MSD Sector instrument quantitates the amount of light emitted and reports this ECL unit response as a result for each sample and standard of the plate. Magnitude of ECL response is directly proportional to the extent of binding antibody in the test article. All calculations are performed within Excel and the GraphPad Prism software, version 7.0. Readouts are provided as Area Under Curve (AUC).

##### 4-plex ECLIA V.2

Testing was performed using the automated 4 plex MSD method. Quantification of IgG concentrations in serum/plasma were performed with a Beckman Biomek based automation platform. This variant 4plex is in the final stages of validation of the S-2P-B.1.351 and confirmation of validation of the S-2P-WA-1, all performed in alignment with the previously validated SARS-CoV-2 3plex as well as the FDA provided guidance on variant validation.

Briefly, MSD SECTOR® plates are precoated by MSD with Wa-1 SARS-CoV-2 spike (S-2P), B.1.351 receptor binding domain (RBD) protein, Nucleocapsid (N) protein and B.1.351 spike protein in each well in a specific spot-designation for each antigen. The assay will be performed with a Beckman Coulter Biomek based automation integration platform including the Biotek 405TS Plate Washer. Serum samples will be heat-inactivated for 30 minutes at 56oC prior to assay. Plates are blocked for 60 minutes at room temperature (RT) with MSD blocker A solution without shaking. Plates are washed and MSD reference standard (calibrator), QC test sample (pool of COVID-19 convalescent sera) and human serum test samples are added to the precoated wells in duplicates in an 8-point dilution series. Reference standard is added in triplicates. MSD Control sera (low, medium and high) are added undiluted in triplicates as per validated assay format. Additional assay controls might be added in triplicates. Samples are incubated at RT for 4 hours with shaking on a Titramax Plate shaker (Heidolph) at 1500 rpm. SARS-CoV-2 specific antibodies present in the sera or controls bind to the coated antigens. Plates are washed to remove unbound antibodies. Antibodies bound to the SARS-CoV-2 viral proteins are detected using an MSD SULFO-TAGTM anti-human IgG detection antibody incubated for 60 minutes at RT and with shaking. Plates are washed and a read solution (MSD GOLDTM read buffer) containing electrochemiluminescence (ECL) substrate is applied to the wells, and the plate is entered into the MSD MESO Sector S 600 detection system. An electric current is applied to the plates and areas of well surface which form antigen-anti human IgG antibody SULFO-TAGTM complex will emit light in the presence of the ECL substrate.

The MSD MESO Sector S 600 detection system quantitates the amount of light emitted and reports the ECL unit response as a result for each test sample, control sample and reference standard of each plate. Analysis is performed with the MSD Discovery Workbench software, Version 4.0. Calculated ECLIA parameters to measure binding antibody activities will include interpolated concentrations or assigned arbitrary units (AU/mL) read from the standard curve. A 4-pl curve was used for the analysis. Data analysis was performed using Microsoft Excel and GraphPad Prism Version 8.0. The WA-1 S2P AU/mL readouts were previously bridged to the WHO International Standard on the validated version 1 4plex.[1] The established conversion factor is applied to the WA-1 S2P data set and data presented as BAU/mL for ease of cross study comparison.

High values of binding Antibody to the SARS-CoV-2 N-protein by 4-plex were classified as potentially indicative of prior infection if they were above 19389.75 AU/mL. The cutoff point was obtained using a reference sample of 89 subjects with reportable binding levels from 100 tested sera samples collected prior to 2009 in healthy individuals, and it was defined as 1.5 times the inter-quartile above the 3$^{rd}$ quartile of the antibody concentrations on the log-scale.

#### Reporting:

For concentrations that are below the Lower Limit of Detection (LLOQ), numeric values equivalent to LLOQ/2 are assigned before and for all descriptive reporting. For concentrations greater than the Upper Limit of Detection (ULOQ), actual values are reported. If actual values above the ULOQ are not provided, observations are replaced with a value equivalent to the ULOQ. Bridging to the WHO standard Binding Antibody Units per milliliter (BAU/mL) was done using a conversion factor of 0.0090, specific to the IgG SARS-CoV-2 Spike antigen.

#### Pseudovirus Neutralization Immunogenicity Method Description

The Duke NAb Laboratory for HIV and COVID-19 Vaccine Research and Development (Duke NAb Lab; PI: Dr. David Montefiori) assessed the magnitude and cross-variant SARS-CoV-2 neutralizing antibody responses in the DMID Protocol #21-0012 by using a fully validated assay in an environment that operates in compliance with Good Clinical Laboratory Practices (GCLP). The endpoint reported here is the SARS-CoV-2-specific neutralizing activity of antibodies in serum. Neutralizing antibodies were assessed with Spike-pseudotyped viruses in 293T/ACE2 cells as a function of reductions in luciferase (Luc) reporter activity.[2] Assays were performed on all samples with Spike-pseudotyped virus SARS-CoV-2 D614G. A subset of samples were assayed with Spike-pseudotyped virus SARS-CoV-2 B.1.617.2 AY.3 (Delta lineage variant) and with Spike-pseudotyped virus SARS-CoV-2 B.1.351 (Beta variant). These viruses were prepared by Dr. Montefiori's laboratory using a lentivirus for pseudotyping and a luciferase reporter gene for quantitative measurements of virus neutralization. Results are reported as Inhibitory Dilution 50 (ID50) and Inhibitory Dilution 80 (ID80) neutralization titers. These values represent the serum dilution that reduces relative luminescence units (RLU) by either 50% or 80% relative to the RLU in the virus control wells after subtraction of background RLU. This study was completed under the oversight of the Quality Assurance Unit for Duke Vaccine Immunogenicity Programs (QADVIP).

Neutralization of SARS-CoV-2 Spike-pseudotyped viruses was assessed in 293T/ACE2 cells as described in SOP "CFAR02-A0026 Measuring Neutralizing Antibodies Against SARS-CoV-2 Using Pseudotyped Virus and 293T/ACE2 Cells." This assay has been formally validated. Assay validation was performed with human serum samples and monoclonal antibodies using the D614G form of the Wuhan-1 Spike. This assay is in the process of being validated for B.1.351 but has not been validated using B.1.617.2, although the FDA recently allowed assay use for B.1.617.2. The assay is performed in 96-well flat-bottom clear standard non-coated or Poly-L-Lysine treated culture plates for high throughput capacity. Relative luminescence units are measured in 96-well flat bottom black/white plates for enhanced luminescence with minimal bleed-over. Use of a clonal cell line provided enhanced precision and uniformity.

SARS-CoV-2 Spike-pseudotyped viruses are prepared and titrated for infectivity by using mutated forms of an expression plasmid encoding codon-optimized full-length Spike of the Wuhan-1 strain (VRC7480) provided by Drs. Barney Graham and Kizzmekia Corbett at the Vaccine Research Center, National Institutes of Health (USA). Mutations were introduced into VRC7480 by site-directed mutagenesis using the QuikChange Lightning Site-Directed Mutagenesis Kit from Agilent Technologies. All mutations were confirmed by full-length Spike gene sequencing. The variants used here are:

| **Variant** | **Spike mutations** |
|---|---|
| D614G | D614G |
| B.1.617.2 (Delta) | T19R, G142D, Δ156-157, R158G, L452R, T478K, D614G, P681R, D950N |
| B.1.351 (Beta) | L18F, D80A, D215G, ΔL242-244, K417N, E484K, N501Y, D614G and A701V |

Pseudovirions are produced in HEK 293T/17 cells by transfection using Fugene 6 Transfection Reagent and a combination of Spike plasmid, lentiviral backbone plasmid (pCMV ΔR8.2) and firefly Luc reporter gene plasmid (pHR' CMV Luc) in a 1:17:17 ratio in Opti-MEM (Life Technologies). Transfection mixtures are added to pre-seeded HEK 293T/17 cells in T-75 flasks containing 12 ml of growth medium or T-225 flask containing 36 ml of growth medium and incubated for 16-20 hours at 37°C. Medium is removed and 15 ml (T-75 flask) or 45 ml (T-225 flask) of fresh growth medium added. Pseudovirus-containing culture medium is collected after an additional 2 days of incubation, clarified of cells by 0.45 µm micron filtration, and stored in aliquots at -80°C. Pseudovirions are titrated for infectious dose (TCID50) by making serial 3-fold or 5-fold dilutions in quadruplicate for a total of 11 dilutions in 96-well flat-bottom clear standard non-coated or poly-L-lysine-coated culture plates. An additional 4 wells serve as background controls; these wells received cells but no virus. Freshly suspended 293T/ACE2.MF cells are added (10,000 cells/well) and incubated for 66-72 hours. Medium is removed by gentle aspiration and 30 µl of Promega 1X lysis buffer added to all wells. After a 10-minute incubation at room temperature, 100-110 µl of Bright-Glo luciferase reagent is added to all wells, mixed, and 105 µl of the mixture added to a black/white plate (Perkin-Elmer). Luminescence is measured using a GloMax Navigator luminometer (Promega). TCID50 is calculated using the method of Reed and Muench.

Neutralization is measured by using lentiviral particles pseudotyped with SARS-CoV-2 Spike and containing a firefly luciferase (Luc) reporter gene for quantitative measurements of infection by relative luminescence units (RLU). A pre-titrated dose of virus is incubated with 8 serial 5-fold dilutions of serum samples in duplicate in a total volume of 150 µl for 1 hr at 37$^{o}$C in 96-well flat-bottom clear standard non-coated or poly-L-lysine-coated culture plates. Cells are detached using TrypLE Select Enzyme solution, suspended in growth medium (100,000 cells/ml) and immediately added to all wells (10,000 cells in 100 µL of growth medium per well). One set of 8 wells receives cells + virus (virus control) and another set of 8 wells receives cells only (background control). After 71-73 hrs of incubation, medium is removed by gentle aspiration and 30 µl of Promega 1X lysis buffer is added to all wells. After a 10 minute incubation at room temperature, 100 µl of Bright-Glo luciferase reagent is added to all wells. After 1-2 minutes, 110 µl of the cell lysate is transferred to a black/white plate. Luminescence is measured using a GloMax Navigator luminometer (Promega). Neutralization titers are the serum dilution at which RLUs are reduced by either 50% (ID50) or 80% (ID80) compared to virus control wells after subtraction of background RLUs. In some cases, ID50 and ID80 titers were calibrated to the WHO International Standards as described[3] and expressed as IU50/mL and IU80/mL. Serum samples are heat-inactivated for 30 minutes at 56$^{o}$C prior to assay.

For assay internal quality controls (IQC), a qualified positive control is tested on each assay plate. The positive control used for assays with D614G is DH1043NHS, which consists of a potent RBD-specific neutralizing monoclonal antibody (mAb), DH1043, diluted in heat inactivated normal human serum (NHS) at 40 µg/ml. The positive control used for assays with B.1.617.2 and B.1.351 is DH1047NHS, which consists of a potent RBD-specific neutralizing mAb, DH1047, diluted in heat inactivated NHS at 100 μg/ml. Both mAbs were produced in the Duke Protein Production Facility with permission for use obtained from Dr. Barton Haynes, Duke Human Vaccine Institute, Duke University Medical Center. DH1043NHS and DH1047NHS aliquots are stored at -80°C until use. Thawed aliquots are stored at 4°C for up to 3 months. Assay run controls consist of COVID-19 convalescent serum samples or SARS-CoV-2 vaccine recipients with high, medium and low ID50 and ID80 titers against the test virus, plus a normal human serum negative control. In addition, internal quality control samples (IQC-High, IQC-Medium, and IQC-Low) from the IQC Program for SARS-CoV-2 Antibody Assay Monitoring (SAAM) program operated by the External Quality Assurance Program Oversight Laboratory (EQAPOL) were included in each experiment. These values were submitted to EQAPOL and statistical summary reports of trendlines were provided using Nelson rules. Levey-Jennings plots were generated quarterly and checked for violations of standards that are based on the Westgard Rules. Violations of the Westgard Rules were investigated.

Specific to Pseudovirus D614G, the assay's lower limit of detection (LLOD), lower limit of quantification (LLOQ) and upper limit of quantification (ULOQ) are as follows:

| <u>ID50</u> | <u>ID80</u> |
|---|---|
| LLOD = 10 | LLOD = 10 |
| LLOQ = 18.5 | LLOQ = 14.3 |
| ULOQ = 45118 | ULOQ = 10232 |

Values below the Lower Limit of Detection (LLOD) are assigned a value equivalent to LLOD/2 = 10/2 = 5. Levels that are reported as above the LLOD but below the Lower Limit of Quantification (LLOQ) are kept as reported. Values that are greater than the upper limit of quantification (ULOQ) are when actual values are provided. If actual values above the ULOQ are not provided, observations are replaced with a value equivalent to the ULOQ.

Selected summaries of neutralizing titers calibrated to the WHO standard International Units (IU50/mL and IU80/mL), are presented. Conversion was done using calibration factors specifically for the SARS-CoV-2 D614G Pseudovirus: a factor of 0.242 for ID50 and a factor of 1.502 for ID80.[3]

**Supplemental Table 1.** Participants experiencing unsolicited AEs after study boost vaccination in Stage 1 (groups 1E to 3E boosted with mRNA-1273) reported overall and for each Preferred Term within System Organ Class (MedDRA code classification), by Group, relationship to study product, and maximum severity (numbers in brackets correspond to severity grades mild, moderate, severe, potentially life-threatening and death, respectively)

| **System Organ Class/MedDRA Preferred Term[1]** | **Group 1E Ad26.COV2.S (n=53)** | | **Group 2E mRNA-1273 (n=51)** | | **Group 3E BNT162b2 (n=50)** | |
|---|---|---|---|---|---|---|
| | **Related** | **Unrelated** | **Related** | **Unrelated** | **Related** | **Unrelated** |
| **Participants with at least one AE[2], overall** | 8 [6,1,1,0,0] | 13 [9,3,0,1,0] | 8 [6,2,0,0,0] | 14 [8,5,1,0,0] | 18 [10,8,0,0,0] | 24 [15,9,0,0,0] |
| Blood and lymphatic system disorders | | | | | | |
| Lymphadenopathy | 0 | 0 | 3 [3,0,0,0,0] | 0 | 1 [1,0,0,0,0] | 0 |
| Ear and labyrinth disorders | | | | | | |
| Ear pain | 0 | 0 | 0 | 0 | 1 [1,0,0,0,0] | 0 |
| Tinnitus | 0 | 0 | 0 | 1 [1,0,0,0,0] | 0 | 0 |
| Eye disorders | | | | | | |
| Ocular pruritus | 0 | 0 | 0 | 0 | 1 [1,0,0,0,0] | 0 |
| Gastrointestinal disorders | | | | | | |
| Cheilitis | 0 | 0 | 0 | 0 | 1 [1,0,0,0,0] | 0 |
| Constipation | 0 | 0 | 0 | 0 | 0 | 1 [0,1,0,0,0] |
| Diarrhea | 1 [0,1,0,0,0] | 0 | 1 [1,0,0,0,0] | 0 | 1 [1,0,0,0,0] | 1 [0,1,0,0,0] |
| Dry mouth | 1 [1,0,0,0,0] | 0 | 0 | 0 | 0 | 0 |
| Gastroesophageal reflux | 0 | 0 | 0 | 0 | 0 | 1 [0,1,0,0,0] |
| Vomiting | 1 [0,0,1,0,0] | 0 | 0 | 0 | 0 | 0 |
| General disorders and administration site conditions | | | | | | |
| Fatigue | 0 | 0 | 0 | 0 | 0 | 1 [1,0,0,0,0] |
| Feeling abnormal | 0 | 0 | 0 | 0 | 0 | 1 [1,0,0,0,0] |
| Injection site pain | 0 | 0 | 1 [1,0,0,0,0] | 0 | 0 | 0 |
| Injection site pruritus | 0 | 0 | 0 | 0 | 1 [1,0,0,0,0] | 0 |
| Injection site warmth | 1 [1,0,0,0,0] | 0 | 0 | 0 | 0 | 0 |
| Noncardiac chest pain | 0 | 0 | 0 | 1 [0,1,0,0,0] | 0 | 0 |
| Thirst | 0 | 0 | 0 | 0 | 0 | 1 [1,0,0,0,0] |
| Vaccination site hematoma | 0 | 1 [1,0,0,0,0] | 0 | 0 | 0 | 0 |

| | | | | | | |
|---|---|---|---|---|---|---|
| Venous puncture site hematoma | 0 | 0 | 0 | 0 | 0 | 1 [1,0,0,0,0] |
| Infections and infestations | | | | | | |
| Otitis externa | 0 | 0 | 0 | 1 [1,0,0,0,0] | 0 | 0 |
| Pharyngitis, streptococcal | 0 | 0 | 0 | 0 | 0 | 1 [0,1,0,0,0] |
| Rhinovirus infection | 0 | 0 | 0 | 0 | 0 | 1 [1,0,0,0,0] |
| Sinusitis | 0 | 0 | 0 | 1 [0,1,0,0,0] | 0 | 0 |
| Tooth abscess | 0 | 1 [0,1,0,0,0] | 0 | 0 | 0 | 0 |
| Upper respiratory tract infection | 0 | 0 | 0 | 0 | 0 | 2 (2,0,0,0,0) |
| Urinary tract infection | 0 | 1 [1,0,0,0,0] | 0 | 0 | 0 | 0 |
| Viral upper respiratory tract infection | 0 | 1 [0,1,0,0,0] | 0 | 0 | 0 | 0 |
| Injury, poisoning and procedural complications | | | | | | |
| Contusion | 0 | 0 | 0 | 0 | 0 | 1 [1,0,0,0,0] |
| Ligament sprain | 0 | 0 | 0 | 1 [1,0,0,0,0] | 0 | 0 |
| Investigations | | | | | | |
| Blood thyroid stimulating hormone increased | 0 | 1 [1,0,0,0,0] | 0 | 0 | 0 | 0 |
| Metabolism and nutrition disorders | | | | | | |
| Decreased appetite | 0 | 0 | 0 | 0 | 1 [0,1,0,0,0] | 0 |
| Musculoskeletal and connective tissue disorders | | | | | | |
| Arthralgia | 0 | 0 | 0 | 0 | 0 | 1 [1,0,0,0,0] |
| Back pain | 0 | 1 [1,0,0,0,0] | 0 | 0 | 0 | 1 [0,1,0,0,0] |
| Flank pain | 0 | 0 | 0 | 1 (0,0,1,0,0) | 0 | 0 |
| Musculoskeletal discomfort | 0 | 0 | 1 [1,0,0,0,0] | 0 | 0 | 0 |
| Myalgia | 0 | 1 [1,0,0,0,0] | 0 | 0 | 0 | 0 |
| Neck pain | 0 | 0 | 0 | 0 | 0 | 1 [0,1,0,0,0] |
| Pain in extremity | 0 | 1 [1,0,0,0,0] | 0 | 0 | 1 [0,1,0,0,0] | 0 |
| Plantar fasciitis | 0 | 0 | 0 | 1 [0,1,0,0,0] | 0 | 0 |
| Nervous System disorders | | | | | | |
| Dizziness | 2 [2,0,0,0,0] | 0 | 0 | 0 | 3 [0,3,0,0,0] | 0 |
| Headache | 0 | 1 [1,0,0,0,0] | 0 | 0 | 0 | 1 [0,1,0,0,0] |
| Paresthesia | 0 | 0 | 0 | 1 [1.0,0,0,0] | 0 | 0 |
| Psychiatric disorders | 0 | 0 | 0 | 0 | 1 [1,0,0,0,0] | 1 [0,1,0,0,0] |
| Insomnia | 0 | 0 | 0 | 0 | 1 [1,0,0,0,0] | 0 |

| | | | | | | |
|---|---|---|---|---|---|---|
| Restlessness | 0 | 0 | 0 | 0 | 0 | 1 [0,1,0,0,0] |
| Renal and urinary disorders | | | | | | |
| Acute kidney injury | 0 | 1 [0,0,0,1,0] | 0 | 0 | 0 | 0 |
| Reproductive system and breast disorders | | | | | | |
| Vaginal hemorrhage | 0 | 0 | 0 | 0 | 1 [1,0,0,0,0] | 1 [1,0,0,0,0] |
| Respiratory, thoracic and mediastinal disorders | | | | | | |
| Asthma | 0 | 0 | 0 | 0 | 0 | 1 [1,0,0,0,0] |
| Cough | 0 | 0 | 0 | 2 [1,1,0,0,0] | 0 | 1 [1,0,0,0,0] |
| Nasal congestion | 0 | 0 | 0 | 1 [1,0,0,0,0] | 0 | 0 |
| Rhinorrhea | 0 | 0 | 0 | 0 | 0 | 1 [0,1,0,0,0] |
| Skin and subcutaneous tissue disorders | | | | | | |
| Dermatitis, atopic | 0 | 0 | 0 | 1 [0,1,0,0,0] | 0 | 0 |
| Hyperhydrosis | 0 | 0 | 0 | 0 | 1 (0,1,0,0,0) | 0 |
| Night sweats | 0 | 0 | 1 [0,1,0,0,0] | 0 | 2 [1,1,0,0,0] | 0 |
| Pruritus | 0 | 0 | 0 | 1 [1,0,0,0,0] | 0 | 0 |
| Psoriasis | 0 | 0 | 0 | 0 | 0 | 1 [1,0,0,0,0] |
| Rash, pruritic | 0 | 0 | 1 [0,1,0,0,0] | 0 | 0 | 0 |
| Sensitive skin | 0 | 0 | 0 | 0 | 1 [1,0,0,0,0] | 0 |
| Solar lentigo | 0 | 0 | 0 | 0 | 0 | 1 [1,0,0,0,0] |
| Vascular disorders | | | | | | |
| Hypertension | 2 [2,0,0,0,0] | 2 [1,1,0,0,0] | 0 | 0 | 0 | 0 |

[1] Only those AEs which have been assigned MedDRA codes by clinical staff are summarized within organ system class. A total of 4 AEs had not been assigned MeDRA codes at the time of the data snapshot for these groups.

[2] For participants reporting multiple events, overall or withing the same MedDRA term, the maximum severity grade is counted.

**Supplemental Table 2.** Participants experiencing unsolicited AEs after study boost vaccination in Stage 2 (groups 4E to 6E boosted with Ad26.COV2.S) reported overall and for each Preferred Term within System Organ Class (MedDRA code classification), by Group relationship to study product, and maximum severity (numbers in brackets correspond to severity grades mild, moderate, severe, potentially life-threatening and death, respectively)

| **System Organ Class/MedDRA Preferred Term[1]** | **Group 4E Ad26.COV2.S (n=50)** | | **Group 5E mRNA-1273 (n=49)** | | **Group 6E BNT162b2 (n=51)** | |
|---|---|---|---|---|---|---|
| | **Related** | **Unrelated** | **Related** | **Unrelated** | **Related** | **Unrelated** |
| **Participants with at least one AE[2], overall** | 3 [3,0,0,0,0] | 18 [13,5,0,0,0] | 7 [3,2,2,0,0] | 10 [7,2,0,1,0] | 8 [3,4,1,0,0] | 14 [13,1,0,0,0] |
| Blood and lymphatic system disorders | | | | | | |
| Lymphadenopathy | 0 | 0 | 1 [1,0,0,0,0] | 0 | 1 [0,1,0,0,0] | 0 |
| Anemia | 0 | 0 | 0 | 0 | 0 | 1 [0,1,0,0,0] |
| Cardiac disorders | | | | | | |
| Bradycardia | 0 | 1 [1,0,0,0,0] | 0 | 0 | 0 | 0 |
| Eye disorders | | | | | | |
| Eye pain | 0 | 0 | 0 | 0 | 0 | 1 [1,0,0,0,0] |
| Eye pruritus | 0 | 1 [1,0,0,0,0] | 0 | 0 | 0 | 0 |
| Gastrointestinal disorders | | | | | | |
| Abdominal pain | 0 | 1 [1,0,0,0,0] | 0 | 0 | 0 | 0 |
| Aphthous ulcer | 0 | 0 | 0 | 1 [1,0,0,0,0] | 0 | 0 |
| Constipation | 0 | 1 [1,0,0,0,0] | 0 | 0 | 0 | 0 |
| Diarrhea | 0 | 1 [1,0,0,0,0] | 1 [1,0,0,0,0] | 0 | 0 | 1 [1,0,0,0,0] |
| Mouth swelling | 0 | 0 | 0 | 1 [1,0,0,0,0] | 0 | 0 |
| Oral mucosa erosion | 0 | 0 | 0 | 1 [1,0,0,0,0] | 0 | 0 |
| Toothache | 0 | 0 | 0 | 0 | 0 | 1 [1,0,0,0,0 |
| Vomiting | 0 | 0 | 1 [0,0,1,0,0] | 0 | 0 | 1 [1,0,0,0,0 |
| General disorders and administration site conditions | | | | | | |
| Axillary pain | 0 | 0 | 1 [1,0,0,0,0] | 0 | 0 | 0 |
| Chest discomfort | 0 | 1 [1,0,0,0,0] | 0 | 0 | 0 | 0 |
| Cyst | 0 | 1 [1,0,0,0,0] | 0 | 0 | 0 | 0 |

| | | | | | | |
|---|---|---|---|---|---|---|
| Fatigue | 0 | 1 [0,1,0,0,0] | 1 [0,0,1,0,0] | 1 [1,0,0,0,0] | 0 | 0 |
| Feeling abnormal | 0 | 0 | 1 [0,1,0,0,0] | 0 | 2 [0,1,1,0,0] | 0 |
| Injection site bruising | 0 | 0 | 0 | 1 [1,0,0,0,0] | 1 [1,0,0,0,0] | 0 |
| Injection site reaction | 0 | 0 | 0 | 0 | 1 [1,0,0,0,0] | 0 |
| Swelling | 0 | 0 | 1 [1,0,0,0,0] | 0 | 0 | 1 [1,0,0,0,0] |
| Venous puncture site bruise | 0 | 1 [1,0,0,0,0] | 0 | 0 | 0 | 0 |
| Hepatobiliary disorders | | | | | | |
| Acute cholecystitis | 0 | 0 | 0 | 1 [0,0,0,1,0] | 0 | 0 |
| Infections and infestations | | | | | | |
| Covid-19 | 0 | 0 | 0 | 1 [1,0,0,0,0] | 0 | |
| Cellulitis | 0 | 1 [1,0,0,0,0] | 0 | 0 | 0 | 0 |
| Herpes simplex | 0 | 1 [1,0,0,0,0] | 0 | 0 | 0 | 0 |
| Localized infection | 0 | 1 [1,0,0,0,0] | 0 | 0 | 0 | 0 |
| Nasopharyngitis | 0 | 1 [0,1,0,0,0] | 0 | 0 | 0 | 0 |
| Upper respiratory tract infection | 0 | 2 [1,1,0,0,0] | 0 | 1 [1,0,0,0,0] | 0 | 1 [1,0,0,0,0] |
| Injury, poisoning and procedural complications | | | | | | |
| Burn, second degree | 0 | 0 | 0 | 1 [0,1,0,0,0] | 0 | 0 |
| Contusion | 1 [1,0,0,0,0] | 0 | 0 | 0 | 0 | |
| Skin laceration | 0 | 1 [1,0,0,0,0] | 0 | 0 | 0 | 0 |
| Investigations | | | | | | |
| Blood pressure increased | 0 | 0 | 0 | 2 [2,0,0,0,0] | 0 | 0 |
| Metabolism and nutrition disorders | | | | | | |
| Gout | 0 | 0 | 0 | 0 | 1 [0,1,0,0,0] | 0 |
| Musculoskeletal and connective tissue disorders | | | | | | |
| Back pain | 1 [1,0,0,0,0] | 0 | 0 | 0 | 0 | 1 [1,0,0,0,0] |
| Myalgia | 0 | 0 | 0 | 0 | 0 | 1 [1,0,0,0,0] |
| Nervous System disorders | | | | | | |
| Dizziness | 1 [1,0,0,0,0] | 3 [2,1,0,0,0] | 0 | 1 [1,0,0,0,0] | 2 [0,2,0,0,0] | 1 [1,0,0,0,0] |
| Headache | 0 | 0 | 0 | 0 | 0 | 1 [0,1,0,0,0] |
| Hypoaesthesia | 0 | 0 | 0 | 0 | 0 | 1 [0,1,0,0,0] |
| Memory impairment | 0 | 0 | 1 [1,0,0,0,0] | 0 | 0 | 0 |
| Migraine | 0 | 1 [0,1,0,0,0] | 1 [1,0,0,0,0] | 0 | 0 | 0 |

| | | | | | | |
|---|---|---|---|---|---|---|
| Notalgia paresthetica | 0 | 0 | 0 | 1 [1,0,0,0,0] | 0 | 0 |
| Paresthesia | 0 | 0 | 0 | 0 | 0 | 3 [3,0,0,0,0] |
| Syncope | 0 | 0 | 0 | 0 | 0 | 1 [1,0,0,0,0] |
| Psychiatric disorders | | | | | | |
| Anxiety | 0 | 1 [1,0,0,0,0] | 0 | 0 | 0 | 0 |
| Confusional state | 0 | 0 | 0 | 1 [1,0,0,0,0] | 0 | 0 |
| Insomnia | 0 | 0 | 1 [0,1,0,0,0] | 0 | 2 [1,0,1,0,0] | 0 |
| Stress | 0 | 0 | 1 [1,0,0,0,0] | 0 | 0 | 0 |
| Reproductive system and breast disorders | | | | | | |
| Intermenstrual bleeding | 0 | 1 [1,0,0,0,0] | 0 | 0 | 0 | 0 |
| Respiratory, thoracic and mediastinal disorders | | | | | | |
| Cough | 0 | 0 | 0 | 0 | 0 | 1 [1,0,0,0,0] |
| Dyspnea | 0 | 1 [1,0,0,0,0] | 0 | 0 | 0 | 0 |
| Nasal congestion | 0 | 0 | 0 | 0 | 0 | 1 [1,0,0,0,0] |
| Oropharyngeal pain | 0 | 1 [1,0,0,0,0] | 1 [1,0,0,0,0] | 0 | 1 [1,0,0,0,0] | 1 [1,0,0,0,0] |
| Respiratory symptom | 0 | 0 | 0 | 1 [0,1,0,0,0] | 0 | 0 |
| Rhinorrhea | 0 | 0 | 0 | 0 | 0 | 1 [0,1,0,0,0] |
| Sneezing | 0 | 1 [0,1,0,0,0] | 0 | 0 | 0 | 1 [0,1,0,0,0] |
| Throat irritation | 0 | 0 | 0 | 0 | 0 | 1 [0,1,0,0,0] |
| Skin and subcutaneous tissue disorders | | | | | | |
| Blood blister | 0 | 0 | 0 | 0 | 0 | 1 [0,1,0,0,0] |
| Erythema | 0 | 1 [0,1,0,0,0] | 0 | 0 | 0 | 0 |
| Ingrown nail | 0 | 0 | 0 | 0 | 0 | 1 [0,1,0,0,0] |
| Surgical and medical procedures | | | | | | |
| Wisdom teeth removal | 0 | 0 | 0 | 1 [0,1,0,0,0] | 0 | 0 |
| Vascular disorders | | | | | | |
| Diastolic hypertension | 0 | 1 [0,1,0,0,0] | 0 | 1 [0,1,0,0,0] | 0 | 0 |

1 Only those AEs which have been assigned MedDRA codes by clinical staff are summarized within organ system class. All AEs had been assigned MeDRA codes at the time of the data snapshot for these groups.

2 For participants reporting multiple events, overall or withing the same MedDRA term, the maximum severity grade is counted.

**Supplemental Table 3.** Participants experiencing unsolicited AEs after study boost vaccination in Stage 3 (groups 7E to 9E boosted with BNT162b2) reported overall and for each Preferred Term within, System Organ Class (MedDRA code classification), by Group, relationship to study product, and maximum severity (numbers in brackets correspond to severity grades mild, moderate, severe, potentially life-threatening and death, respectively)

| **System Organ Class/MedDRA Preferred Term**[1] | **Group 7E Ad26.COV2.S (n=53)** | | **Group 8E mRNA-1273 (n=51)** | | **Group 9E BNT162b2 (n=50)** | |
|---|---|---|---|---|---|---|
| | **Related** | **Unrelated** | **Related** | **Unrelated** | **Related** | **Unrelated** |
| **Participants with at least one AE**[2]**, overall** | 6 [4,2,0,0,0] | 11 [6,5,0,0,0] | 10 [9,1,0,0,0] | 14 [9,5,0,0,0] | 6 [3,3,0,0,0] | 12 [5,6,1,0,0] |
| Blood and lymphatic system disorders | | | | | | |
| Lymphadenopathy | 2 [2,0,0,0,0] | 0 | 2 [2,0,0,0,0] | 0 | 3 [1,2,0,0,0] | 0 |
| Cardiac disorders | | | | | | |
| Bradycardia | 0 | 1 [1,0,0,0,0] | 0 | 1 [0,1,0,0,0] | 0 | 1 [0,0,1,0,0] |
| Ear and labyrinth disorders | | | | | | |
| Hypoacusis | 0 | 0 | 1 [1,0,0,0,0] | 0 | 0 | 0 |
| Positional vertigo | 0 | 1 [0,1,0,0,0] | 0 | 0 | 0 | 0 |
| Eye disorders | | | | | | |
| Conjunctival hemorrhage | 1 [1,0,0,0,0] | 0 | 0 | 0 | 0 | 0 |
| Gastrointestinal disorders | | | | | | |
| Abdominal pain, upper | 0 | 0 | 0 | 0 | 0 | 1 [0,1,0,0,0] |
| Constipation | 0 | 0 | 0 | 0 | 0 | 1 [0,1,0,0,0] |
| Diarrhea | 0 | 0 | 1 [1,0,0,0,0] | 0 | 0 | 1 [0,1,0,0,0] |
| Oral paresthesia | 0 | 0 | 1 [1,0,0,0,0] | 0 | 0 | 0 |
| General disorders and administration site conditions | | | | | | |
| Axillary pain | 1 [0,1,0,0,0] | 0 | 1 [1,0,0,0,0] | 0 | 1 [1,0,0,0,0] | 0 |
| Cyst | 0 | 1 [0,1,0,0,0] | 0 | 0 | 0 | 0 |
| Injection site bruising | 0 | 0 | 0 | 0 | 0 | 1 [1,0,0,0,0] |
| Injection site pain | 0 | 0 | 1 [0,1,0,0,0] | 0 | 0 | 0 |
| Injection site pruritus | 0 | 0 | 1 [1,0,0,0,0] | 0 | 1 [1,0,0,0,0] | 0 |
| Injection site warmth | 0 | 0 | 1 [1,0,0,0,0] | 0 | 0 | 0 |
| Thirst | 0 | 0 | 0 | 0 | 1 [1,0,0,0,0] | 0 |

| | | | | | | |
|---|---|---|---|---|---|---|
| Vessel puncture site bruise | 0 | 0 | 0 | 2 [2,0,0,0,0] | 0 | 0 |
| Vessel puncture site hemorrhage | 0 | 0 | 0 | 0 | 0 | 1 [0,1,0,0,0] |
| Immune system disorders | | | | | | |
| Seasonal allergy | 0 | 1 [0,1,0,0,0] | 0 | 0 | 0 | 0 |
| Infections and infestations | | | | | | |
| Bacterial keratitis | 0 | 0 | 0 | 0 | 0 | 1 [1,0,0,0,0] |
| Respiratory tract infection | 0 | 0 | 0 | 1 [1,0,0,0,0] | 0 | 0 |
| Upper respiratory tract infection | 0 | 1 [1,0,0,0,0] | 0 | 0 | 0 | 1 [1,0,0,0,0] |
| Injury, poisoning and procedural complications | | | | | | |
| Arthropod bite | 0 | 1 [1,0,0,0,0] | 0 | 0 | 0 | 0 |
| Contusion | 0 | 0 | 0 | 1 [1,0,0,0,0] | 0 | 1 [1,0,0,0,0] |
| Ligament sprain | 0 | 0 | 0 | 1 [1,0,0,0,0] | 0 | 0 |
| Muscle strain | 0 | 1 [1,0,0,0,0] | 0 | 0 | 0 | 0 |
| Nasal injury | 0 | 0 | 0 | 1 [1,0,0,0,0] | 0 | 0 |
| Skin laceration | 0 | 3 [2,1,0,0,0] | 0 | 0 | 0 | 0 |
| Investigations | | | | | | |
| Blood pressure increased | 0 | 0 | 0 | 2 [1,1,0,0,0] | 0 | 0 |
| Heart rate decreased | 0 | 0 | 0 | 1 [1,0,0,0,0] | 0 | 0 |
| Metabolism and nutrition disorders | | | | | | |
| Gout | | | | | | |
| Musculoskeletal and connective tissue disorders | | | | | | |
| Back pain | 0 | 0 | 0 | 2 [0,2,0,0,0] | 0 | 0 |
| Muscle spasms | 0 | 0 | 0 | 1 [0,1,0,0,0] | 0 | 0 |
| Musculoskeletal chest pain | 0 | 0 | 0 | 1 [1,0,0,0,0] | 0 | 0 |
| Musculoskeletal discomfort | 0 | 0 | 0 | 0 | 1 [0,1,0,0,0] | 0 |
| Myalgia | 0 | 0 | 0 | 0 | 0 | 1 [0,1,0,0,0] |
| Osteoporosis | 0 | 1 [0,1,0,0,0] | 0 | 0 | 0 | 0 |
| Pain in extremity | 1 [0,1,0,0,0] | 1 [1,0,0,0,0] | 0 | 0 | 0 | 0 |
| Nervous System disorders | | | | | | |
| Ophthalmic migraine | 0 | 0 | 0 | 1 [1,0,0,0,0] | 0 | 0 |
| Paresthesia | 0 | 0 | 0 | 1 [1,0,0,0,0] | 0 | 0 |
| Product issues | | | | | | |

| | | | | | | |
|---|---|---|---|---|---|---|
| Device physical property issue | 0 | 0 | 0 | 1 [0,1,0,0,0] | 0 | 0 |
| Psychiatric disorders | | | | | | |
| Anxiety | 0 | 0 | 0 | 1 [0,1,0,0,0] | 0 | 1 [1,0,0,0,0] |
| Depression | 0 | 0 | 0 | 1 [0,1,0,0,0] | 0 | 0 |
| Insomnia | 2 [2,0,0,0,0] | 0 | 0 | 0 | 0 | 0 |
| Reproductive system and breast disorders | | | | | | |
| Dysmenorrhea | 0 | 1 [0,1,0,0,0] | 0 | 0 | 0 | 0 |
| Intermenstrual bleeding | 0 | 1 [1,0,0,0,0] | 0 | 0 | 0 | 0 |
| Respiratory, thoracic and mediastinal disorders | | | | | | |
| Cough | 0 | 0 | 0 | 1 [1,0,0,0,0 | 0 | 2 [1,1,0,0,0] |
| Nasal congestion | 0 | 0 | 0 | 0 | 0 | 1 [0,1,0,0,0] |
| Oropharyngeal pain | 0 | 1 [1,0,0,0,0] | 0 | 1 [1,0,0,0,0] | 0 | 0 |
| Paranasal sinus discomfort | 0 | 0 | 0 | 0 | 0 | 1 [0,1,0,0,0] |
| Respiratory disorder | 0 | 0 | 0 | 1 [1,0,0,0,0] | 0 | 0 |
| Throat irritation | 0 | 0 | 0 | 1 [1,0,0,0,0] | 0 | 0 |
| Skin and subcutaneous tissue disorders | | | | | | |
| Contact dermatitis | 0 | 1 [1,0,0,0,0] | 0 | 0 | 0 | 0 |
| Erythema | 0 | 1 [1,0,0,0,0] | 0 | 0 | 0 | 0 |
| Hyperhidrosis | 0 | 0 | 1 [0,1,0,0,0] | 0 | 0 | 0 |
| Rash | 0 | 0 | 1 [1,0,0,0,0] | 0 | 0 | 0 |
| Transient acantholytic dermatosis | 0 | 0 | 0 | 0 | 0 | 1 [1,0,0,0,0] |
| Urticaria | 2 [1,1,0,0,0] | 0 | 0 | 0 | 0 | 0 |
| Vascular disorders | | | | | | |
| Flushing | 0 | 0 | 1 [1,0,0,0,0] | 0 | 0 | 0 |
| Hypertension | 0 | 0 | 0 | 0 | 0 | 1 [1,0,0,0,0] |
| White coat hypertension | 0 | 0 | 0 | 0 | 0 | 1 [0,1,0,0,0] |

[1] Only those AEs which have been assigned MedDRA codes by clinical staff are summarized within organ system class. All AEs had been assigned MeDRA codes at the time of the data snapshot for these groups.

[2] For participants reporting multiple events, overall or withing the same MedDRA term, the maximum severity grade is counted.

Note: One additional AE (Syncope, Nervous System Disorders, Grade 3, not related to boost vaccination) was reported for a participant enrolled into Group 7 but who did not receive the boost vaccination and terminated study early.

**Supplemental Table 4**. Local Solicited Events Stage 1 (groups 1E to 3E boosted with mRNA-1273) by Symptom, Maximum Severity, Age and Enrollment Group

| | Ad26.COV2.S 18-55 years (N=21) | Ad26.COV2.S 56+ years (N=32) | mRNA-1273 18-55 years (N=26) | mRNA-1273 56+ years (N=25) | BNT162b2 18-55 years (N=25) | BNT162b2 56+ years (N=25) | Total (N=154) |
|---|---|---|---|---|---|---|---|
| **Erythema/redness[1]** | | | | | | | |
| None/Not Gradable | 20 (95.2%) | 29 (90.6%) | 22 (84.6%) | 18 (72.0%) | 22 (88.0%) | 22 (88.0%) | 133 (86.4%) |
| Mild | 1 (4.8%) | 3 (9.4%) | 4 (15.4%) | 7 (28.0%) | 3 (12.0%) | 3 (12.0%) | 21 (13.6%) |
| Moderate | 0 (0.0%) | 0 (0.0%) | 0 (0.0%) | 0 (0.0%) | 0 (0.0%) | 0 (0.0%) | 0 (0.0%) |
| Severe | 0 (0.0%) | 0 (0.0%) | 0 (0.0%) | 0 (0.0%) | 0 (0.0%) | 0 (0.0%) | 0 (0.0%) |
| **Erythema/redness largest diameter[2] (cm)** | | | | | | | |
| N | 3 | 3 | 6 | 7 | 4 | 4 | 27 |
| Mean (SD) | 0.8 (0.4) | 4.6 (5.7) | 3.0 (3.7) | 4.2 (4.9) | 0.4 (0.3) | 0.4 (0.4) | 2.5 (3.7) |
| Median | 1.0 | 2.5 | 2.0 | 3.0 | 0.3 | 0.3 | 0.8 |
| 25th, 75th %tile | 0.3, 1.0 | 0.2, 11.0 | 0.2, 3.3 | 0.5, 6.0 | 0.2, 0.6 | 0.1, 0.8 | 0.2, 3.0 |
| Min, Max | 0.3, 1.0 | 0.2, 11.0 | 0.2, 10.0 | 0.2, 14.0 | 0.1, 0.7 | 0.1, 1.0 | 0.1, 14.0 |
| **Induration/swelling[1]** | | | | | | | |
| None/Not Gradable | 17 (81.0%) | 25 (78.1%) | 15 (57.7%) | 16 (64.0%) | 15 (60.0%) | 20 (80.0%) | 108 (70.1%) |
| Mild | 3 (14.3%) | 7 (21.9%) | 8 (30.8%) | 8 (32.0%) | 8 (32.0%) | 4 (16.0%) | 38 (24.7%) |
| Moderate | 1 (4.8%) | 0 (0.0%) | 2 (7.7%) | 1 (4.0%) | 2 (8.0%) | 1 (4.0%) | 7 (4.5%) |
| Severe | 0 (0.0%) | 0 (0.0%) | 1 (3.8%) | 0 (0.0%) | 0 (0.0%) | 0 (0.0%) | 1 (0.6%) |
| **Induration/swelling largest diameter[2] (cm)** | | | | | | | |
| N | 4 | 7 | 11 | 9 | 8 | 5 | 44 |
| Mean (SD) | 3.5 (2.6) | 3.8 (2.3) | 5.5 (4.5) | 3.0 (2.5) | 2.1 (1.7) | 1.9 (2.1) | 3.5 (3.1) |
| Median | 4.0 | 4.0 | 5.3 | 3.0 | 1.5 | 0.8 | 3.0 |
| 25th, 75th %tile | 1.6, 5.5 | 2.0, 6.0 | 2.0, 8.0 | 0.8, 4.0 | 0.7, 4.0 | 0.4, 3.0 | 0.8, 5.0 |
| Min, Max | 0.1, 6.0 | 0.2, 7.0 | 0.6, 15.0 | 0.3, 8.0 | 0.3, 4.0 | 0.1, 5.0 | 0.1, 15.0 |
| **Pain and/or tenderness[1]** | | | | | | | |
| Missing | 0 ( 0.0%) | 0 ( 0.0%) | 0 ( 0.0%) | 0 ( 0.0%) | 0 ( 0.0%) | 1 ( 4.0%) | 1 ( 0.6%) |
| None | 1 ( 4.8%) | 12 ( 37.5%) | 1 ( 3.8%) | 6 ( 24.0%) | 2 ( 8.0%) | 6 ( 24.0%) | 28 ( 18.2%) |
| Mild | 14 ( 66.7%) | 17 ( 53.1%) | 17 ( 65.4%) | 12 ( 48.0%) | 12 ( 48.0%) | 14 ( 56.0%) | 86 ( 55.8%) |
| Moderate | 6 ( 28.6%) | 3 ( 9.4%) | 7 ( 26.9%) | 7 ( 28.0%) | 11 ( 44.0%) | 4 ( 16.0%) | 38 ( 24.7%) |
| Severe | 0 ( 0.0%) | 0 ( 0.0%) | 1 ( 3.8%) | 0 ( 0.0%) | 0 ( 0.0%) | 0 ( 0.0%) | 1 ( 0.6%) |

[1] For a given sign or symptom, each participant's solicited AE will be counted once under the maximum severity for all post-administration assessments.
[2] For erythema/redness and induration/swelling, at each assessment time the maximum diameter at the injection site is recorded. For a given largest diameter, each participant will be counted once under the maximum largest diameter for all post-administration assessments.

**Supplemental Table 5**. Local Solicited Events Stage 2 (groups 4E to 6E boosted with Ad26.COV2.S) by Symptom, Maximum Severity, Age and Enrollment Group

| | Janssen 18-55 (N=26) | Janssen 56+ (N=24) | Moderna 18-55 (N=24) | Moderna 56+ (N=25) | Pfizer 18-55 (N=25) | Pfizer 56+ (N=26) | Total (N=150) |
|---|---|---|---|---|---|---|---|
| **Erythema/redness[1]** | | | | | | | |
| Missing | 0 ( 0.0%) | 0 ( 0.0%) | 0 ( 0.0%) | 0 ( 0.0%) | 0 ( 0.0%) | 0 ( 0.0%) | 0 ( 0.0%) |
| None/Not Gradable | 23 ( 88.5%) | 20 ( 83.3%) | 24 (100.0%) | 23 ( 92.0%) | 20 ( 80.0%) | 23 ( 88.5%) | 133 ( 88.7%) |
| Mild | 3 ( 11.5%) | 4 ( 16.7%) | 0 ( 0.0%) | 2 ( 8.0%) | 5 ( 20.0%) | 2 ( 7.7%) | 16 ( 10.7%) |
| Moderate | 0 ( 0.0%) | 0 ( 0.0%) | 0 ( 0.0%) | 0 ( 0.0%) | 0 ( 0.0%) | 1 ( 3.8%) | 1 ( 0.7%) |
| Severe | 0 ( 0.0%) | 0 ( 0.0%) | 0 ( 0.0%) | 0 ( 0.0%) | 0 ( 0.0%) | 0 ( 0.0%) | 0 ( 0.0%) |
| Potentially life-threatening | 0 ( 0.0%) | 0 ( 0.0%) | 0 ( 0.0%) | 0 ( 0.0%) | 0 ( 0.0%) | 0 ( 0.0%) | 0 ( 0.0%) |
| **Erythema/redness largest diameter[2] (cm)** | | | | | | | |
| N | 3 | 6 | 3 | 3 | 5 | 2 | 22 |
| Mean (SD) | 0.2 (0.1) | 0.6 (0.5) | 0.6 (0.8) | 0.4 (0.5) | 1.6 (1.8) | 1.2 (0.2) | 0.8 (1.0) |
| Median | 0.1 | 0.5 | 0.1 | 0.1 | 0.4 | 1.2 | 0.4 |
| 25th, 75th %tile | 0.1 , 0.3 | 0.2 , 1.0 | 0.1 , 1.5 | 0.1 , 0.9 | 0.2 , 3.0 | 1.0 , 1.3 | 0.1 , 1.0 |
| Min, Max | 0.1 , 0.3 | 0.1 , 1.5 | 0.1 , 1.5 | 0.1 , 0.9 | 0.2 , 4.1 | 1.0 , 1.3 | 0.1 , 4.1 |
| **Induration/swelling[1]** | | | | | | | |
| Missing | 0 ( 0.0%) | 0 ( 0.0%) | 0 ( 0.0%) | 0 ( 0.0%) | 0 ( 0.0%) | 0 ( 0.0%) | 0 ( 0.0%) |
| None/Not Gradable | 20 ( 76.9%) | 21 ( 87.5%) | 23 ( 95.8%) | 24 ( 96.0%) | 23 ( 92.0%) | 23 ( 88.5%) | 134 ( 89.3%) |
| Mild | 6 ( 23.1%) | 3 ( 12.5%) | 1 ( 4.2%) | 1 ( 4.0%) | 2 ( 8.0%) | 2 ( 7.7%) | 15 ( 10.0%) |
| Moderate | 0 ( 0.0%) | 0 ( 0.0%) | 0 ( 0.0%) | 0 ( 0.0%) | 0 ( 0.0%) | 1 ( 3.8%) | 1 ( 0.7%) |
| Severe | 0 ( 0.0%) | 0 ( 0.0%) | 0 ( 0.0%) | 0 ( 0.0%) | 0 ( 0.0%) | 0 ( 0.0%) | 0 ( 0.0%) |
| Potentially life-threatening | 0 ( 0.0%) | 0 ( 0.0%) | 0 ( 0.0%) | 0 ( 0.0%) | 0 ( 0.0%) | 0 ( 0.0%) | 0 ( 0.0%) |
| **Induration/swelling largest diameter[2] (cm)** | | | | | | | |
| N | 6 | 4 | 1 | 3 | 3 | 2 | 19 |
| Mean (SD) | 1.4 (1.6) | 0.7 (0.9) | 0.1 (.) | 0.3 (0.2) | 3.0 (1.1) | 0.7 (0.4) | 1.2 (1.4) |
| Median | 0.6 | 0.3 | 0.1 | 0.2 | 3.0 | 0.7 | 0.4 |
| 25th, 75th %tile | 0.3 , 2.5 | 0.2 , 1.2 | 0.1 , 0.1 | 0.2 , 0.5 | 2.0 , 4.1 | 0.4 , 1.0 | 0.2 , 2.0 |
| Min, Max | 0.1 , 4.2 | 0.2 , 2.0 | 0.1 , 0.1 | 0.2 , 0.5 | 2.0 , 4.1 | 0.4 , 1.0 | 0.1 , 4.2 |

**Supplemental Table 6**. Local Solicited Events Stage 3(groups 7E to 9E boosted with BNT162b2) by Symptom, Maximum Severity, Age and Enrollment Group

| | Janssen 18-55 (N=30) | Janssen 56+ (N=22) | Moderna 18-55 (N=22) | Moderna 56+ (N=29) | Pfizer 18-55 (N=24) | Pfizer 56+ (N=26) | Total (N=153) |
|---|---|---|---|---|---|---|---|
| **Erythema/redness[1]** | | | | | | | |
| Missing | 0 ( 0.0%) | 0 ( 0.0%) | 0 ( 0.0%) | 0 ( 0.0%) | 0 ( 0.0%) | 0 ( 0.0%) | 0 ( 0.0%) |
| None/Not Gradable | 26 ( 86.7%) | 22 (100.0%) | 20 ( 90.9%) | 26 ( 89.7%) | 22 ( 91.7%) | 23 ( 88.5%) | 139 ( 90.8%) |
| Mild | 3 ( 10.0%) | 0 ( 0.0%) | 2 ( 9.1%) | 3 ( 10.3%) | 2 ( 8.3%) | 2 ( 7.7%) | 12 ( 7.8%) |
| Moderate | 1 ( 3.3%) | 0 ( 0.0%) | 0 ( 0.0%) | 0 ( 0.0%) | 0 ( 0.0%) | 1 ( 3.8%) | 2 ( 1.3%) |
| Severe | 0 ( 0.0%) | 0 ( 0.0%) | 0 ( 0.0%) | 0 ( 0.0%) | 0 ( 0.0%) | 0 ( 0.0%) | 0 ( 0.0%) |
| Potentially life-threatening | 0 ( 0.0%) | 0 ( 0.0%) | 0 ( 0.0%) | 0 ( 0.0%) | 0 ( 0.0%) | 0 ( 0.0%) | 0 ( 0.0%) |
| **Erythema/redness largest diameter[2] (cm)** | | | | | | | |
| N | 5 | 2 | 3 | 5 | 3 | 3 | 21 |
| Mean (SD) | 7.6 (7.2) | 0.2 (0.1) | 1.8 (2.3) | 1.8 (2.3) | 1.4 (0.8) | 11.5 (11.8) | 4.4 (6.5) |
| Median | 7.0 | 0.2 | 0.5 | 0.6 | 1.7 | 10.0 | 1.0 |
| 25th, 75th %tile | 1.0 , 12.7 | 0.1 , 0.2 | 0.5 , 4.5 | 0.4 , 2.0 | 0.5 , 2.0 | 0.6 , 24.0 | 0.5 , 5.7 |
| Min, Max | 0.4 , 17.0 | 0.1 , 0.2 | 0.5 , 4.5 | 0.1 , 5.7 | 0.5 , 2.0 | 0.6 , 24.0 | 0.1 , 24.0 |
| **Induration/swelling[1]** | | | | | | | |
| Missing | 0 ( 0.0%) | 0 ( 0.0%) | 0 ( 0.0%) | 0 ( 0.0%) | 0 ( 0.0%) | 0 ( 0.0%) | 0 ( 0.0%) |
| None/Not Gradable | 26 ( 86.7%) | 20 ( 90.9%) | 19 ( 86.4%) | 25 ( 86.2%) | 22 ( 91.7%) | 22 ( 84.6%) | 134 ( 87.6%) |
| Mild | 2 ( 6.7%) | 1 ( 4.5%) | 3 ( 13.6%) | 3 ( 10.3%) | 2 ( 8.3%) | 4 ( 15.4%) | 15 ( 9.8%) |
| Moderate | 2 ( 6.7%) | 1 ( 4.5%) | 0 ( 0.0%) | 1 ( 3.4%) | 0 ( 0.0%) | 0 ( 0.0%) | 4 ( 2.6%) |
| Severe | 0 ( 0.0%) | 0 ( 0.0%) | 0 ( 0.0%) | 0 ( 0.0%) | 0 ( 0.0%) | 0 ( 0.0%) | 0 ( 0.0%) |
| Potentially life-threatening | 0 ( 0.0%) | 0 ( 0.0%) | 0 ( 0.0%) | 0 ( 0.0%) | 0 ( 0.0%) | 0 ( 0.0%) | 0 ( 0.0%) |
| **Induration/swelling largest diameter[2] (cm)** | | | | | | | |
| N | 5 | 2 | 4 | 7 | 3 | 5 | 26 |
| Mean (SD) | 5.2 (3.0) | 0.7 (0.5) | 4.3 (2.8) | 3.7 (3.1) | 1.5 (1.0) | 5.8 (10.3) | 4.0 (4.9) |
| Median | 5.0 | 0.7 | 5.0 | 5.0 | 1.5 | 0.6 | 2.9 |
| 25th, 75th %tile | 3.4 , 6.4 | 0.3 , 1.0 | 2.7 , 6.0 | 0.5 , 7.0 | 0.5 , 2.4 | 0.5 , 3.8 | 0.5 , 5.5 |
| Min, Max | 1.5 , 9.5 | 0.3 , 1.0 | 0.3 , 7.0 | 0.2 , 7.0 | 0.5 , 2.4 | 0.2 , 24.0 | 0.2 , 24.0 |

**Supplemental Table 7.** Systemic Solicited Events Stage 1 (groups 1E to 3E boosted with mRNA-1273) by Symptom, Maximum Severity, Age and Enrollment Group

| | Ad26.COV2.S 18-55 years (N=21) | Ad26.COV2.S 56+ years (N=32) | mRNA-1273 18-55 years (N=26) | mRNA-1273 56+ years (N=25) | BNT162b2 18-55 years (N=25) | BNT162b2 56+ years (N=25) | Total (N=154) |
|---|---|---|---|---|---|---|---|
| **Malaise and/or fatigue**[1] | | | | | | | |
| None | 4 (19.0%) | 13 (40.6%) | 5 ( 19.2%) | 6 ( 24.0%) | 2 ( 8.0%) | 11 ( 44.0%) | 41 ( 26.6%) |
| Mild | 10 (47.6%) | 14 (43.8%) | 10 ( 38.5%) | 10 ( 40.0%) | 11 ( 44.0%) | 5 ( 20.0%) | 60 ( 39.0%) |
| Moderate | 6 (28.6%) | 2 (6.3%) | 10 ( 38.5%) | 8 ( 32.0%) | 11 ( 44.0%) | 9 ( 36.0%) | 46 ( 29.9%) |
| Severe | 1 (4.8%) | 3 (9.4%) | 1 ( 3.8%) | 1 ( 4.0%) | 1 ( 4.0%) | 0 ( 0.0%) | 7 ( 4.5%) |
| **Myalgia**[1] | | | | | | | |
| None | 16 (76.2%) | 17 (53.1%) | 5 ( 19.2%) | 8 ( 32.0%) | 3 ( 12.0%) | 11 ( 44.0%) | 60 ( 39.0%) |
| Mild | 0 (0.0%) | 10 (31.3%) | 8 ( 30.8%) | 10 ( 40.0%) | 15 ( 60.0%) | 10 ( 40.0%) | 53 ( 34.4%) |
| Moderate | 5 (23.8%) | 4 (12.5%) | 12 ( 46.2%) | 7 ( 28.0%) | 6 ( 24.0%) | 4 ( 16.0%) | 38 ( 24.7%) |
| Severe | 0 (0.0%) | 1 (3.1%) | 1 ( 3.8%) | 0 ( 0.0%) | 1 ( 4.0%) | 0 ( 0.0%) | 3 ( 1.9%) |
| **Headache**[1] | | | | | | | |
| None | 13 ( 61.9%) | 24 ( 75.0%) | 4 ( 15.4%) | 14 ( 56.0%) | 5 ( 20.0%) | 12 ( 48.0%) | 72 ( 46.8%) |
| Mild | 6 ( 28.6%) | 5 ( 15.6%) | 14 ( 53.8%) | 10 ( 40.0%) | 14 ( 56.0%) | 8 ( 32.0%) | 57 ( 37.0%) |
| Moderate | 1 ( 4.8%) | 3 ( 9.4%) | 7 ( 26.9%) | 1 ( 4.0%) | 6 ( 24.0%) | 5 ( 20.0%) | 23 ( 14.9%) |
| Severe | 1 ( 4.8%) | 0 ( 0.0%) | 1 ( 3.8%) | 0 ( 0.0%) | 0 ( 0.0%) | 0 ( 0.0%) | 2 ( 1.3%) |
| **Nausea**[1] | | | | | | | |
| None | 15 ( 71.4%) | 29 ( 90.6%) | 19 ( 73.1%) | 21 ( 84.0%) | 18 ( 72.0%) | 21 ( 84.0%) | 123 ( 79.9%) |
| Mild | 5 ( 23.8%) | 3 ( 9.4%) | 7 ( 26.9%) | 2 ( 8.0%) | 6 ( 24.0%) | 2 ( 8.0%) | 25 ( 16.2%) |
| Moderate | 0 ( 0.0%) | 0 ( 0.0%) | 0 ( 0.0%) | 2 ( 8.0%) | 1 ( 4.0%) | 2 ( 8.0%) | 5 ( 3.2%) |
| Severe | 1 ( 4.8%) | 0 ( 0.0%) | 0 ( 0.0%) | 0 ( 0.0%) | 0 ( 0.0%) | 0 ( 0.0%) | 1 ( 0.6%) |
| **Chills**[1] | | | | | | | |
| None | 15 ( 71.4%) | 28 ( 87.5%) | 15 ( 57.7%) | 15 ( 60.0%) | 10 ( 40.0%) | 18 ( 72.0%) | 101 ( 65.6%) |
| Mild | 5 ( 23.8%) | 1 ( 3.1%) | 5 ( 19.2%) | 6 ( 24.0%) | 8 ( 32.0%) | 3 ( 12.0%) | 28 ( 18.2%) |
| Moderate | 1 ( 4.8%) | 2 ( 6.3%) | 4 ( 15.4%) | 3 ( 12.0%) | 6 ( 24.0%) | 4 ( 16.0%) | 20 ( 13.0%) |
| Severe | 0 ( 0.0%) | 1 ( 3.1%) | 2 ( 7.7%) | 1 ( 4.0%) | 1 ( 4.0%) | 0 ( 0.0%) | 5 ( 3.2%) |
| **Arthralgia**[1] | | | | | | | |
| None | 18 ( 85.7%) | 25 ( 78.1%) | 16 ( 61.5%) | 16 ( 64.0%) | 15 ( 60.0%) | 17 ( 68.0%) | 107 ( 69.5%) |
| Mild | 2 ( 9.5%) | 5 ( 15.6%) | 4 ( 15.4%) | 6 ( 24.0%) | 8 ( 32.0%) | 5 ( 20.0%) | 30 ( 19.5%) |
| Moderate | 1 ( 4.8%) | 2 ( 6.3%) | 5 ( 19.2%) | 3 ( 12.0%) | 2 ( 8.0%) | 3 ( 12.0%) | 16 ( 10.4%) |
| Severe | 0 ( 0.0%) | 0 ( 0.0%) | 1 ( 3.8%) | 0 ( 0.0%) | 0 ( 0.0%) | 0 ( 0.0%) | 1 ( 0.6%) |
| **Fever**[1] | | | | | | | |
| None | 18 ( 85.7%) | 29 ( 90.6%) | 21 ( 80.8%) | 21 ( 84.0%) | 18 ( 72.0%) | 23 ( 92.0%) | 130 ( 84.4%) |
| Mild | 0 ( 0.0%) | 1 ( 3.1%) | 4 ( 15.4%) | 4 ( 16.0%) | 4 ( 16.0%) | 1 ( 4.0%) | 14 ( 9.1%) |
| Moderate | 3 ( 14.3%) | 1 ( 3.1%) | 0 ( 0.0%) | 0 ( 0.0%) | 3 ( 12.0%) | 1 ( 4.0%) | 8 ( 5.2%) |
| Severe | 0 ( 0.0%) | 1 ( 3.1%) | 1 ( 3.8%) | 0 ( 0.0%) | 0 ( 0.0%) | 0 ( 0.0%) | 2 ( 1.3%) |

**Supplemental Table 8**. Systemic Solicited Events Stage 2 (groups 4E to 6E boosted with Ad26.COV2.S) by Symptom, Maximum Severity, Age and Enrollment Group

| | Janssen 18-55 (N=26) | Janssen 56+ (N=24) | Moderna 18-55 (N=24) | Moderna 56+ (N=25) | Pfizer 18-55 (N=25) | Pfizer 56+ (N=26) | Total (N=150) |
|---|---|---|---|---|---|---|---|
| **Malaise and/or fatigue[1]** | | | | | | | |
| Missing | 0 ( 0.0%) | 0 ( 0.0%) | 0 ( 0.0%) | 0 ( 0.0%) | 0 ( 0.0%) | 0 ( 0.0%) | 0 ( 0.0%) |
| None | 6 ( 23.1%) | 13 ( 54.2%) | 3 ( 12.5%) | 8 ( 32.0%) | 4 ( 16.0%) | 9 ( 34.6%) | 43 ( 28.7%) |
| Mild | 13 ( 50.0%) | 5 ( 20.8%) | 7 ( 29.2%) | 8 ( 32.0%) | 9 ( 36.0%) | 7 ( 26.9%) | 49 ( 32.7%) |
| Moderate | 7 ( 26.9%) | 6 ( 25.0%) | 14 ( 58.3%) | 6 ( 24.0%) | 12 ( 48.0%) | 9 ( 34.6%) | 54 ( 36.0%) |
| Severe | 0 ( 0.0%) | 0 ( 0.0%) | 0 ( 0.0%) | 3 ( 12.0%) | 0 ( 0.0%) | 1 ( 3.8%) | 4 ( 2.7%) |
| Potentially life-threatening | 0 ( 0.0%) | 0 ( 0.0%) | 0 ( 0.0%) | 0 ( 0.0%) | 0 ( 0.0%) | 0 ( 0.0%) | 0 ( 0.0%) |
| **Myalgia[1]** | | | | | | | |
| Missing | 0 ( 0.0%) | 0 ( 0.0%) | 0 ( 0.0%) | 0 ( 0.0%) | 0 ( 0.0%) | 0 ( 0.0%) | 0 ( 0.0%) |
| None | 9 ( 34.6%) | 16 ( 66.7%) | 6 ( 25.0%) | 11 ( 44.0%) | 6 ( 24.0%) | 14 ( 53.8%) | 62 ( 41.3%) |
| Mild | 11 ( 42.3%) | 6 ( 25.0%) | 10 ( 41.7%) | 11 ( 44.0%) | 9 ( 36.0%) | 8 ( 30.8%) | 55 ( 36.7%) |
| Moderate | 6 ( 23.1%) | 2 ( 8.3%) | 7 ( 29.2%) | 1 ( 4.0%) | 8 ( 32.0%) | 4 ( 15.4%) | 28 ( 18.7%) |
| Severe | 0 ( 0.0%) | 0 ( 0.0%) | 1 ( 4.2%) | 2 ( 8.0%) | 2 ( 8.0%) | 0 ( 0.0%) | 5 ( 3.3%) |
| Potentially life-threatening | 0 ( 0.0%) | 0 ( 0.0%) | 0 ( 0.0%) | 0 ( 0.0%) | 0 ( 0.0%) | 0 ( 0.0%) | 0 ( 0.0%) |
| **Headache[1]** | | | | | | | |
| Missing | 0 ( 0.0%) | 0 ( 0.0%) | 0 ( 0.0%) | 0 ( 0.0%) | 0 ( 0.0%) | 0 ( 0.0%) | 0 ( 0.0%) |
| None | 10 ( 38.5%) | 16 ( 66.7%) | 9 ( 37.5%) | 15 ( 60.0%) | 13 ( 52.0%) | 15 ( 57.7%) | 78 ( 52.0%) |
| Mild | 9 ( 34.6%) | 6 ( 25.0%) | 5 ( 20.8%) | 5 ( 20.0%) | 7 ( 28.0%) | 8 ( 30.8%) | 40 ( 26.7%) |
| Moderate | 7 ( 26.9%) | 2 ( 8.3%) | 8 ( 33.3%) | 3 ( 12.0%) | 4 ( 16.0%) | 3 ( 11.5%) | 27 ( 18.0%) |
| Severe | 0 ( 0.0%) | 0 ( 0.0%) | 2 ( 8.3%) | 2 ( 8.0%) | 1 ( 4.0%) | 0 ( 0.0%) | 5 ( 3.3%) |
| Potentially life-threatening | 0 ( 0.0%) | 0 ( 0.0%) | 0 ( 0.0%) | 0 ( 0.0%) | 0 ( 0.0%) | 0 ( 0.0%) | 0 ( 0.0%) |
| **Nausea[1]** | | | | | | | |
| Missing | 0 ( 0.0%) | 0 ( 0.0%) | 0 ( 0.0%) | 0 ( 0.0%) | 0 ( 0.0%) | 0 ( 0.0%) | 0 ( 0.0%) |
| None | 20 ( 76.9%) | 21 ( 87.5%) | 16 ( 66.7%) | 21 ( 84.0%) | 18 ( 72.0%) | 23 ( 88.5%) | 119 ( 79.3%) |
| Mild | 4 ( 15.4%) | 3 ( 12.5%) | 5 ( 20.8%) | 2 ( 8.0%) | 3 ( 12.0%) | 2 ( 7.7%) | 19 ( 12.7%) |
| Moderate | 2 ( 7.7%) | 0 ( 0.0%) | 2 ( 8.3%) | 0 ( 0.0%) | 3 ( 12.0%) | 1 ( 3.8%) | 8 ( 5.3%) |
| Severe | 0 ( 0.0%) | 0 ( 0.0%) | 1 ( 4.2%) | 2 ( 8.0%) | 1 ( 4.0%) | 0 ( 0.0%) | 4 ( 2.7%) |
| Potentially life-threatening | 0 ( 0.0%) | 0 ( 0.0%) | 0 ( 0.0%) | 0 ( 0.0%) | 0 ( 0.0%) | 0 ( 0.0%) | 0 ( 0.0%) |
| **Chills[1]** | | | | | | | |
| Missing | 0 ( 0.0%) | 0 ( 0.0%) | 0 ( 0.0%) | 0 ( 0.0%) | 0 ( 0.0%) | 0 ( 0.0%) | 0 ( 0.0%) |
| None | 19 ( 73.1%) | 22 ( 91.7%) | 8 ( 33.3%) | 18 ( 72.0%) | 12 ( 48.0%) | 22 ( 84.6%) | 101 ( 67.3%) |
| Mild | 5 ( 19.2%) | 1 ( 4.2%) | 4 ( 16.7%) | 2 ( 8.0%) | 4 ( 16.0%) | 3 ( 11.5%) | 19 ( 12.7%) |
| Moderate | 2 ( 7.7%) | 1 ( 4.2%) | 11 ( 45.8%) | 3 ( 12.0%) | 7 ( 28.0%) | 1 ( 3.8%) | 25 ( 16.7%) |
| Severe | 0 ( 0.0%) | 0 ( 0.0%) | 1 ( 4.2%) | 2 ( 8.0%) | 2 ( 8.0%) | 0 ( 0.0%) | 5 ( 3.3%) |
| Potentially life-threatening | 0 ( 0.0%) | 0 ( 0.0%) | 0 ( 0.0%) | 0 ( 0.0%) | 0 ( 0.0%) | 0 ( 0.0%) | 0 ( 0.0%) |
| **Arthralgia[1]** | | | | | | | |
| Missing | 0 ( 0.0%) | 0 ( 0.0%) | 0 ( 0.0%) | 0 ( 0.0%) | 0 ( 0.0%) | 0 ( 0.0%) | 0 ( 0.0%) |

| | Janssen 18-55 (N=26) | Janssen 56+ (N=24) | Moderna 18-55 (N=24) | Moderna 56+ (N=25) | Pfizer 18-55 (N=25) | Pfizer 56+ (N=26) | Total (N=150) |
|---|---|---|---|---|---|---|---|
| None | 19 ( 73.1%) | 20 ( 83.3%) | 12 ( 50.0%) | 20 ( 80.0%) | 15 ( 60.0%) | 17 ( 65.4%) | 103 ( 68.7%) |
| Mild | 5 ( 19.2%) | 4 ( 16.7%) | 6 ( 25.0%) | 3 ( 12.0%) | 3 ( 12.0%) | 5 ( 19.2%) | 26 ( 17.3%) |
| Moderate | 2 ( 7.7%) | 0 ( 0.0%) | 6 ( 25.0%) | 1 ( 4.0%) | 5 ( 20.0%) | 4 ( 15.4%) | 18 ( 12.0%) |
| Severe | 0 ( 0.0%) | 0 ( 0.0%) | 0 ( 0.0%) | 1 ( 4.0%) | 2 ( 8.0%) | 0 ( 0.0%) | 3 ( 2.0%) |
| Potentially life-threatening | 0 ( 0.0%) | 0 ( 0.0%) | 0 ( 0.0%) | 0 ( 0.0%) | 0 ( 0.0%) | 0 ( 0.0%) | 0 ( 0.0%) |
| **Fever**[1] | | | | | | | |
| Missing | 0 ( 0.0%) | 0 ( 0.0%) | 0 ( 0.0%) | 0 ( 0.0%) | 0 ( 0.0%) | 0 ( 0.0%) | 0 ( 0.0%) |
| None | 24 ( 92.3%) | 23 ( 95.8%) | 13 ( 54.2%) | 20 ( 80.0%) | 18 ( 72.0%) | 22 ( 84.6%) | 120 ( 80.0%) |
| Mild | 1 ( 3.8%) | 1 ( 4.2%) | 6 ( 25.0%) | 3 ( 12.0%) | 6 ( 24.0%) | 2 ( 7.7%) | 19 ( 12.7%) |
| Moderate | 1 ( 3.8%) | 0 ( 0.0%) | 3 ( 12.5%) | 1 ( 4.0%) | 0 ( 0.0%) | 2 ( 7.7%) | 7 ( 4.7%) |
| Severe | 0 ( 0.0%) | 0 ( 0.0%) | 2 ( 8.3%) | 1 ( 4.0%) | 1 ( 4.0%) | 0 ( 0.0%) | 4 ( 2.7%) |
| Potentially life-threatening | 0 ( 0.0%) | 0 ( 0.0%) | 0 ( 0.0%) | 0 ( 0.0%) | 0 ( 0.0%) | 0 ( 0.0%) | 0 ( 0.0%) |

**Supplemental Table 9.** Systemic Solicited Events Stage 3 (groups 7E to 9E boosted with BNT162b2) by Symptom, Maximum Severity, Age and Enrollment Group

| | Janssen 18-55 (N=30) | Janssen 56+ (N=22) | Moderna 18-55 (N=22) | Moderna 56+ (N=29) | Pfizer 18-55 (N=24) | Pfizer 56+ (N=26) | Total (N=153) |
|---|---|---|---|---|---|---|---|
| **Malaise and/or fatigue[1]** | | | | | | | |
| Missing | 0 ( 0.0%) | 0 ( 0.0%) | 0 ( 0.0%) | 0 ( 0.0%) | 0 ( 0.0%) | 0 ( 0.0%) | 0 ( 0.0%) |
| None | 11 ( 36.7%) | 7 ( 31.8%) | 5 ( 22.7%) | 11 ( 37.9%) | 4 ( 16.7%) | 6 ( 23.1%) | 44 ( 28.8%) |
| Mild | 10 ( 33.3%) | 11 ( 50.0%) | 11 ( 50.0%) | 13 ( 44.8%) | 8 ( 33.3%) | 12 ( 46.2%) | 65 ( 42.5%) |
| Moderate | 8 ( 26.7%) | 4 ( 18.2%) | 6 ( 27.3%) | 4 ( 13.8%) | 12 ( 50.0%) | 7 ( 26.9%) | 41 ( 26.8%) |
| Severe | 1 ( 3.3%) | 0 ( 0.0%) | 0 ( 0.0%) | 1 ( 3.4%) | 0 ( 0.0%) | 1 ( 3.8%) | 3 ( 2.0%) |
| Potentially life-threatening | 0 ( 0.0%) | 0 ( 0.0%) | 0 ( 0.0%) | 0 ( 0.0%) | 0 ( 0.0%) | 0 ( 0.0%) | 0 ( 0.0%) |
| **Myalgia[1]** | | | | | | | |
| Missing | 0 ( 0.0%) | 0 ( 0.0%) | 0 ( 0.0%) | 0 ( 0.0%) | 0 ( 0.0%) | 0 ( 0.0%) | 0 ( 0.0%) |
| None | 15 ( 50.0%) | 16 ( 72.7%) | 11 ( 50.0%) | 19 ( 65.5%) | 7 ( 29.2%) | 17 ( 65.4%) | 85 ( 55.6%) |
| Mild | 7 ( 23.3%) | 5 ( 22.7%) | 8 ( 36.4%) | 9 ( 31.0%) | 11 ( 45.8%) | 6 ( 23.1%) | 46 ( 30.1%) |
| Moderate | 8 ( 26.7%) | 1 ( 4.5%) | 3 ( 13.6%) | 1 ( 3.4%) | 6 ( 25.0%) | 3 ( 11.5%) | 22 ( 14.4%) |
| Severe | 0 ( 0.0%) | 0 ( 0.0%) | 0 ( 0.0%) | 0 ( 0.0%) | 0 ( 0.0%) | 0 ( 0.0%) | 0 ( 0.0%) |
| Potentially life-threatening | 0 ( 0.0%) | 0 ( 0.0%) | 0 ( 0.0%) | 0 ( 0.0%) | 0 ( 0.0%) | 0 ( 0.0%) | 0 ( 0.0%) |
| **Headache[1]** | | | | | | | |
| Missing | 0 ( 0.0%) | 0 ( 0.0%) | 0 ( 0.0%) | 0 ( 0.0%) | 0 ( 0.0%) | 0 ( 0.0%) | 0 ( 0.0%) |
| None | 16 ( 53.3%) | 17 ( 77.3%) | 8 ( 36.4%) | 15 ( 51.7%) | 12 ( 50.0%) | 13 ( 50.0%) | 81 ( 52.9%) |
| Mild | 8 ( 26.7%) | 3 ( 13.6%) | 10 ( 45.5%) | 14 ( 48.3%) | 7 ( 29.2%) | 10 ( 38.5%) | 52 ( 34.0%) |
| Moderate | 6 ( 20.0%) | 2 ( 9.1%) | 4 ( 18.2%) | 0 ( 0.0%) | 5 ( 20.8%) | 2 ( 7.7%) | 19 ( 12.4%) |
| Severe | 0 ( 0.0%) | 0 ( 0.0%) | 0 ( 0.0%) | 0 ( 0.0%) | 0 ( 0.0%) | 1 ( 3.8%) | 1 ( 0.7%) |
| Potentially life-threatening | 0 ( 0.0%) | 0 ( 0.0%) | 0 ( 0.0%) | 0 ( 0.0%) | 0 ( 0.0%) | 0 ( 0.0%) | 0 ( 0.0%) |
| **Nausea[1]** | | | | | | | |
| Missing | 0 ( 0.0%) | 0 ( 0.0%) | 0 ( 0.0%) | 0 ( 0.0%) | 0 ( 0.0%) | 0 ( 0.0%) | 0 ( 0.0%) |
| None | 25 ( 83.3%) | 20 ( 90.9%) | 18 ( 81.8%) | 24 ( 82.8%) | 19 ( 79.2%) | 23 ( 88.5%) | 129 ( 84.3%) |
| Mild | 2 ( 6.7%) | 2 ( 9.1%) | 3 ( 13.6%) | 3 ( 10.3%) | 1 ( 4.2%) | 2 ( 7.7%) | 13 ( 8.5%) |
| Moderate | 3 ( 10.0%) | 0 ( 0.0%) | 1 ( 4.5%) | 2 ( 6.9%) | 4 ( 16.7%) | 1 ( 3.8%) | 11 ( 7.2%) |
| Severe | 0 ( 0.0%) | 0 ( 0.0%) | 0 ( 0.0%) | 0 ( 0.0%) | 0 ( 0.0%) | 0 ( 0.0%) | 0 ( 0.0%) |
| Potentially life-threatening | 0 ( 0.0%) | 0 ( 0.0%) | 0 ( 0.0%) | 0 ( 0.0%) | 0 ( 0.0%) | 0 ( 0.0%) | 0 ( 0.0%) |
| **Chills[1]** | | | | | | | |
| Missing | 0 ( 0.0%) | 0 ( 0.0%) | 0 ( 0.0%) | 0 ( 0.0%) | 0 ( 0.0%) | 0 ( 0.0%) | 0 ( 0.0%) |
| None | 25 ( 83.3%) | 18 ( 81.8%) | 20 ( 90.9%) | 28 ( 96.6%) | 16 ( 66.7%) | 22 ( 84.6%) | 129 ( 84.3%) |
| Mild | 4 ( 13.3%) | 4 ( 18.2%) | 0 ( 0.0%) | 1 ( 3.4%) | 6 ( 25.0%) | 3 ( 11.5%) | 18 ( 11.8%) |
| Moderate | 1 ( 3.3%) | 0 ( 0.0%) | 2 ( 9.1%) | 0 ( 0.0%) | 2 ( 8.3%) | 1 ( 3.8%) | 6 ( 3.9%) |
| Severe | 0 ( 0.0%) | 0 ( 0.0%) | 0 ( 0.0%) | 0 ( 0.0%) | 0 ( 0.0%) | 0 ( 0.0%) | 0 ( 0.0%) |
| Potentially life-threatening | 0 ( 0.0%) | 0 ( 0.0%) | 0 ( 0.0%) | 0 ( 0.0%) | 0 ( 0.0%) | 0 ( 0.0%) | 0 ( 0.0%) |
| **Arthralgia[1]** | | | | | | | |
| Missing | 0 ( 0.0%) | 0 ( 0.0%) | 0 ( 0.0%) | 0 ( 0.0%) | 0 ( 0.0%) | 0 ( 0.0%) | 0 ( 0.0%) |
| None | 23 ( 76.7%) | 22 (100.0%) | 18 ( 81.8%) | 24 ( 82.8%) | 20 ( 83.3%) | 21 ( 80.8%) | 128 ( 83.7%) |

| | Janssen 18-55 (N=30) | Janssen 56+ (N=22) | Moderna 18-55 (N=22) | Moderna 56+ (N=29) | Pfizer 18-55 (N=24) | Pfizer 56+ (N=26) | Total (N=153) |
|---|---|---|---|---|---|---|---|
| Mild | 4 ( 13.3%) | 0 ( 0.0%) | 1 ( 4.5%) | 4 ( 13.8%) | 3 ( 12.5%) | 3 ( 11.5%) | 15 ( 9.8%) |
| Moderate | 3 ( 10.0%) | 0 ( 0.0%) | 3 ( 13.6%) | 0 ( 0.0%) | 1 ( 4.2%) | 2 ( 7.7%) | 9 ( 5.9%) |
| Severe | 0 ( 0.0%) | 0 ( 0.0%) | 0 ( 0.0%) | 1 ( 3.4%) | 0 ( 0.0%) | 0 ( 0.0%) | 1 ( 0.7%) |
| Potentially life-threatening | 0 ( 0.0%) | 0 ( 0.0%) | 0 ( 0.0%) | 0 ( 0.0%) | 0 ( 0.0%) | 0 ( 0.0%) | 0 ( 0.0%) |
| **Fever**[1] | | | | | | | |
| Missing | 0 ( 0.0%) | 0 ( 0.0%) | 0 ( 0.0%) | 0 ( 0.0%) | 0 ( 0.0%) | 0 ( 0.0%) | 0 ( 0.0%) |
| None | 29 ( 96.7%) | 22 (100.0%) | 22 (100.0%) | 28 ( 96.6%) | 23 ( 95.8%) | 24 ( 92.3%) | 148 ( 96.7%) |
| Mild | 0 ( 0.0%) | 0 ( 0.0%) | 0 ( 0.0%) | 1 ( 3.4%) | 0 ( 0.0%) | 2 ( 7.7%) | 3 ( 2.0%) |
| Moderate | 0 ( 0.0%) | 0 ( 0.0%) | 0 ( 0.0%) | 0 ( 0.0%) | 1 ( 4.2%) | 0 ( 0.0%) | 1 ( 0.7%) |
| Severe | 1 ( 3.3%) | 0 ( 0.0%) | 0 ( 0.0%) | 0 ( 0.0%) | 0 ( 0.0%) | 0 ( 0.0%) | 1 ( 0.7%) |
| Potentially life-threatening | 0 ( 0.0%) | 0 ( 0.0%) | 0 ( 0.0%) | 0 ( 0.0%) | 0 ( 0.0%) | 0 ( 0.0%) | 0 ( 0.0%) |

Supplemental Table 10: mRNA-1273 IgG Serum Binding Antibody Response to S-2P-WA-1 by 4-plex ECLIA V.2 by Group and Timepoint – Results are reported as Binding Antibody Units (BAU/mL) as bridged to the WHO International Standard

| | Group 1E [Dosed Janssen, Boost Moderna] (N=53) | Group 2E [Dosed Moderna, Boost Moderna] (N=51) | Group 3E [Dosed Pfizer, Boost Moderna] (N=50) |
|---|---|---|---|
| **Day 1 Visit (Pre-boost)** | | | |
| N (non-missing) | 52 | 50 | 50 |
| Median ($P_{25}$, $P_{75}$), BAU/mL | 50.49 (31.82-94.73) | 656.11 (420.07-1479.12) | 401.87 (213.85-638.90) |
| Minimum - Maximum, BAU/mL | 13.39-756.07 | 222.91-16396.44 | 16.59-4312.41 |
| Geometric Mean (95% CI), BAU/mL | 57.11 (44.57-73.18) | 859.21 (668.33-1104.60) | 356.60 (262.48-484.48) |
| **Day 15 Visit (14 days post-boost)** | | | |
| N (non-missing) | 52 | 50 | 50 |
| Median ($P_{25}$, $P_{75}$), BAU/mL | 3480.44 (1800.08-6517.36) | 6531.55 (4628.83-8388.57) | 6213.50 (2912.03-12224.15) |
| Minimum - Maximum, BAU/mL | 233.97-18514.83 | 2457.54-34515.83 | 1396.84-22863.69 |
| Geometric Mean (95% CI), BAU/mL | 3203.11 (2499.45-4104.87) | 6799.76 (5771.77-8010.85) | 6155.01 (4895.42-7738.70) |
| **Day 29 Visit (28 days post-boost)** | | | |
| N (non-missing) | 43 | 48 | 43 |
| Median ($P_{25}$, $P_{75}$), BAU/mL | 2606.39 (1751.32-4569.32) | 5326.58 (4154.75-7935.84) | 4676.78 (3145.07-8931.80) |
| Minimum - Maximum, BAU/mL | 437.76-11147.65 | 1979.01-20379.80 | 1634.84-18726.02 |
| Geometric Mean (95% CI), BAU/mL | 2803.11 (2263.30-3471.68) | 5917.38 (5070.39-6905.85) | 5170.23 (4156.89-6430.60) |

[1] Values below the lower limit of detection (LLOD = 0.3076 BAU/mL) were assigned a value equal to LLOD/2. Values greater than the upper limit of quantification (ULOQ = 172226.25 BAU/mL) were taken as reported, or a ceiling value equivalent to the ULOQ is assigned if values are not provided.

[2] Bridging to the WHO standard Binding Antibody Units per milliliter (BAU/mL) was done using a conversion factor of 0.0090, specific to the IgG SARS-CoV-2 Spike antigen.

#### Supplemental Table 11: mRNA-1273 IgG Serum Binding Antibody Response to S-2P-WA-1 by 4-plex ECLIA V.2 by Group, Age Group and Timepoint – Results are reported as Binding Antibody Units (BAU/mL) as bridged to the WHO International Standard

| | Group 1E [Dosed Janssen, Boost Moderna] Age 18-55 yo (N=21) | Group 1E [Dosed Janssen, Boost Moderna] Age ≥56 yo (N=32) | Group 2E [Dosed Moderna, Boost Moderna] Age 18-55 yo (N=26) | Group 2E [Dosed Moderna, Boost Moderna] Age ≥56 yo (N=25) | Group 3E [Dosed Pfizer, Boost Moderna] Age 18-55 yo (N=25) | Group 3E [Dosed Pfizer, Boost Moderna] Age ≥56 yo (N=25) |
|---|---|---|---|---|---|---|
| **Day 1 Visit (Pre-boost)** | | | | | | |
| N (non-missing) | 21 | 31 | 26 | 24 | 25 | 25 |
| Median ($P_{25}$, $P_{75}$), BAU/mL | 49.30 (29.72-85.56) | 55.11 (33.33-105.18) | 619.47 (420.07-1312.86) | 818.70 (409.44-2006.85) | 444.63 (277.19-718.73) | 360.54 (110.25-486.98) |
| Minimum - Maximum, BAU/mL | 14.85-756.07 | 13.39-755.18 | 278.22-4673.33 | 222.91-16396.44 | 16.59-4312.41 | 26.34-2567.20 |
| Geometric Mean (95% CI), BAU/mL | 56.40 (37.03-85.90) | 57.60 (41.68-79.62) | 776.75 (573.04-1052.87) | 958.45 (625.11-1469.53) | 467.95 (297.17-736.86) | 271.75 (179.47-411.48) |
| **Day 15 Visit (14 days post-boost)** | | | | | | |
| N (non-missing) | 21 | 31 | 26 | 24 | 25 | 25 |
| Median ($P_{25}$, $P_{75}$), BAU/mL | 3596.55 (1546.84-6666.55) | 3158.85 (1963.74-6368.17) | 6778.23 (5465.02-7744.00) | 5752.68 (3875.40-10106.32) | 9108.29 (5219.95-14236.65) | 4010.63 (2545.63-7854.52) |
| Minimum - Maximum, BAU/mL | 233.97-14453.32 | 980.44-18514.83 | 3411.91-24976.08 | 2457.54-34515.83 | 1955.46-22863.69 | 1396.84-21621.37 |
| Geometric Mean (95% CI), BAU/mL | 2777.85 (1693.75-4555.85) | 3527.60 (2695.32-4616.87) | 6840.99 (5821.02-8039.69) | 6755.38 (4960.54-9199.64) | 8195.56 (6160.57-10902.77) | 4622.52 (3291.95-6490.91) |
| **Day 29 Visit (28 days post-boost)** | | | | | | |
| N (non-missing) | 18 | 25 | 25 | 23 | 21 | 22 |
| Median ($P_{25}$, $P_{75}$), BAU/mL | 3213.83 (1999.68-4569.32) | 2282.94 (1421.92-4483.85) | 5317.40 (4335.43-7456.04) | 5335.76 (3659.51-8488.38) | 5896.38 (3714.01-11189.39) | 4015.42 (2507.19-8079.52) |
| Minimum - Maximum, BAU/mL | 1266.67-9196.84 | 437.76-11147.65 | 2456.08-18276.92 | 1979.01-20379.80 | 1634.84-16131.36 | 1705.32-18726.02 |
| Geometric Mean (95% CI), BAU/mL | 3204.53 (2441.98-4205.20) | 2545.61 (1843.69-3514.77) | 6065.29 (4977.93-7390.15) | 5760.70 (4455.03-7449.04) | 5985.27 (4423.96-8097.61) | 4496.00 (3242.82-6233.46) |

[1] Values below the lower limit of detection (LLOD = 0.3076 BAU/mL) were assigned a value equal to LLOD/2. Values greater than the upper limit of quantification (ULOQ = 172226.25 BAU/mL) were taken as reported, or a ceiling value equivalent to the ULOQ is assigned if values are not provided.

[2] Bridging to the WHO standard Binding Antibody Units per milliliter (BAU/mL) was done using a conversion factor of 0.0090, specific to the IgG SARS-CoV-2 Spike antigen.

Supplemental Table 12: Ad26.COV2.S IgG Serum Binding Antibody Response to S-2P-WA-1 by 4-plex ECLIA V.2 by Group and Timepoint – Results are reported as Binding Antibody Units (BAU/mL) as bridged to the WHO International Standard

| | Group 4E [Dosed Janssen, Boost Janssen] (N=50) | Group 5E [Dosed Moderna, Boost Janssen] (N=49) | Group 6E [Dosed Pfizer, Boost Janssen] (N=51) |
|---|---|---|---|
| **Day 1 Visit (Pre-boost)** | | | |
| N (non-missing) | 50 | 49 | 51 |
| Median ($P_{25}$, $P_{75}$), BAU/mL | 79.30 (34.04-138.32) | 620.97 (436.21-1076.63) | 318.33 (195.63-537.74) |
| Minimum - Maximum, BAU/mL | 0.15-4096.29 | 100.43-3578.91 | 29.67-2420.15 |
| Geometric Mean (95% CI), BAU/mL | 71.28 (47.86-106.16) | 638.75 (513.74-794.17) | 320.63 (250.66-410.13) |
| **Day 15 Visit (14 days post-boost)** | | | |
| N (non-missing) | 50 | 49 | 50 |
| Median ($P_{25}$, $P_{75}$), BAU/mL | 349.23 (207.26-486.53) | 2535.08 (1555.66-5370.72) | 1705.55 (1112.86-3816.95) |
| Minimum - Maximum, BAU/mL | 1.25-3788.08 | 959.34-26276.95 | 410.34-10018.47 |
| Geometric Mean (95% CI), BAU/mL | 325.97 (235.76-450.70) | 3029.40 (2433.18-3771.70) | 1904.73 (1497.80-2422.23) |
| **Day 29 Visit (28 days post-boost)** | | | |
| N (non-missing) | 48 | 47 | 50 |
| Median ($P_{25}$, $P_{75}$), BAU/mL | 365.56 (254.31-576.25) | 3849.45 (2305.87-8597.92) | 2819.75 (1554.31-4646.02) |
| Minimum - Maximum, BAU/mL | 46.26-3212.50 | 770.40-50354.53 | 581.17-10641.30 |
| Geometric Mean (95% CI), BAU/mL | 368.64 (290.75-467.40) | 4560.09 (3544.18-5867.21) | 2599.64 (2085.90-3239.90) |

[1] Values below the lower limit of detection (LLOD = 0.3076 BAU/mL) were assigned a value equal to LLOD/2. Values greater than the upper limit of quantification (ULOQ = 172226.25 BAU/mL) were taken as reported, or a ceiling value equivalent to the ULOQ is assigned if values are not provided.

[2] Bridging to the WHO standard Binding Antibody Units per milliliter (BAU/mL) was done using a conversion factor of 0.0090, specific to the IgG SARS-CoV-2 Spike antigen.

Supplemental Table 13: Ad26.COV2.S IgG Serum Binding Antibody Response to S-2P-WA-1 by 4-plex ECLIA V.2 by Group, Age Group and Timepoint – Results are reported as Binding Antibody Units (BAU/mL) as bridged to the WHO International Standard

| | Group 4E [Dosed Janssen, Boost Janssen] Age 18-55 yo (N=26) | Group 4E [Dosed Janssen, Boost Janssen] Age ≥56 yo (N=24) | Group 5E [Dosed Moderna, Boost Janssen] Age 18-55 yo (N=24) | Group 5E [Dosed Moderna, Boost Janssen] Age ≥56 yo (N=25) | Group 6E [Dosed Pfizer, Boost Janssen] Age 18-55 yo (N=25) | Group 6E [Dosed Pfizer, Boost Janssen] Age ≥56 yo (N=26) |
|---|---|---|---|---|---|---|
| **Day 1 Visit (Pre-boost)** | | | | | | |
| N (non-missing) | 26 | 24 | 24 | 25 | 25 | 26 |
| Median ($P_{25}$, $P_{75}$), BAU/mL | 73.68 (34.62-117.27) | 87.70 (31.96-146.83) | 693.82 (416.81-1320.74) | 617.88 (472.54-916.28) | 320.13 (234.49-624.26) | 260.72 (137.53-463.04) |
| Minimum - Maximum, BAU/mL | 13.43-4096.29 | 0.15-3103.22 | 147.62-3578.91 | 100.43-1529.31 | 126.72-2197.49 | 29.67-2420.15 |
| Geometric Mean (95% CI), BAU/mL | 72.89 (47.00-113.05) | 69.57 (33.89-142.81) | 741.01 (527.19-1041.54) | 553.88 (415.67-738.05) | 402.17 (296.94-544.68) | 257.86 (175.28-379.34) |
| **Day 15 Visit (14 days post-boost)** | | | | | | |
| N (non-missing) | 26 | 24 | 24 | 25 | 24 | 26 |
| Median ($P_{25}$, $P_{75}$), BAU/mL | 336.52 (178.33-481.38) | 360.07 (228.24-627.44) | 3038.39 (1716.07-7483.29) | 2423.78 (1506.75-4080.00) | 2056.07 (1155.79-4188.74) | 1596.47 (913.33-2805.14) |
| Minimum - Maximum, BAU/mL | 75.56-3176.44 | 1.25-3788.08 | 968.14-26276.95 | 959.34-12265.54 | 410.34-9043.37 | 459.21-10018.47 |
| Geometric Mean (95% CI), BAU/mL | 326.13 (231.85-458.76) | 325.80 (179.50-591.36) | 3592.36 (2526.38-5108.12) | 2572.13 (1955.19-3383.75) | 2113.48 (1456.86-3066.05) | 1730.39 (1243.98-2406.99) |
| **Day 29 Visit (28 days post-boost)** | | | | | | |
| N (non-missing) | 24 | 24 | 23 | 24 | 25 | 25 |
| Median ($P_{25}$, $P_{75}$), BAU/mL | 351.09 (169.05-552.72) | 372.53 (306.25-626.16) | 4204.09 (2687.00-10786.12) | 3733.10 (1914.13-5759.10) | 2705.64 (1575.88-4862.34) | 2848.98 (1437.32-4026.91) |
| Minimum - Maximum, BAU/mL | 93.31-2288.23 | 46.26-3212.50 | 1549.22-50354.53 | 770.40-26269.63 | 624.93-10641.30 | 581.17-9385.55 |
| Geometric Mean (95% CI), BAU/mL | 342.14 (243.17-481.38) | 397.20 (278.98-565.51) | 5363.02 (3680.67-7814.34) | 3903.67 (2743.65-5554.14) | 2734.70 (1997.17-3744.59) | 2471.25 (1776.11-3438.45) |

[1] Values below the lower limit of detection (LLOD = 0.3076 BAU/mL) were assigned a value equal to LLOD/2. Values greater than the upper limit of quantification (ULOQ = 172226.25 BAU/mL) were taken as reported, or a ceiling value equivalent to the ULOQ is assigned if values are not provided.

[2] Bridging to the WHO standard Binding Antibody Units per milliliter (BAU/mL) was done using a conversion factor of 0.0090, specific to the IgG SARS-CoV-2 Spike antigen.

**Supplemental Table 14:** BNT162b2 IgG Serum Binding Antibody Response to S-2P-WA-1 by 4-plex ECLIA V.2 by Group and Timepoint – Results are reported as Binding Antibody Units (BAU/mL) as bridged to the WHO International Standard

| | Group 7E [Dosed Janssen, Boost Pfizer] (N=53) | Group 8E [Dosed Moderna, Boost Pfizer] (N=51) | Group 9E [Dosed Pfizer, Boost Pfizer] (N=50) |
|---|---|---|---|
| **Day 1 Visit (Pre-boost)** | | | |
| N (non-missing) | 53 | 51 | 50 |
| Median ($P_{25}$, $P_{75}$), BAU/mL | 77.38 (40.56-134.69) | 636.23 (355.94-837.20) | 226.53 (130.59-366.82) |
| Minimum - Maximum, BAU/mL | 5.02-4279.92 | 99.56-1881.23 | 17.04-1234.78 |
| Geometric Mean (95% CI), BAU/mL | 75.07 (54.83-102.78) | 534.37 (444.77-642.02) | 223.52 (177.36-281.69) |
| **Day 15 Visit (14 days post-boost)** | | | |
| N (non-missing) | 50 | 48 | 48 |
| Median ($P_{25}$, $P_{75}$), BAU/mL | 2579.84 (1710.50-3715.63) | 5212.05 (3673.36-6716.43) | 3496.80 (2063.90-6150.76) |
| Minimum - Maximum, BAU/mL | 420.32-24305.59 | 1626.48-20018.49 | 607.22-19207.12 |
| Geometric Mean (95% CI), BAU/mL | 2549.53 (2038.09-3189.32) | 5195.62 (4433.10-6089.29) | 3409.05 (2760.64-4209.76) |

[1] Values below the lower limit of detection (LLOD = 0.3076 BAU/mL) were assigned a value equal to LLOD/2. Values greater than the upper limit of quantification (ULOQ = 172226.25 BAU/mL) were taken as reported, or a ceiling value equivalent to the ULOQ is assigned if values are not provided.

[2] Bridging to the WHO standard Binding Antibody Units per milliliter (BAU/mL) was done using a conversion factor of 0.0090, specific to the IgG SARS-CoV-2 Spike antigen.

**Supplemental Table 15:** BNT162b2 IgG Serum Binding Antibody Response to S-2P-WA-1 by 4-plex ECLIA V.2 by Group, Age Group and Timepoint – Results are reported as Binding Antibody Units (BAU/mL) as bridged to the WHO International Standard

| | Group 7E [Dosed Janssen, Boost Pfizer] Age 18-55 yo (N=31) | Group 7E [Dosed Janssen, Boost Pfizer] Age ≥56 yo (N=22) | Group 8E [Dosed Moderna, Boost Pfizer] Age 18-55 yo (N=22) | Group 8E [Dosed Moderna, Boost Pfizer] Age ≥56 yo (N=29) | Group 9E [Dosed Pfizer, Boost Pfizer] Age 18-55 yo (N=24) | Group 9E [Dosed Pfizer, Boost Pfizer] Age ≥56 yo (N=26) |
|---|---|---|---|---|---|---|
| **Day 1 Visit (Pre-boost)** | | | | | | |
| N (non-missing) | 31 | 22 | 22 | 29 | 24 | 26 |
| Median ($P_{25}$, $P_{75}$), BAU/mL | 78.47 (38.41-136.23) | 51.21 (40.56-134.69) | 778.71 (464.23-934.22) | 434.59 (251.25-740.84) | 258.87 (172.08-423.79) | 208.82 (116.16-315.68) |
| Minimum - Maximum, BAU/mL | 7.66-4279.92 | 5.02-543.77 | 355.94-1150.09 | 99.56-1881.23 | 81.30-1234.78 | 17.04-696.23 |
| Geometric Mean (95% CI), BAU/mL | 82.01 (52.32-128.53) | 66.29 (42.13-104.30) | 700.61 (588.17-834.55) | 435.11 (328.82-575.77) | 275.09 (202.03-374.56) | 184.54 (130.76-260.43) |
| **Day 15 Visit (14 days post-boost)** | | | | | | |
| N (non-missing) | 28 | 22 | 20 | 28 | 23 | 25 |
| Median ($P_{25}$, $P_{75}$), BAU/mL | 2996.63 (1872.74-4429.56) | 2383.21 (1163.60-3278.58) | 5659.91 (4429.81-7740.79) | 4461.69 (3326.99-6293.30) | 3647.23 (2130.66-7072.20) | 3172.78 (1997.14-5746.37) |
| Minimum - Maximum, BAU/mL | 735.92-24305.59 | 420.32-19884.77 | 1955.62-20018.49 | 1626.48-14296.01 | 844.68-19207.12 | 607.22-8348.38 |
| Geometric Mean (95% CI), BAU/mL | 2852.98 (2148.81-3787.90) | 2209.54 (1515.61-3221.20) | 5877.33 (4647.86-7432.04) | 4757.65 (3815.60-5932.28) | 3770.47 (2689.69-5285.53) | 3107.23 (2351.47-4105.89) |

[1] Values below the lower limit of detection (LLOD = 0.3076 BAU/mL) were assigned a value equal to LLOD/2. Values greater than the upper limit of quantification (ULOQ = 172226.25 BAU/mL) were taken as reported, or a ceiling value equivalent to the ULOQ is assigned if values are not provided.

[2] Bridging to the WHO standard Binding Antibody Units per milliliter (BAU/mL) was done using a conversion factor of 0.0090, specific to the IgG SARS-CoV-2 Spike antigen.

**Supplemental Table 16:** mRNA-1273 IgG Serum Binding Antibody Response to S-2P-WA-1 by 4-plex ECLIA V.2 by Group and Timepoint – Results are reported as Arbitrary Units (AU/mL)

| | Group 1E [Dosed Janssen, Boost Moderna] (N=53) | Group 2E [Dosed Moderna, Boost Moderna] (N=51) | Group 3E [Dosed Pfizer, Boost Moderna] (N=50) |
|---|---|---|---|
| **Day 1 Visit (Pre-boost)** | | | |
| N (non-missing) | 52 | 50 | 50 |
| Median ($P_{25}$, $P_{75}$), AU/mL | 5610.24 (3535.00-10525.50) | 72901.32 (46674.29-164346.92) | 44652.64 (23761.14-70988.74) |
| Minimum - Maximum, AU/mL | 1487.37-84008.26 | 24767.47-1821826.39 | 1842.83-479156.78 |
| Geometric Mean (95% CI), AU/mL | 6345.91 (4952.36-8131.59) | 95467.64 (74259.23-122733.15) | 39622.40 (29164.26-53830.77) |
| **Day 15 Visit (14 days post-boost)** | | | |
| N (non-missing) | 52 | 50 | 50 |
| Median ($P_{25}$, $P_{75}$), AU/mL | 386715.53 (200009.15-724150.81) | 725728.27 (514314.23-932062.91) | 690388.67 (323558.75-1358239.16) |
| Minimum - Maximum, AU/mL | 25996.95-2057203.30 | 273060.19-3835092.35 | 155203.93-2540410.44 |
| Geometric Mean (95% CI), AU/mL | 355901.12 (277716.35-456097.05) | 755529.37 (641308.04-890094.29) | 683890.33 (543935.26-859855.97) |
| N* (non-missing pre- and post-boost) | 52 | 50 | 50 |
| Participants with ≥ 2-fold rise[2], 95% CI | 100.0% (93.2%-100.0%) | 96.0% (86.3%-99.5%) | 98.0% (89.4%-99.9%) |
| Participants with ≥ 4-fold rise[2], 95% CI | 98.1% (89.7%-100.0%) | 84.0% (70.9%-92.8%) | 98.0% (89.4%-99.9%) |
| Geometric Mean Fold Rise[2], 95% CI | 56.08 (40.73-77.23) | 7.91 (6.20-10.10) | 17.26 (13.30-22.41) |
| **Day 29 Visit (28 days post-boost)** | | | |
| N (non-missing) | 43 | 48 | 43 |
| Median ($P_{25}$, $P_{75}$), AU/mL | 289599.14 (194590.99-507701.79) | 591842.50 (461638.68-881759.78) | 519642.38 (349452.21-992421.69) |
| Minimum - Maximum, AU/mL | 48639.88-1238627.37 | 219889.52-2264421.99 | 181648.46-2080669.29 |
| Geometric Mean (95% CI), AU/mL | 311457.03 (251477.47-385742.23) | 657486.68 (563377.05-767316.90) | 574470.43 (461876.84-714511.42) |
| N* (non-missing pre- and post-boost) | 42 | 47 | 43 |
| Participants with ≥ 2-fold rise[2], 95% CI | 100.0% (91.6%-100.0%) | 95.7% (85.5%-99.5%) | 97.7% (87.7%-99.9%) |
| Participants with ≥ 4-fold rise[2], 95% CI | 97.6% (87.4%-99.9%) | 74.5% (59.7%-86.1%) | 97.7% (87.7%-99.9%) |
| Geometric Mean Fold Rise[2], 95% CI | 48.64 (35.84-66.03) | 6.80 (5.28-8.75) | 15.36 (11.62-20.32) |

[1] Antibody values below the lower limit of quantification (LLOQ = 34.18 AU/mL) were assigned a value equal to LLOQ/2. Values greater than the upper limit of quantification (ULOQ = 19136250 AU/mL) are taken as reported, or a ceiling value equivalent to the ULOQ is assigned if values are not provided.
[2] Relative to pre-vaccination (Day 1 Visit) levels, among participants with no missing observations at both pre- and post-boost timepoints.

**Supplemental Table 17:** mRNA-1273 IgG Serum Binding Antibody Response to S-2P-WA-1 by 4-plex ECLIA V.2 by Group, Age Group and Timepoint – Results are reported as Arbitrary Units (AU/mL)

| | Group 1E [Dosed Janssen, Boost Moderna] Age 18-55 yo (N=21) | Group 1E [Dosed Janssen, Boost Moderna] Age ≥56 yo (N=32) | Group 2E [Dosed Moderna, Boost Moderna] Age 18-55 yo (N=26) | Group 2E [Dosed Moderna, Boost Moderna] Age ≥56 yo (N=25) | Group 3E [Dosed Pfizer, Boost Moderna] Age 18-55 yo (N=25) | Group 3E [Dosed Pfizer, Boost Moderna] Age ≥56 yo (N=25) |
|---|---|---|---|---|---|---|
| **Day 1 Visit (Pre-boost)** | | | | | | |
| N (non-missing) | 21 | 31 | 26 | 24 | 25 | 25 |
| Median ($P_{25}$, $P_{75}$), AU/mL | 5477.34 (3302.50-9506.49) | 6123.03 (3703.22-11686.38) | 68830.48 (46674.29-145873.55) | 90966.79 (45493.54-222983.76) | 49403.56 (30799.17-79859.16) | 40059.81 (12249.78-54109.19) |
| Minimum - Maximum, AU/mL | 1650.51-84008.26 | 1487.37-83909.44 | 30913.71-519258.33 | 24767.47-1821826.39 | 1842.83-479156.78 | 2926.42-285244.22 |
| Geometric Mean (95% CI), AU/mL | 6266.41 (4114.23-9544.40) | 6400.34 (4630.59-8846.45) | 86305.59 (63671.29-116986.10) | 106493.92 (69456.48-163281.47) | 51994.24 (33019.28-81873.43) | 30194.39 (19941.06-45719.79) |
| **Day 15 Visit (14 days post-boost)** | | | | | | |
| N (non-missing) | 21 | 31 | 26 | 24 | 25 | 25 |
| Median ($P_{25}$, $P_{75}$), AU/mL | 399616.59 (171871.27-740727.48) | 350983.59 (218193.67-707574.14) | 753136.29 (607224.29-860444.78) | 639186.20 (430599.61-1122924.73) | 1012031.77 (579994.32-1581849.69) | 445626.00 (282847.83-872724.17) |
| Minimum - Maximum, AU/mL | 25996.95-1605924.80 | 108937.75-2057203.30 | 379100.97-2775119.55 | 273060.19-3835092.35 | 217273.51-2540410.44 | 155203.93-2402374.88 |
| Geometric Mean (95% CI), AU/mL | 308650.10 (188194.26-506205.06) | 391955.27 (299479.87-512985.84) | 760110.29 (646779.63-893299.09) | 750597.85 (551171.17-1022181.77) | 910617.97 (684507.29-1211418.92) | 513613.83 (365771.72-721212.59) |
| N* (non-missing pre- and post-boost) | 21 | 31 | 26 | 24 | 25 | 25 |
| Participants with ≥ 2-fold rise[2], 95% CI | 100.0% (83.9%-100.0%) | 100.0% (88.8%-100.0%) | 100.0% (86.8%-100.0%) | 91.7% (73.0%-99.0%) | 96.0% (79.6%-99.9%) | 100.0% (86.3%-100.0%) |
| Participants with ≥ 4-fold rise[2], 95% CI | 95.2% (76.2%-99.9%) | 100.0% (88.8%-100.0%) | 88.5% (69.8%-97.6%) | 79.2% (57.8%-92.9%) | 96.0% (79.6%-99.9%) | 100.0% (86.3%-100.0%) |
| Geometric Mean Fold Rise[2], 95% CI | 49.25 (25.93-93.56) | 61.24 (43.23-86.75) | 8.81 (6.78-11.44) | 7.05 (4.53-10.96) | 17.51 (11.10-27.63) | 17.01 (12.70-22.79) |
| **Day 29 Visit (28 days post-boost)** | | | | | | |
| N (non-missing) | 18 | 25 | 25 | 23 | 21 | 22 |
| Median ($P_{25}$, $P_{75}$), AU/mL | 357091.84 (222186.72-507701.79) | 253659.78 (157990.57-498205.33) | 590822.49 (481714.08-828449.30) | 592862.52 (406611.93-943153.32) | 655152.79 (412668.33-1243265.24) | 446157.36 (278576.82-897724.45) |
| Minimum - Maximum, AU/mL | 140741.44-1021871.10 | 48639.88-1238627.37 | 272897.85-2030769.35 | 219889.52-2264421.99 | 181648.46-1792373.29 | 189479.72-2080669.29 |
| Geometric Mean (95% CI), AU/mL | 356058.61 (271330.90-467244.02) | 282845.52 (204854.13-390529.53) | 673920.72 (553103.73-821128.32) | 640077.81 (495003.05-827670.86) | 665030.24 (491550.73-899734.64) | 499555.43 (360313.72-692606.50) |

[1] Antibody values below the lower limit of quantification (LLOQ = 34.18 AU/mL) were assigned a value equal to LLOQ/2. Values greater than the upper limit of quantification (ULOQ = 19136250 AU/mL) are taken as reported, or a ceiling value equivalent to the ULOQ is assigned if values are not provided.
[2] Relative to pre-vaccination (Day 1 Visit) levels, among participants with no missing observations at both pre- and post-boost timepoints.

Supplemental Table 18: Ad26.COV2.S IgG Serum Binding Antibody Response to S-2P-WA-1 by 4-plex ECLIA V.2 by Group and Timepoint – Results are reported as Arbitrary Units (AU/mL)

| | Group 4E [Dosed Janssen, Boost Janssen] (N=50) | Group 5E [Dosed Moderna, Boost Janssen] (N=49) | Group 6E [Dosed Pfizer, Boost Janssen] (N=51) |
|---|---|---|---|
| **Day 1 Visit (Pre-boost)** | | | |
| N (non-missing) | 50 | 49 | 51 |
| Median ($P_{25}$, $P_{75}$), AU/mL | 8811.09 (3782.66-15369.22) | 68996.79 (48467.74-119625.82) | 35369.49 (21737.18-59749.44) |
| Minimum - Maximum, AU/mL | 17.10-455143.36 | 11158.39-397656.63 | 3296.39-268905.68 |
| Geometric Mean (95% CI), AU/mL | 7919.93 (5317.81-11795.31) | 70971.97 (57082.53-88241.03) | 35625.45 (27850.89-45570.26) |
| **Day 15 Visit (14 days post-boost)** | | | |
| N (non-missing) | 50 | 49 | 50 |
| Median ($P_{25}$, $P_{75}$), AU/mL | 38803.05 (23029.38-54058.43) | 281675.13 (172850.86-596746.78) | 189505.90 (123651.31-424105.29) |
| Minimum - Maximum, AU/mL | 138.97-420898.25 | 106593.09-2919661.56 | 45593.83-1113163.49 |
| Geometric Mean (95% CI), AU/mL | 36219.34 (26195.90-50078.09) | 336599.73 (270353.77-419078.23) | 211637.19 (166422.09-269136.75) |
| N* (non-missing pre- and post-boost) | 50 | 49 | 50 |
| Participants with ≥ 2-fold rise[2], 95% CI | 86.0% (73.3%-94.2%) | 83.7% (70.3%-92.7%) | 92.0% (80.8%-97.8%) |
| Participants with ≥ 4-fold rise[2], 95% CI | 48.0% (33.7%-62.6%) | 53.1% (38.3%-67.5%) | 60.0% (45.2%-73.6%) |
| Geometric Mean Fold Rise[2], 95% CI | 4.57 (3.66-5.72) | 4.74 (3.64-6.18) | 6.17 (4.68-8.14) |
| **Day 29 Visit (28 days post-boost)** | | | |
| N (non-missing) | 48 | 47 | 50 |
| Median ($P_{25}$, $P_{75}$), AU/mL | 40617.95 (28256.13-64027.24) | 427716.33 (256207.80-955324.82) | 313306.05 (172700.65-516224.68) |
| Minimum - Maximum, AU/mL | 5140.40-356944.36 | 85600.03-5594947.50 | 64574.67-1182366.62 |
| Geometric Mean (95% CI), AU/mL | 40960.22 (32305.60-51933.39) | 506677.18 (393797.99-651912.33) | 288848.70 (231767.09-359988.85) |
| N* (non-missing pre- and post-boost) | 48 | 47 | 50 |
| Participants with ≥ 2-fold rise[2], 95% CI | 95.8% (85.7%-99.5%) | 89.4% (76.9%-96.5%) | 94.0% (83.5%-98.7%) |
| Participants with ≥ 4-fold rise[2], 95% CI | 58.3% (43.2%-72.4%) | 70.2% (55.1%-82.7%) | 74.0% (59.7%-85.4%) |
| Geometric Mean Fold Rise[2], 95% CI | 5.30 (4.01-7.00) | 7.02 (5.15-9.58) | 7.92 (5.96-10.53) |

[1] Antibody values below the lower limit of quantification (LLOQ = 34.18 AU/mL) were assigned a value equal to LLOQ/2. Values greater than the upper limit of quantification (ULOQ = 19136250 AU/mL) are taken as reported, or a ceiling value equivalent to the ULOQ is assigned if values are not provided.
[2] Relative to pre-vaccination (Day 1 Visit) levels, among participants with no missing observations at both pre- and post-boost timepoints.

**Supplemental Table 19:** Ad26.COV2.S IgG Serum Binding Antibody Response to S-2P-WA-1 by 4-plex ECLIA V.2 by Group, Age Group and Timepoint – Results are reported as Arbitrary Units (AU/mL)

| | Group 4E [Dosed Janssen, Boost Janssen] Age 18-55 yo (N=26) | Group 4E [Dosed Janssen, Boost Janssen] Age ≥56 yo (N=24) | Group 5E [Dosed Moderna, Boost Janssen] Age 18-55 yo (N=24) | Group 5E [Dosed Moderna, Boost Janssen] Age ≥56 yo (N=25) | Group 6E [Dosed Pfizer, Boost Janssen] Age 18-55 yo (N=25) | Group 6E [Dosed Pfizer, Boost Janssen] Age ≥56 yo (N=26) |
|---|---|---|---|---|---|---|
| **Day 1 Visit (Pre-boost)** | | | | | | |
| N (non-missing) | 26 | 24 | 24 | 25 | 25 | 26 |
| Median ($P_{25}$, $P_{75}$), AU/mL | 8186.43 (3846.19-13029.68) | 9744.73 (3551.28-16314.85) | 77090.87 (46312.48-146748.70) | 68653.58 (52504.16-101808.41) | 35570.03 (26054.41-69362.25) | 28968.84 (15281.40-51448.67) |
| Minimum - Maximum, AU/mL | 1491.79-455143.36 | 17.10-344802.21 | 16402.24-397656.63 | 11158.39-169923.47 | 14080.03-244165.54 | 3296.39-268905.68 |
| Geometric Mean (95% CI), AU/mL | 8099.26 (5222.23-12561.32) | 7730.13 (3765.78-15867.83) | 82334.24 (58576.92-115726.93) | 61542.20 (46185.21-82005.53) | 44685.22 (32993.78-60519.56) | 28651.11 (19475.81-42149.02) |
| **Day 15 Visit (14 days post-boost)** | | | | | | |
| N (non-missing) | 26 | 24 | 24 | 25 | 24 | 26 |
| Median ($P_{25}$, $P_{75}$), AU/mL | 37390.68 (19814.27-53486.25) | 40007.96 (25360.05-69716.10) | 337598.64 (190674.14-831476.74) | 269308.58 (167416.14-453333.83) | 228452.05 (128421.40-465415.09) | 177386.01 (101481.46-311682.55) |
| Minimum - Maximum, AU/mL | 8395.34-352937.99 | 138.97-420898.25 | 107571.06-2919661.56 | 106593.09-1362837.84 | 45593.83-1004818.68 | 51023.12-1113163.49 |
| Geometric Mean (95% CI), AU/mL | 36237.08 (25760.99-50973.43) | 36200.13 (19944.01-65706.44) | 399151.24 (280709.22-567568.50) | 285792.61 (217243.47-375971.80) | 234831.24 (161873.35-340671.95) | 192265.90 (138220.44-267443.62) |
| N* (non-missing pre- and post-boost) | 26 | 24 | 24 | 25 | 24 | 26 |
| Participants with ≥ 2-fold rise[2], 95% CI | 88.5% (69.8%-97.6%) | 83.3% (62.6%-95.3%) | 91.7% (73.0%-99.0%) | 76.0% (54.9%-90.6%) | 95.8% (78.9%-99.9%) | 88.5% (69.8%-97.6%) |
| Participants with ≥ 4-fold rise[2], 95% CI | 50.0% (29.9%-70.1%) | 45.8% (25.6%-67.2%) | 45.8% (25.6%-67.2%) | 60.0% (38.7%-78.9%) | 54.2% (32.8%-74.4%) | 65.4% (44.3%-82.8%) |
| Geometric Mean Fold Rise[2], 95% CI | 4.47 (3.30-6.06) | 4.68 (3.29-6.67) | 4.85 (3.22-7.30) | 4.64 (3.21-6.73) | 5.64 (3.88-8.21) | 6.71 (4.38-10.28) |
| **Day 29 Visit (28 days post-boost)** | | | | | | |
| N (non-missing) | 24 | 24 | 23 | 24 | 25 | 25 |
| Median ($P_{25}$, $P_{75}$), AU/mL | 39010.08 (18783.62-61413.25) | 41391.67 (34028.23-69573.06) | 467120.98 (298555.23-1198457.51) | 414788.84 (212681.58-639899.76) | 300626.63 (175097.49-540260.22) | 316552.96 (159701.72-447434.66) |
| Minimum - Maximum, AU/mL | 10367.81-254247.22 | 5140.40-356944.36 | 172135.51-5594947.50 | 85600.03-2918848.17 | 69436.54-1182366.62 | 64574.67-1042839.17 |
| Geometric Mean (95% CI), AU/mL | 38015.42 (27019.08-53487.11) | 44133.13 (30997.92-62834.32) | 595891.61 (408963.86-868259.62) | 433740.78 (304850.21-617126.24) | 303855.41 (221907.70-416065.38) | 274583.13 (197345.75-382049.76) |
| N* (non-missing pre- and post-boost) | 24 | 24 | 23 | 24 | 25 | 25 |
| Participants with ≥ 2-fold rise[2], 95% CI | 95.8% (78.9%-99.9%) | 95.8% (78.9%-99.9%) | 95.7% (78.1%-99.9%) | 83.3% (62.6%-95.3%) | 92.0% (74.0%-99.0%) | 96.0% (79.6%-99.9%) |

[1] Antibody values below the lower limit of quantification (LLOQ = 34.18 AU/mL) were assigned a value equal to LLOQ/2. Values greater than the upper limit of quantification (ULOQ = 19136250 AU/mL) are taken as reported, or a ceiling value equivalent to the ULOQ is assigned if values are not provided.
[2] Relative to pre-vaccination (Day 1 Visit) levels, among participants with no missing observations at both pre- and post-boost timepoints.

**Supplemental Table 20:** BNT162b2 IgG Serum Binding Antibody Response to S-2P-WA-1 by 4-plex ECLIA V.2 by Group and Timepoint – Results are reported as Arbitrary Units (AU/mL)

| | Group 7E [Dosed Janssen, Boost Pfizer] (N=53) | Group 8E [Dosed Moderna, Boost Pfizer] (N=51) | Group 9E [Dosed Pfizer, Boost Pfizer] (N=50) |
|---|---|---|---|
| **Day 1 Visit (Pre-boost)** | | | |
| N (non-missing) | 53 | 51 | 50 |
| Median ($P_{25}$, $P_{75}$), AU/mL | 8597.27 (4507.07-14965.97) | 70692.45 (39548.52-93021.68) | 25169.72 (14510.02-40758.20) |
| Minimum - Maximum, AU/mL | 558.14-475546.54 | 11062.48-209025.58 | 1893.55-137197.67 |
| Geometric Mean (95% CI), AU/mL | 8341.58 (6092.72-11420.50) | 59374.50 (49418.79-71335.84) | 24835.30 (19706.29-31299.25) |
| **Day 15 Visit (14 days post-boost)** | | | |
| N (non-missing) | 50 | 48 | 48 |
| Median ($P_{25}$, $P_{75}$), AU/mL | 286648.40 (190055.88-412847.34) | 579116.52 (408150.98-746270.31) | 388533.30 (229321.83-683417.73) |
| Minimum - Maximum, AU/mL | 46702.05-2700621.15 | 180719.99-2224277.09 | 67468.64-2134124.23 |
| Geometric Mean (95% CI), AU/mL | 283281.28 (226454.27-354368.60) | 577290.94 (492566.63-676588.32) | 378783.46 (306737.37-467751.64) |
| N* (non-missing pre- and post-boost) | 50 | 48 | 48 |
| Participants with ≥ 2-fold rise[2], 95% CI | 98.0% (89.4%-99.9%) | 100.0% (92.6%-100.0%) | 100.0% (92.6%-100.0%) |
| Participants with ≥ 4-fold rise[2], 95% CI | 98.0% (89.4%-99.9%) | 93.8% (82.8%-98.7%) | 95.8% (85.7%-99.5%) |
| Geometric Mean Fold Rise[2], 95% CI | 32.84 (24.63-43.78) | 9.71 (8.01-11.77) | 14.94 (11.82-18.88) |

[1] Antibody values below the lower limit of quantification (LLOQ = 34.18 AU/mL) were assigned a value equal to LLOQ/2. Values greater than the upper limit of quantification (ULOQ = 19136250 AU/mL) are taken as reported, or a ceiling value equivalent to the ULOQ is assigned if values are not provided.
[2] Relative to pre-vaccination (Day 1 Visit) levels, among participants with no missing observations at both pre- and post-boost timepoints.

**Supplemental Table 21:** BNT162b2 IgG Serum Binding Antibody Response to S-2P-WA-1 by 4-plex ECLIA V.2 by Group, Age Group and Timepoint – Results are reported as Arbitrary Units (AU/mL)

| | Group 7E [Dosed Janssen, Boost Pfizer] Age 18-55 yo (N=31) | Group 7E [Dosed Janssen, Boost Pfizer] Age ≥56 yo (N=22) | Group 8E [Dosed Moderna, Boost Pfizer] Age 18-55 yo (N=22) | Group 8E [Dosed Moderna, Boost Pfizer] Age ≥56 yo (N=29) | Group 9E [Dosed Pfizer, Boost Pfizer] Age 18-55 yo (N=24) | Group 9E [Dosed Pfizer, Boost Pfizer] Age ≥56 yo (N=26) |
|---|---|---|---|---|---|---|
| **Day 1 Visit (Pre-boost)** | | | | | | |
| N (non-missing) | 31 | 22 | 22 | 29 | 24 | 26 |
| Median ($P_{25}$, $P_{75}$), AU/mL | 8718.61 (4267.36-15136.52) | 5690.34 (4507.07-14965.97) | 86523.77 (51581.66-103801.95) | 48288.14 (27916.38-82315.64) | 28763.36 (19120.18-47087.62) | 23201.85 (12906.28-35075.66) |
| Minimum - Maximum, AU/mL | 851.12-475546.54 | 558.14-60418.45 | 39548.52-127788.15 | 11062.48-209025.58 | 9033.17-137197.67 | 1893.55-77359.37 |
| Geometric Mean (95% CI), AU/mL | 9111.80 (5813.67-14280.98) | 7365.48 (4681.23-11588.88) | 77845.87 (65352.14-92728.08) | 48345.85 (36535.10-63974.67) | 30565.21 (22447.67-41618.22) | 20504.37 (14529.04-28937.18) |
| **Day 15 Visit (14 days post-boost)** | | | | | | |
| N (non-missing) | 28 | 22 | 20 | 28 | 23 | 25 |
| Median ($P_{25}$, $P_{75}$), AU/mL | 332959.15 (208081.97-492173.05) | 264801.44 (129288.51-364286.34) | 628878.61 (492201.17-860087.60) | 495742.83 (369665.08-699255.14) | 405247.57 (236739.65-785799.47) | 352531.56 (221904.01-638485.94) |
| Minimum - Maximum, AU/mL | 81769.30-2700621.15 | 46702.05-2209418.77 | 217291.65-2224277.09 | 180719.99-1588445.93 | 93852.88-2134124.23 | 67468.64-927597.84 |
| Geometric Mean (95% CI), AU/mL | 316997.90 (238757.18-420878.11) | 245504.67 (168400.65-357911.58) | 653037.14 (516428.42-825782.42) | 528627.36 (423955.38-659142.22) | 418940.71 (298853.96-587281.22) | 345247.37 (261273.91-456209.90) |
| N* (non-missing pre- and post-boost) | 28 | 22 | 20 | 28 | 23 | 25 |
| Participants with ≥ 2-fold rise[2], 95% CI | 96.4% (81.7%-99.9%) | 100.0% (84.6%-100.0%) | 100.0% (83.2%-100.0%) | 100.0% (87.7%-100.0%) | 100.0% (85.2%-100.0%) | 100.0% (86.3%-100.0%) |
| Participants with ≥ 4-fold rise[2], 95% CI | 96.4% (81.7%-99.9%) | 100.0% (84.6%-100.0%) | 90.0% (68.3%-98.8%) | 96.4% (81.7%-99.9%) | 91.3% (72.0%-98.9%) | 100.0% (86.3%-100.0%) |
| Geometric Mean Fold Rise[2], 95% CI | 32.46 (21.29-49.50) | 33.33 (22.06-50.36) | 7.87 (6.07-10.20) | 11.28 (8.60-14.80) | 13.41 (9.58-18.77) | 16.49 (11.68-23.29) |

[1] Antibody values below the lower limit of quantification (LLOQ = 34.18 AU/mL) were assigned a value equal to LLOQ/2. Values greater than the upper limit of quantification (ULOQ = 19136250 AU/mL) are taken as reported, or a ceiling value equivalent to the ULOQ is assigned if values are not provided.
[2] Relative to pre-vaccination (Day 1 Visit) levels, among participants with no missing observations at both pre- and post-boost timepoints.

**Supplemental Table 22:** Variants of Concern: mRNA-1273 IgG Serum Binding Antibody Response to S-2P-WA-1 (control) by FFP 10-plex ECLIA by Group and Timepoint – Results are reported as Area Under Curve (AUC)

| | Group 4E [Dosed Janssen, Boost Janssen] (N=50) | Group 5E [Dosed Moderna, Boost Janssen] (N=49) | Group 6E [Dosed Pfizer, Boost Janssen] (N=51) |
|---|---|---|---|
| **Day 1 Visit (Pre-boost)** | | | |
| N (non-missing) | 50 | 49 | 51 |
| Median ($P_{25}$, $P_{75}$), AUC | 3031.50 (1455.00-4965.00) | 19057.00 (14608.00-33786.00) | 10495.00 (6865.00-17920.00) |
| Minimum - Maximum, AUC | 53.72-50700.00 | 3173.00-47335.00 | 1275.00-44083.00 |
| Geometric Mean (95% CI), AUC | 2706.54 (1987.71-3685.33) | 19727.12 (16571.28-23483.96) | 10813.77 (8790.20-13303.18) |
| **Day 15 Visit (14 days post-boost)** | | | |
| N (non-missing) | 50 | 49 | 50 |
| Median ($P_{25}$, $P_{75}$), AUC | 10998.50 (6611.00-14542.00) | 45849.00 (37372.00-59381.00) | 38346.00 (32645.00-50536.00) |
| Minimum - Maximum, AUC | 77.03-48198.00 | 23983.00-68352.00 | 12782.00-62657.00 |
| Geometric Mean (95% CI), AUC | 9822.92 (7485.72-12889.85) | 45825.57 (42581.21-49317.13) | 36136.92 (32136.69-40635.07) |
| N* (non-missing pre- and post-boost) | 50 | 49 | 50 |
| Participants with ≥ 2-fold rise[1], 95% CI | 82.0% (68.6%-91.4%) | 55.1% (40.2%-69.3%) | 72.0% (57.5%-83.8%) |
| Participants with ≥ 4-fold rise[1], 95% CI | 42.0% (28.2%-56.8%) | 16.3% (7.3%-29.7%) | 42.0% (28.2%-56.8%) |
| Geometric Mean Fold Rise[1], 95% CI | 3.63 (2.96-4.45) | 2.32 (1.96-2.75) | 3.43 (2.79-4.22) |
| **Day 29 Visit (28 days post-boost)** | | | |
| N (non-missing) | 48 | 47 | 50 |
| Median ($P_{25}$, $P_{75}$), AUC | 10949.50 (7134.00-16366.00) | 52868.00 (42994.00-62659.00) | 45376.00 (35829.00-52548.00) |
| Minimum - Maximum, AUC | 1753.00-50071.00 | 25284.00-70644.00 | 17572.00-65009.00 |
| Geometric Mean (95% CI), AUC | 10536.86 (8586.26-12930.59) | 51025.30 (47728.29-54550.05) | 41488.05 (37555.26-45832.69) |
| N* (non-missing pre- and post-boost) | 48 | 47 | 50 |
| Participants with ≥ 2-fold rise[1], 95% CI | 89.6% (77.3%-96.5%) | 61.7% (46.4%-75.5%) | 78.0% (64.0%-88.5%) |
| Participants with ≥ 4-fold rise[1], 95% CI | 43.8% (29.5%-58.8%) | 19.1% (9.1%-33.3%) | 46.0% (31.8%-60.7%) |
| Geometric Mean Fold Rise[1], 95% CI | 3.97 (3.20-4.94) | 2.55 (2.12-3.06) | 3.77 (3.05-4.67) |

[1] Relative to pre-vaccination (Day 1 Visit) levels, among participants with no missing observations at both pre- and post-boost timepoints.

**Supplemental Table 23:** Variants of Concern: mRNA-1273 IgG Serum Binding Antibody Response to S-2P-WA-1 (control) by FFP 10-plex ECLIA by Group, Age Group, and Timepoint – Results are reported as Area Under Curve (AUC)

| | Group 1E [Dosed Janssen, Boost Moderna] Age 18-55 yo (N=21) | Group 1E [Dosed Janssen, Boost Moderna] Age ≥56 yo (N=32) | Group 2E [Dosed Moderna, Boost Moderna] Age 18-55 yo (N=26) | Group 2E [Dosed Moderna, Boost Moderna] Age ≥56 yo (N=25) | Group 3E [Dosed Pfizer, Boost Moderna] Age 18-55 yo (N=25) | Group 3E [Dosed Pfizer, Boost Moderna] Age ≥56 yo (N=25) |
|---|---|---|---|---|---|---|
| **Day 1 Visit (Pre-boost)** | | | | | | |
| N (non-missing) | 21 | 31 | 26 | 24 | 25 | 25 |
| Median ($P_{25}$, $P_{75}$), AUC | 2342.00 (1435.00-4981.00) | 2413.00 (1462.00-3422.00) | 23736.50 (17728.00-38221.00) | 25872.50 (14919.50-39560.50) | 15371.00 (11212.00-22136.00) | 13850.00 (6372.00-17394.00) |
| Minimum - Maximum, AUC | 799.80-27530.00 | 666.40-23981.00 | 11569.00-61246.00 | 7284.00-55018.00 | 803.40-50653.00 | 1783.00-52077.00 |
| Geometric Mean (95% CI), AUC | 2729.68 (1836.36-4057.59) | 2547.91 (1893.75-3428.02) | 24582.44 (20215.31-29892.99) | 23808.26 (18843.60-30080.93) | 14551.90 (10404.34-20352.83) | 11221.59 (7966.67-15806.37) |
| **Day 15 Visit (14 days post-boost)** | | | | | | |
| N (non-missing) | 21 | 31 | 26 | 24 | 25 | 25 |
| Median ($P_{25}$, $P_{75}$), AUC | 51935.00 (45830.00-61228.00) | 49734.00 (39452.00-62011.00) | 61870.50 (59515.00-63656.00) | 62151.00 (55693.50-64466.50) | 62664.00 (54879.00-64951.00) | 55935.00 (48192.00-64121.00) |
| Minimum - Maximum, AUC | 36970.00-68253.00 | 27685.00-66048.00 | 43701.00-67600.00 | 41495.00-68226.00 | 48735.00-67277.00 | 35262.00-71101.00 |
| Geometric Mean (95% CI), AUC | 52153.68 (48030.00-56631.39) | 48224.76 (44132.60-52696.35) | 61109.71 (59107.84-63179.39) | 58587.47 (55432.85-61921.61) | 59698.34 (57036.09-62484.85) | 53885.68 (49632.83-58502.94) |
| N* (non-missing pre- and post-boost) | 21 | 31 | 26 | 24 | 25 | 25 |
| Participants with ≥ 2-fold rise[2], 95% CI | 100.0% (83.9%-100.0%) | 100.0% (88.8%-100.0%) | 65.4% (44.3%-82.8%) | 58.3% (36.6%-77.9%) | 88.0% (68.8%-97.5%) | 92.0% (74.0%-99.0%) |
| Participants with ≥ 4-fold rise[2], 95% CI | 95.2% (76.2%-99.9%) | 96.8% (83.3%-99.9%) | 11.5% (2.4%-30.2%) | 29.2% (12.6%-51.1%) | 44.0% (24.4%-65.1%) | 56.0% (34.9%-75.6%) |
| Geometric Mean Fold Rise[2], 95% CI | 19.11 (12.93-28.23) | 18.93 (14.27-25.11) | 2.49 (2.07-2.99) | 2.46 (2.00-3.02) | 4.10 (2.95-5.70) | 4.80 (3.58-6.44) |
| **Day 29 Visit (28 days post-boost)** | | | | | | |
| N (non-missing) | 18 | 25 | 25 | 23 | 21 | 22 |
| Median ($P_{25}$, $P_{75}$), AUC | 43333.50 (38791.00-52193.00) | 41573.00 (32106.00-51180.00) | 56538.00 (51300.00-59395.00) | 53933.00 (47716.00-60948.00) | 57595.00 (48785.00-60834.00) | 46501.50 (38581.00-59845.00) |
| Minimum - Maximum, AUC | 29471.00-62586.00 | 12350.00-64775.00 | 41341.00-64312.00 | 36823.00-65951.00 | 38787.00-62516.00 | 31533.00-65490.00 |
| Geometric Mean (95% CI), AUC | 44534.67 (40484.43-48990.11) | 39335.24 (34033.86-45462.39) | 55229.50 (52815.82-57753.48) | 53325.68 (50025.94-56843.07) | 53784.73 (50396.66-57400.58) | 47790.50 (43131.24-52953.08) |
| N* (non-missing pre- and post-boost) | 18 | 24 | 25 | 22 | 21 | 22 |
| Participants with ≥ 2-fold rise[2], 95% CI | 94.4% (72.7%-99.9%) | 100.0% (85.8%-100.0%) | 56.0% (34.9%-75.6%) | 50.0% (28.2%-71.8%) | 90.5% (69.6%-98.8%) | 86.4% (65.1%-97.1%) |

[1] Relative to pre-vaccination (Day 1 Visit) levels, among participants with no missing observations at both pre- and post-boost timepoints.

**Supplemental Table 24:** Variants of Concern: Ad26.COV2.S IgG Serum Binding Antibody Response to S-2P-WA-1 (control) by FFP 10-plex ECLIA by Group and Timepoint – Results are reported as Area Under Curve (AUC)

| | Group 4E [Dosed Janssen, Boost Janssen] (N=50) | Group 5E [Dosed Moderna, Boost Janssen] (N=49) | Group 6E [Dosed Pfizer, Boost Janssen] (N=51) |
|---|---|---|---|
| **Day 1 Visit (Pre-boost)** | | | |
| N (non-missing) | 50 | 49 | 51 |
| Median ($P_{25}$, $P_{75}$), AUC | 3031.50 (1455.00-4965.00) | 19057.00 (14608.00-33786.00) | 10495.00 (6865.00-17920.00) |
| Minimum - Maximum, AUC | 53.72-50700.00 | 3173.00-47335.00 | 1275.00-44083.00 |
| Geometric Mean (95% CI), AUC | 2706.54 (1987.71-3685.33) | 19727.12 (16571.28-23483.96) | 10813.77 (8790.20-13303.18) |
| **Day 15 Visit (14 days post-boost)** | | | |
| N (non-missing) | 50 | 49 | 50 |
| Median ($P_{25}$, $P_{75}$), AUC | 10998.50 (6611.00-14542.00) | 45849.00 (37372.00-59381.00) | 38346.00 (32645.00-50536.00) |
| Minimum - Maximum, AUC | 77.03-48198.00 | 23983.00-68352.00 | 12782.00-62657.00 |
| Geometric Mean (95% CI), AUC | 9822.92 (7485.72-12889.85) | 45825.57 (42581.21-49317.13) | 36136.92 (32136.69-40635.07) |
| N* (non-missing pre- and post-boost) | 50 | 49 | 50 |
| Participants with ≥ 2-fold rise[1], 95% CI | 82.0% (68.6%-91.4%) | 55.1% (40.2%-69.3%) | 72.0% (57.5%-83.8%) |
| Participants with ≥ 4-fold rise[1], 95% CI | 42.0% (28.2%-56.8%) | 16.3% (7.3%-29.7%) | 42.0% (28.2%-56.8%) |
| Geometric Mean Fold Rise[1], 95% CI | 3.63 (2.96-4.45) | 2.32 (1.96-2.75) | 3.43 (2.79-4.22) |
| **Day 29 Visit (28 days post-boost)** | | | |
| N (non-missing) | 48 | 47 | 50 |
| Median ($P_{25}$, $P_{75}$), AUC | 10949.50 (7134.00-16366.00) | 52868.00 (42994.00-62659.00) | 45376.00 (35829.00-52548.00) |
| Minimum - Maximum, AUC | 1753.00-50071.00 | 25284.00-70644.00 | 17572.00-65009.00 |
| Geometric Mean (95% CI), AUC | 10536.86 (8586.26-12930.59) | 51025.30 (47728.29-54550.05) | 41488.05 (37555.26-45832.69) |
| N* (non-missing pre- and post-boost) | 48 | 47 | 50 |
| Participants with ≥ 2-fold rise[1], 95% CI | 89.6% (77.3%-96.5%) | 61.7% (46.4%-75.5%) | 78.0% (64.0%-88.5%) |
| Participants with ≥ 4-fold rise[1], 95% CI | 43.8% (29.5%-58.8%) | 19.1% (9.1%-33.3%) | 46.0% (31.8%-60.7%) |
| Geometric Mean Fold Rise[1], 95% CI | 3.97 (3.20-4.94) | 2.55 (2.12-3.06) | 3.77 (3.05-4.67) |

[1] Relative to pre-vaccination (Day 1 Visit) levels, among participants with no missing observations at both pre- and post-boost timepoints.

**Supplemental Table 25:** Variants of Concern: Ad26.COV2.S IgG Serum Binding Antibody Response to S-2P-WA-1 (control) by FFP 10-plex ECLIA by Group, Age Group, and Timepoint – Results are reported as Area Under Curve (AUC)

| | Group 4E [Dosed Janssen, Boost Janssen] Age 18-55 yo (N=26) | Group 4E [Dosed Janssen, Boost Janssen] Age ≥56 yo (N=24) | Group 5E [Dosed Moderna, Boost Janssen] Age 18-55 yo (N=24) | Group 5E [Dosed Moderna, Boost Janssen] Age ≥56 yo (N=25) | Group 6E [Dosed Pfizer, Boost Janssen] Age 18-55 yo (N=25) | Group 6E [Dosed Pfizer, Boost Janssen] Age ≥56 yo (N=26) |
|---|---|---|---|---|---|---|
| **Day 1 Visit (Pre-boost)** | | | | | | |
| N (non-missing) | 26 | 24 | 24 | 25 | 25 | 26 |
| Median ($P_{25}$, $P_{75}$), AUC | 2935.50 (1546.00-4615.00) | 3197.00 (1350.50-5669.50) | 20270.00 (15106.00-36667.50) | 18843.00 (14298.00-28695.00) | 12333.00 (9136.00-19188.00) | 9131.50 (4894.00-16277.00) |
| Minimum - Maximum, AUC | 437.80-50700.00 | 53.72-47732.00 | 5910.00-47335.00 | 3173.00-40665.00 | 4217.00-40327.00 | 1275.00-44083.00 |
| Geometric Mean (95% CI), AUC | 2700.05 (1856.41-3927.06) | 2713.60 (1596.43-4612.55) | 22335.74 (17537.37-28446.98) | 17509.94 (13521.90-22674.17) | 13392.14 (10449.69-17163.13) | 8803.93 (6358.95-12188.99) |
| **Day 15 Visit (14 days post-boost)** | | | | | | |
| N (non-missing) | 26 | 24 | 24 | 25 | 24 | 26 |
| Median ($P_{25}$, $P_{75}$), AUC | 10654.00 (5301.00-14471.00) | 11174.00 (7490.00-18668.00) | 50139.50 (37916.00-62454.00) | 42749.00 (37372.00-49570.00) | 40456.50 (32666.00-51659.50) | 37308.50 (25454.00-45638.00) |
| Minimum - Maximum, AUC | 2758.00-47107.00 | 77.03-48198.00 | 28672.00-68352.00 | 23983.00-67326.00 | 12782.00-62657.00 | 13501.00-62236.00 |
| Geometric Mean (95% CI), AUC | 9809.76 (7425.83-12959.00) | 9837.20 (5935.98-16302.35) | 48843.60 (43773.91-54500.45) | 43103.86 (39018.66-47616.76) | 37923.14 (31768.77-45269.75) | 34562.86 (29290.79-40783.86) |
| N* (non-missing pre- and post-boost) | 26 | 24 | 24 | 25 | 24 | 26 |
| Participants with ≥ 2-fold rise[1], 95% CI | 84.6% (65.1%-95.6%) | 79.2% (57.8%-92.9%) | 45.8% (25.6%-67.2%) | 64.0% (42.5%-82.0%) | 66.7% (44.7%-84.4%) | 76.9% (56.4%-91.0%) |
| Participants with ≥ 4-fold rise[1], 95% CI | 42.3% (23.4%-63.1%) | 41.7% (22.1%-63.4%) | 12.5% (2.7%-32.4%) | 20.0% (6.8%-40.7%) | 29.2% (12.6%-51.1%) | 53.8% (33.4%-73.4%) |
| Geometric Mean Fold Rise[1], 95% CI | 3.63 (2.75-4.81) | 3.63 (2.63-5.00) | 2.19 (1.73-2.76) | 2.46 (1.90-3.19) | 2.96 (2.34-3.76) | 3.93 (2.79-5.52) |
| **Day 29 Visit (28 days post-boost)** | | | | | | |
| N (non-missing) | 24 | 24 | 23 | 24 | 25 | 25 |
| Median ($P_{25}$, $P_{75}$), AUC | 10175.50 (5466.50-15171.50) | 11114.00 (8449.50-17268.50) | 54196.00 (46617.00-63975.00) | 51993.50 (41424.00-58574.50) | 45258.00 (36321.00-52548.00) | 45494.00 (35829.00-50275.00) |
| Minimum - Maximum, AUC | 2750.00-44680.00 | 1753.00-50071.00 | 35513.00-69381.00 | 25284.00-70644.00 | 18835.00-64456.00 | 17572.00-65009.00 |
| Geometric Mean (95% CI), AUC | 9732.68 (7229.91-13101.82) | 11407.49 (8445.28-15408.70) | 53486.66 (48992.00-58393.66) | 48772.85 (43989.43-54076.42) | 42255.57 (36903.93-48383.28) | 40734.47 (34858.93-47600.35) |
| N* (non-missing pre- and post-boost) | 24 | 24 | 23 | 24 | 25 | 25 |
| Participants with ≥ 2-fold rise[1], 95% CI | 87.5% (67.6%-97.3%) | 91.7% (73.0%-99.0%) | 52.2% (30.6%-73.2%) | 70.8% (48.9%-87.4%) | 76.0% (54.9%-90.6%) | 80.0% (59.3%-93.2%) |
| Participants with ≥ 4-fold rise[1], 95% CI | 41.7% (22.1%-63.4%) | 45.8% (25.6%-67.2%) | 17.4% (5.0%-38.8%) | 20.8% (7.1%-42.2%) | 36.0% (18.0%-57.5%) | 56.0% (34.9%-75.6%) |
| Geometric Mean Fold Rise[1], 95% CI | 3.76 (2.84-4.97) | 4.20 (2.95-5.99) | 2.30 (1.81-2.93) | 2.81 (2.10-3.75) | 3.16 (2.49-4.00) | 4.51 (3.15-6.47) |

[1] Relative to pre-vaccination (Day 1 Visit) levels, among participants with no missing observations at both pre- and post-boost timepoints.

**Supplemental Table 26:** Variants of Concern: BNT162b2 IgG Serum Binding Antibody Response to S-2P-WA-1 (control) by FFP 10-plex ECLIA by Group and Timepoint – Results are reported as Area Under Curve (AUC)

| | Group 7E [Dosed Janssen, Boost Pfizer] (N=53) | Group 8E [Dosed Moderna, Boost Pfizer] (N=51) | Group 9E [Dosed Pfizer, Boost Pfizer] (N=50) |
|---|---|---|---|
| **Day 1 Visit (Pre-boost)** | | | |
| N (non-missing) | 53 | 51 | 50 |
| Median ($P_{25}$, $P_{75}$), AUC | 3107.00 (2030.00-5702.00) | 21077.00 (14770.00-30985.00) | 10091.00 (6887.00-15435.00) |
| Minimum - Maximum, AUC | 348.80-51711.00 | 5532.00-43467.00 | 990.60-41508.00 |
| Geometric Mean (95% CI), AUC | 3232.04 (2455.97-4253.35) | 20133.71 (17513.09-23146.47) | 9854.17 (8032.98-12088.24) |
| **Day 15 Visit (14 days post-boost)** | | | |
| N (non-missing) | 50 | 48 | 48 |
| Median ($P_{25}$, $P_{75}$), AUC | 44122.50 (40561.00-49190.00) | 57606.50 (51274.00-61981.00) | 50342.50 (40357.50-59798.50) |
| Minimum - Maximum, AUC | 16445.00-66728.00 | 40402.00-69494.00 | 18919.00-66342.00 |
| Geometric Mean (95% CI), AUC | 43001.80 (39750.42-46519.13) | 55687.02 (53386.44-58086.75) | 47637.59 (44293.24-51234.46) |
| N* (non-missing pre- and post-boost) | 50 | 48 | 48 |
| Participants with ≥ 2-fold rise[1], 95% CI | 98.0% (89.4%-99.9%) | 66.7% (51.6%-79.6%) | 91.7% (80.0%-97.7%) |
| Participants with ≥ 4-fold rise[1], 95% CI | 90.0% (78.2%-96.7%) | 22.9% (12.0%-37.3%) | 64.6% (49.5%-77.8%) |
| Geometric Mean Fold Rise[1], 95% CI | 12.87 (10.06-16.45) | 2.77 (2.42-3.18) | 4.72 (3.90-5.73) |

[1] Relative to pre-vaccination (Day 1 Visit) levels, among participants with no missing observations at both pre- and post-boost timepoints.

**Supplemental Table 27:** Variants of Concern: BNT162b2 IgG Serum Binding Antibody Response to S-2P-WA-1 (control) by FFP 10-plex ECLIA by Group, Age Group, and Timepoint – Results are reported as Area Under Curve (AUC)

| | Group 7E [Dosed Janssen, Boost Pfizer] Age 18-55 yo (N=31) | Group 7E [Dosed Janssen, Boost Pfizer] Age ≥56 yo (N=22) | Group 8E [Dosed Moderna, Boost Pfizer] Age 18-55 yo (N=22) | Group 8E [Dosed Moderna, Boost Pfizer] Age ≥56 yo (N=29) | Group 9E [Dosed Pfizer, Boost Pfizer] Age 18-55 yo (N=24) | Group 9E [Dosed Pfizer, Boost Pfizer] Age ≥56 yo (N=26) |
|---|---|---|---|---|---|---|
| **Day 1 Visit (Pre-boost)** | | | | | | |
| N (non-missing) | 31 | 22 | 22 | 29 | 24 | 26 |
| Median ($P_{25}$, $P_{75}$), AUC | 3364.00 (2139.00-7309.00) | 2674.00 (1799.00-5479.00) | 27875.00 (19731.00-32738.00) | 16368.00 (11364.00-24025.00) | 11994.50 (7080.00-19453.00) | 8728.50 (5374.00-13798.00) |
| Minimum - Maximum, AUC | 360.00-51711.00 | 348.80-16893.00 | 14954.00-39432.00 | 5532.00-43467.00 | 4898.00-41508.00 | 990.60-21392.00 |
| Geometric Mean (95% CI), AUC | 3482.28 (2341.83-5178.14) | 2909.65 (1973.70-4289.44) | 25688.25 (22616.25-29177.51) | 16736.07 (13611.95-20577.22) | 12421.00 (9535.25-16180.08) | 7958.24 (5892.45-10748.26) |
| **Day 15 Visit (14 days post-boost)** | | | | | | |
| N (non-missing) | 28 | 22 | 20 | 28 | 23 | 25 |
| Median ($P_{25}$, $P_{75}$), AUC | 45064.50 (40639.00-52772.50) | 43270.00 (40440.00-47859.00) | 59473.50 (55949.00-61541.50) | 55258.50 (47931.00-62095.50) | 51234.00 (40753.00-61623.00) | 47369.00 (39174.00-54527.00) |
| Minimum - Maximum, AUC | 19479.00-65164.00 | 16445.00-66728.00 | 42061.00-67523.00 | 40402.00-69494.00 | 30854.00-66342.00 | 18919.00-63223.00 |
| Geometric Mean (95% CI), AUC | 44644.04 (40652.27-49027.77) | 40998.76 (35652.78-47146.33) | 57827.71 (54793.92-61029.46) | 54206.65 (50944.95-57677.18) | 49607.94 (45084.73-54584.96) | 45894.06 (40981.30-51395.75) |
| N* (non-missing pre- and post-boost) | 28 | 22 | 20 | 28 | 23 | 25 |
| Participants with ≥ 2-fold rise[1], 95% CI | 96.4% (81.7%-99.9%) | 100.0% (84.6%-100.0%) | 50.0% (27.2%-72.8%) | 78.6% (59.0%-91.7%) | 87.0% (66.4%-97.2%) | 96.0% (79.6%-99.9%) |
| Participants with ≥ 4-fold rise[1], 95% CI | 85.7% (67.3%-96.0%) | 95.5% (77.2%-99.9%) | 5.0% (0.1%-24.9%) | 35.7% (18.6%-55.9%) | 56.5% (34.5%-76.8%) | 72.0% (50.6%-87.9%) |
| Geometric Mean Fold Rise[1], 95% CI | 11.98 (8.37-17.16) | 14.09 (9.93-20.00) | 2.18 (1.92-2.48) | 3.29 (2.71-4.01) | 3.92 (3.05-5.04) | 5.60 (4.20-7.48) |

[1] Relative to pre-vaccination (Day 1 Visit) levels, among participants with no missing observations at both pre- and post-boost timepoints.

**Supplemental Table 28:** Variants of Concern: mRNA-1273 IgG Serum Binding Antibody Response to S-2P-B.1.617.2 by FFP 10-plex ECLIA by Group, and Timepoint – Results are reported as Area Under Curve (AUC)

| | Group 1E [Dosed Janssen, Boost Moderna] (N=53) | Group 2E [Dosed Moderna, Boost Moderna] (N=51) | Group 3E [Dosed Pfizer, Boost Moderna] (N=50) |
|---|---|---|---|
| **Day 1 Visit (Pre-boost)** | | | |
| N (non-missing) | 52 | 50 | 50 |
| Median ($P_{25}$, $P_{75}$), AUC | 1228.00 (757.15-2420.00) | 14338.50 (9436.00-25441.00) | 8672.50 (5043.00-12092.00) |
| Minimum - Maximum, AUC | 379.30-14547.00 | 4217.00-48856.00 | 551.80-39146.00 |
| Geometric Mean (95% CI), AUC | 1429.94 (1118.13-1828.70) | 15172.76 (12772.45-18024.15) | 7696.02 (6085.21-9733.22) |
| **Day 15 Visit (14 days post-boost)** | | | |
| N (non-missing) | 52 | 50 | 50 |
| Median ($P_{25}$, $P_{75}$), AUC | 42315.00 (33166.50-51865.00) | 50759.00 (46336.00-55780.00) | 49369.00 (42693.00-62433.00) |
| Minimum - Maximum, AUC | 16847.00-67696.00 | 32687.00-67612.00 | 26082.00-68586.00 |
| Geometric Mean (95% CI), AUC | 40907.72 (37505.30-44618.81) | 50557.92 (48214.00-53015.79) | 48494.17 (44985.03-52277.04) |
| N* (non-missing pre- and post-boost) | 52 | 50 | 50 |
| Participants with ≥ 2-fold rise[2], 95% CI | 100.0% (93.2%-100.0%) | 84.0% (70.9%-92.8%) | 94.0% (83.5%-98.7%) |
| Participants with ≥ 4-fold rise[2], 95% CI | 98.1% (89.7%-100.0%) | 42.0% (28.2%-56.8%) | 80.0% (66.3%-90.0%) |
| Geometric Mean Fold Rise[2], 95% CI | 28.61 (22.67-36.10) | 3.33 (2.87-3.87) | 6.30 (5.15-7.70) |
| **Day 29 Visit (28 days post-boost)** | | | |
| N (non-missing) | 43 | 48 | 43 |
| Median ($P_{25}$, $P_{75}$), AUC | 34555.00 (25813.00-43421.00) | 44963.00 (40354.00-48919.50) | 45423.00 (35744.00-53931.00) |
| Minimum - Maximum, AUC | 9150.00-62317.00 | 27937.00-64575.00 | 22181.00-64597.00 |
| Geometric Mean (95% CI), AUC | 32495.50 (28850.58-36600.91) | 44553.87 (42196.15-47043.32) | 43063.77 (39413.11-47052.59) |
| N* (non-missing pre- and post-boost) | 42 | 47 | 43 |
| Participants with ≥ 2-fold rise[2], 95% CI | 100.0% (91.6%-100.0%) | 70.2% (55.1%-82.7%) | 95.3% (84.2%-99.4%) |
| Participants with ≥ 4-fold rise[2], 95% CI | 95.2% (83.8%-99.4%) | 29.8% (17.3%-44.9%) | 74.4% (58.8%-86.5%) |
| Geometric Mean Fold Rise[2], 95% CI | 22.91 (17.50-29.99) | 2.89 (2.49-3.36) | 5.63 (4.54-6.98) |

[1] Relative to pre-vaccination (Day 1 Visit) levels, among participants with no missing observations at both pre- and post-boost timepoints.

**Supplemental Table 29:** Variants of Concern: mRNA-1273 IgG Serum Binding Antibody Response to S-2P-B.1.617.2 by FFP 10-plex ECLIA by Group, Age Group and Timepoint – Results are reported as Area Under Curve (AUC)

| | Group 1E [Dosed Janssen, Boost Moderna] Age 18-55 yo (N=21) | Group 1E [Dosed Janssen, Boost Moderna] Age ≥56 yo (N=32) | Group 2E [Dosed Moderna, Boost Moderna] Age 18-55 yo (N=26) | Group 2E [Dosed Moderna, Boost Moderna] Age ≥56 yo (N=25) | Group 3E [Dosed Pfizer, Boost Moderna] Age 18-55 yo (N=25) | Group 3E [Dosed Pfizer, Boost Moderna] Age ≥56 yo (N=25) |
|---|---|---|---|---|---|---|
| **Day 1 Visit (Pre-boost)** | | | | | | |
| N (non-missing) | 21 | 31 | 26 | 24 | 25 | 25 |
| Median ($P_{25}$, $P_{75}$), AUC | 1250.00 (776.60-2467.00) | 1141.00 (670.00-2373.00) | 14338.50 (9532.00-24934.00) | 14039.00 (9040.00-26412.00) | 8950.00 (6876.00-11391.00) | 8085.00 (2922.00-12748.00) |
| Minimum - Maximum, AUC | 412.00-11609.00 | 379.30-14547.00 | 5125.00-48856.00 | 4217.00-44714.00 | 551.80-38878.00 | 1517.00-39146.00 |
| Geometric Mean (95% CI), AUC | 1524.34 (1027.61-2261.20) | 1369.33 (981.94-1909.54) | 15714.65 (12470.98-19802.00) | 14606.78 (11098.06-19224.80) | 8697.59 (6236.81-12129.30) | 6809.78 (4804.84-9651.34) |
| **Day 15 Visit (14 days post-boost)** | | | | | | |
| N (non-missing) | 21 | 31 | 26 | 24 | 25 | 25 |
| Median ($P_{25}$, $P_{75}$), AUC | 43991.00 (39883.00-52980.00) | 41212.00 (30214.00-51259.00) | 51340.00 (48818.00-54865.00) | 48959.50 (44133.00-55992.50) | 55425.00 (43714.00-63179.00) | 47082.00 (36742.00-57327.00) |
| Minimum - Maximum, AUC | 25830.00-66703.00 | 16847.00-67696.00 | 38734.00-67612.00 | 32687.00-67195.00 | 34440.00-67899.00 | 26082.00-68586.00 |
| Geometric Mean (95% CI), AUC | 43660.88 (39044.00-48823.71) | 39142.00 (34467.45-44450.52) | 52074.41 (49485.42-54798.84) | 48964.85 (44996.81-53282.81) | 52877.84 (48832.00-57258.88) | 44473.91 (39298.38-50331.05) |
| N* (non-missing pre- and post-boost) | 21 | 31 | 26 | 24 | 25 | 25 |
| Participants with ≥ 2-fold rise[2], 95% CI | 100.0% (83.9%-100.0%) | 100.0% (88.8%-100.0%) | 84.6% (65.1%-95.6%) | 83.3% (62.6%-95.3%) | 92.0% (74.0%-99.0%) | 96.0% (79.6%-99.9%) |
| Participants with ≥ 4-fold rise[2], 95% CI | 95.2% (76.2%-99.9%) | 100.0% (88.8%-100.0%) | 42.3% (23.4%-63.1%) | 41.7% (22.1%-63.4%) | 76.0% (54.9%-90.6%) | 84.0% (63.9%-95.5%) |
| Geometric Mean Fold Rise[2], 95% CI | 28.64 (19.67-41.71) | 28.58 (20.89-39.11) | 3.31 (2.69-4.08) | 3.35 (2.66-4.22) | 6.08 (4.42-8.36) | 6.53 (4.99-8.55) |
| **Day 29 Visit (28 days post-boost)** | | | | | | |
| N (non-missing) | 18 | 25 | 25 | 23 | 21 | 22 |
| Median ($P_{25}$, $P_{75}$), AUC | 36225.00 (28498.00-42428.00) | 33879.00 (22820.00-43543.00) | 45409.00 (41882.00-49421.00) | 43373.00 (37682.00-48418.00) | 48744.00 (37682.00-53931.00) | 41008.50 (31930.00-51300.00) |
| Minimum - Maximum, AUC | 20733.00-58763.00 | 9150.00-62317.00 | 28569.00-64575.00 | 27937.00-63530.00 | 28305.00-64597.00 | 22181.00-63636.00 |
| Geometric Mean (95% CI), AUC | 35434.06 (30920.88-40605.98) | 30531.84 (25400.64-36699.58) | 45542.75 (42458.45-48851.09) | 43503.34 (39802.40-47548.39) | 46726.80 (42160.63-51787.50) | 39835.45 (34526.34-45960.93) |
| N* (non-missing pre- and post-boost) | 18 | 24 | 25 | 22 | 21 | 22 |
| Participants with ≥ 2-fold rise[2], 95% CI | 100.0% (81.5%-100.0%) | 100.0% (85.8%-100.0%) | 72.0% (50.6%-87.9%) | 68.2% (45.1%-86.1%) | 95.2% (76.2%-99.9%) | 95.5% (77.2%-99.9%) |

[1] Relative to pre-vaccination (Day 1 Visit) levels, among participants with no missing observations at both pre- and post-boost timepoints.

**Supplemental Table 30:** Variants of Concern: Ad26.COV2.S IgG Serum Binding Antibody Response to S-2P-B.1.617.2 by FFP 10-plex ECLIA by Group, and Timepoint – Results are reported as Area Under Curve (AUC)

| | Group 4E [Dosed Janssen, Boost Janssen] (N=50) | Group 5E [Dosed Moderna, Boost Janssen] (N=49) | Group 6E [Dosed Pfizer, Boost Janssen] (N=51) |
|---|---|---|---|
| **Day 1 Visit (Pre-boost)** | | | |
| N (non-missing) | 50 | 49 | 51 |
| Median ($P_{25}$, $P_{75}$), AUC | 1875.00 (800.40-2706.00) | 11832.00 (8763.00-20078.00) | 6838.00 (5090.00-11000.00) |
| Minimum - Maximum, AUC | 8.50-47338.00 | 1551.00-35757.00 | 930.40-32652.00 |
| Geometric Mean (95% CI), AUC | 1485.86 (1040.83-2121.16) | 11948.60 (9858.92-14481.21) | 7121.54 (5782.42-8770.80) |
| **Day 15 Visit (14 days post-boost)** | | | |
| N (non-missing) | 50 | 49 | 50 |
| Median ($P_{25}$, $P_{75}$), AUC | 7000.00 (4367.00-10211.00) | 33747.00 (23795.00-46331.00) | 26955.50 (18223.00-40665.00) |
| Minimum - Maximum, AUC | 33.82-43505.00 | 14199.00-69524.00 | 7031.00-55058.00 |
| Geometric Mean (95% CI), AUC | 6269.28 (4670.91-8414.61) | 32833.40 (29392.07-36677.65) | 25498.94 (21934.14-29643.10) |
| N* (non-missing pre- and post-boost) | 50 | 49 | 50 |
| Participants with ≥ 2-fold rise[1], 95% CI | 88.0% (75.7%-95.5%) | 61.2% (46.2%-74.8%) | 78.0% (64.0%-88.5%) |
| Participants with ≥ 4-fold rise[1], 95% CI | 48.0% (33.7%-62.6%) | 24.5% (13.3%-38.9%) | 44.0% (30.0%-58.7%) |
| Geometric Mean Fold Rise[1], 95% CI | 4.22 (3.41-5.22) | 2.75 (2.26-3.34) | 3.69 (2.98-4.57) |
| **Day 29 Visit (28 days post-boost)** | | | |
| N (non-missing) | 48 | 47 | 50 |
| Median ($P_{25}$, $P_{75}$), AUC | 7257.50 (4686.00-9413.50) | 40515.00 (29706.00-52266.00) | 36397.00 (22725.00-43173.00) |
| Minimum - Maximum, AUC | 960.20-42226.00 | 13719.00-72036.00 | 10427.00-62507.00 |
| Geometric Mean (95% CI), AUC | 6755.88 (5448.46-8377.03) | 39441.68 (35539.50-43772.32) | 30718.44 (26765.96-35254.58) |
| N* (non-missing pre- and post-boost) | 48 | 47 | 50 |
| Participants with ≥ 2-fold rise[1], 95% CI | 91.7% (80.0%-97.7%) | 72.3% (57.4%-84.4%) | 86.0% (73.3%-94.2%) |
| Participants with ≥ 4-fold rise[1], 95% CI | 47.9% (33.3%-62.8%) | 34.0% (20.9%-49.3%) | 48.0% (33.7%-62.6%) |
| Geometric Mean Fold Rise[1], 95% CI | 4.64 (3.61-5.96) | 3.24 (2.63-4.00) | 4.25 (3.40-5.32) |

[1] Relative to pre-vaccination (Day 1 Visit) levels, among participants with no missing observations at both pre- and post-boost timepoints.

**Supplemental Table 31:** Variants of Concern: Ad26.COV2.S IgG Serum Binding Antibody Response to S-2P-B.1.617.2 by FFP 10-plex ECLIA by Group, Age Group, and Timepoint – Results are reported as Area Under Curve (AUC)

| | Group 4E [Dosed Janssen, Boost Janssen] Age 18-55 yo (N=26) | Group 4E [Dosed Janssen, Boost Janssen] Age ≥56 yo (N=24) | Group 5E [Dosed Moderna, Boost Janssen] Age 18-55 yo (N=24) | Group 5E [Dosed Moderna, Boost Janssen] Age ≥56 yo (N=25) | Group 6E [Dosed Pfizer, Boost Janssen] Age 18-55 yo (N=25) | Group 6E [Dosed Pfizer, Boost Janssen] Age ≥56 yo (N=26) |
|---|---|---|---|---|---|---|
| **Day 1 Visit (Pre-boost)** | | | | | | |
| N (non-missing) | 26 | 24 | 24 | 25 | 25 | 26 |
| Median ($P_{25}$, $P_{75}$), AUC | 1898.50 (927.70-2599.00) | 1875.00 (682.70-2905.00) | 15032.50 (9199.50-23326.50) | 11401.00 (8075.00-15795.00) | 8115.00 (6209.00-11676.00) | 5572.50 (3402.00-8583.00) |
| Minimum - Maximum, AUC | 204.50-47338.00 | 8.50-33237.00 | 2689.00-35757.00 | 1551.00-23946.00 | 2657.00-32652.00 | 930.40-31786.00 |
| Geometric Mean (95% CI), AUC | 1597.17 (1047.35-2435.64) | 1374.01 (740.43-2549.74) | 14026.60 (10600.32-18560.34) | 10243.94 (7829.85-13402.35) | 9010.87 (6973.58-11643.34) | 5679.53 (4134.08-7802.72) |
| **Day 15 Visit (14 days post-boost)** | | | | | | |
| N (non-missing) | 26 | 24 | 24 | 25 | 24 | 26 |
| Median ($P_{25}$, $P_{75}$), AUC | 7420.50 (4004.00-10002.00) | 6656.50 (4558.50-10843.00) | 37376.00 (24439.50-49330.00) | 28081.00 (23035.00-39797.00) | 30556.50 (20080.50-43254.50) | 24654.50 (15815.00-34496.00) |
| Minimum - Maximum, AUC | 963.20-43505.00 | 33.82-35239.00 | 15317.00-69524.00 | 14199.00-60096.00 | 7031.00-55058.00 | 8630.00-54759.00 |
| Geometric Mean (95% CI), AUC | 6571.26 (4783.48-9027.21) | 5957.77 (3487.85-10176.77) | 35829.01 (30377.34-42259.07) | 30193.51 (25932.83-35154.21) | 27278.34 (21635.80-34392.43) | 23959.61 (19462.92-29495.21) |
| N* (non-missing pre- and post-boost) | 26 | 24 | 24 | 25 | 24 | 26 |
| Participants with ≥ 2-fold rise[1], 95% CI | 88.5% (69.8%-97.6%) | 87.5% (67.6%-97.3%) | 54.2% (32.8%-74.4%) | 68.0% (46.5%-85.1%) | 75.0% (53.3%-90.2%) | 80.8% (60.6%-93.4%) |
| Participants with ≥ 4-fold rise[1], 95% CI | 50.0% (29.9%-70.1%) | 45.8% (25.6%-67.2%) | 20.8% (7.1%-42.2%) | 28.0% (12.1%-49.4%) | 41.7% (22.1%-63.4%) | 46.2% (26.6%-66.6%) |
| Geometric Mean Fold Rise[1], 95% CI | 4.11 (3.10-5.45) | 4.34 (3.07-6.13) | 2.55 (1.92-3.40) | 2.95 (2.21-3.92) | 3.19 (2.43-4.20) | 4.22 (3.03-5.88) |
| **Day 29 Visit (28 days post-boost)** | | | | | | |
| N (non-missing) | 24 | 24 | 23 | 24 | 25 | 25 |
| Median ($P_{25}$, $P_{75}$), AUC | 7240.00 (3744.50-9748.50) | 7257.50 (5168.00-9323.50) | 41849.00 (36358.00-57300.00) | 38005.00 (27786.00-45899.00) | 35962.00 (23923.00-43450.00) | 36832.00 (20483.00-40641.00) |
| Minimum - Maximum, AUC | 1211.00-42226.00 | 960.20-33558.00 | 22428.00-72036.00 | 13719.00-65792.00 | 10427.00-62507.00 | 12036.00-57163.00 |
| Geometric Mean (95% CI), AUC | 6545.32 (4719.97-9076.58) | 6973.22 (5143.36-9454.11) | 42905.54 (37291.61-49364.60) | 36384.85 (31114.47-42547.97) | 32303.18 (26696.73-39087.01) | 29211.45 (23657.53-36069.22) |
| N* (non-missing pre- and post-boost) | 24 | 24 | 23 | 24 | 25 | 25 |
| Participants with ≥ 2-fold rise[1], 95% CI | 91.7% (73.0%-99.0%) | 91.7% (73.0%-99.0%) | 65.2% (42.7%-83.6%) | 79.2% (57.8%-92.9%) | 84.0% (63.9%-95.5%) | 88.0% (68.8%-97.5%) |
| Participants with ≥ 4-fold rise[1], 95% CI | 50.0% (29.1%-70.9%) | 45.8% (25.6%-67.2%) | 26.1% (10.2%-48.4%) | 41.7% (22.1%-63.4%) | 44.0% (24.4%-65.1%) | 52.0% (31.3%-72.2%) |
| Geometric Mean Fold Rise[1], 95% CI | 4.23 (3.18-5.64) | 5.08 (3.29-7.83) | 2.93 (2.20-3.90) | 3.57 (2.59-4.93) | 3.58 (2.75-4.68) | 5.04 (3.48-7.29) |

[1] Relative to pre-vaccination (Day 1 Visit) levels, among participants with no missing observations at both pre- and post-boost timepoints.

**Supplemental Table 32:** Variants of Concern: BNT162b2 IgG Serum Binding Antibody Response to S-2P-B.1.617.2 by FFP 10-plex ECLIA by Group, and Timepoint – Results are reported as Area Under Curve (AUC)

| | Group 7E [Dosed Janssen, Boost Pfizer] (N=53) | Group 8E [Dosed Moderna, Boost Pfizer] (N=51) | Group 9E [Dosed Pfizer, Boost Pfizer] (N=50) |
|---|---|---|---|
| **Day 1 Visit (Pre-boost)** | | | |
| N (non-missing) | 53 | 51 | 50 |
| Median ($P_{25}$, $P_{75}$), AUC | 2090.00 (1102.00-2955.00) | 13629.00 (7405.00-17406.00) | 6433.50 (4252.00-10855.00) |
| Minimum - Maximum, AUC | 95.44-46836.00 | 3954.00-29343.00 | 998.40-36991.00 |
| Geometric Mean (95% CI), AUC | 1845.42 (1385.10-2458.72) | 12196.16 (10499.06-14167.57) | 6454.35 (5287.39-7878.87) |
| **Day 15 Visit (14 days post-boost)** | | | |
| N (non-missing) | 50 | 48 | 48 |
| Median ($P_{25}$, $P_{75}$), AUC | 38821.00 (32263.00-43989.00) | 46301.00 (38130.50-52349.50) | 40201.00 (33542.50-50197.50) |
| Minimum - Maximum, AUC | 11892.00-67654.00 | 30565.00-66759.00 | 19000.00-63834.00 |
| Geometric Mean (95% CI), AUC | 35446.38 (31962.21-39310.35) | 45451.87 (43014.66-48027.17) | 39405.78 (36050.51-43073.32) |
| N* (non-missing pre- and post-boost) | 50 | 48 | 48 |
| Participants with ≥ 2-fold rise[1], 95% CI | 98.0% (89.4%-99.9%) | 95.8% (85.7%-99.5%) | 95.8% (85.7%-99.5%) |
| Participants with ≥ 4-fold rise[1], 95% CI | 96.0% (86.3%-99.5%) | 37.5% (24.0%-52.6%) | 77.1% (62.7%-88.0%) |
| Geometric Mean Fold Rise[1], 95% CI | 18.57 (14.41-23.92) | 3.70 (3.23-4.24) | 5.97 (4.98-7.15) |

[1] Relative to pre-vaccination (Day 1 Visit) levels, among participants with no missing observations at both pre- and post-boost timepoints.

**Supplemental Table 33:** Variants of Concern: BNT162b2 IgG Serum Binding Antibody Response to S-2P-B.1.617.2 by FFP 10-plex ECLIA by Group, Age Group, and Timepoint – Results are reported as Area Under Curve (AUC)

| | Group 7E [Dosed Janssen, Boost Pfizer] Age 18-55 yo (N=31) | Group 7E [Dosed Janssen, Boost Pfizer] Age ≥56 yo (N=22) | Group 8E [Dosed Moderna, Boost Pfizer] Age 18-55 yo (N=22) | Group 8E [Dosed Moderna, Boost Pfizer] Age ≥56 yo (N=29) | Group 9E [Dosed Pfizer, Boost Pfizer] Age 18-55 yo (N=24) | Group 9E [Dosed Pfizer, Boost Pfizer] Age ≥56 yo (N=26) |
|---|---|---|---|---|---|---|
| **Day 1 Visit (Pre-boost)** | | | | | | |
| N (non-missing) | 31 | 22 | 22 | 29 | 24 | 26 |
| Median ($P_{25}$, $P_{75}$), AUC | 2209.00 (1108.00-3313.00) | 1506.00 (1075.00-2712.00) | 16398.00 (12659.00-20728.00) | 12115.00 (6283.00-16045.00) | 8030.00 (4941.50-12396.50) | 5097.00 (3170.00-7654.00) |
| Minimum - Maximum, AUC | 246.40-46836.00 | 95.44-8892.00 | 7081.00-29090.00 | 3954.00-29343.00 | 2873.00-36991.00 | 998.40-14898.00 |
| Geometric Mean (95% CI), AUC | 2088.91 (1386.48-3147.19) | 1549.71 (1030.96-2329.49) | 15840.57 (13553.24-18513.94) | 10001.91 (8066.74-12401.33) | 8372.13 (6350.18-11037.90) | 5076.45 (3876.31-6648.17) |
| **Day 15 Visit (14 days post-boost)** | | | | | | |
| N (non-missing) | 28 | 22 | 20 | 28 | 23 | 25 |
| Median ($P_{25}$, $P_{75}$), AUC | 40147.50 (33120.50-44797.50) | 36658.00 (29942.00-40070.00) | 48182.50 (44037.50-52458.00) | 44743.00 (37355.50-50971.00) | 42958.00 (35384.00-50746.00) | 39122.00 (32010.00-43458.00) |
| Minimum - Maximum, AUC | 12392.00-66657.00 | 11892.00-67654.00 | 36787.00-66759.00 | 30565.00-63244.00 | 19571.00-63834.00 | 19000.00-61469.00 |
| Geometric Mean (95% CI), AUC | 37547.83 (33295.22-42343.61) | 32941.02 (27354.65-39668.24) | 47855.92 (44459.67-51511.60) | 43808.99 (40465.10-47429.21) | 41699.17 (36547.98-47576.39) | 37407.42 (32986.71-42420.56) |
| N* (non-missing pre- and post-boost) | 28 | 22 | 20 | 28 | 23 | 25 |
| Participants with ≥ 2-fold rise[1], 95% CI | 96.4% (81.7%-99.9%) | 100.0% (84.6%-100.0%) | 95.0% (75.1%-99.9%) | 96.4% (81.7%-99.9%) | 91.3% (72.0%-98.9%) | 100.0% (86.3%-100.0%) |
| Participants with ≥ 4-fold rise[1], 95% CI | 92.9% (76.5%-99.1%) | 100.0% (84.6%-100.0%) | 15.0% (3.2%-37.9%) | 53.6% (33.9%-72.5%) | 69.6% (47.1%-86.8%) | 84.0% (63.9%-95.5%) |
| Geometric Mean Fold Rise[1], 95% CI | 16.69 (11.50-24.23) | 21.26 (14.93-30.26) | 2.86 (2.50-3.27) | 4.45 (3.68-5.38) | 4.84 (3.79-6.20) | 7.23 (5.61-9.33) |

[1] Relative to pre-vaccination (Day 1 Visit) levels, among participants with no missing observations at both pre- and post-boost timepoints.

**Supplemental Table 34:** mRNA-1273 IgG Serum Binding Antibody Response to S-2P-B.351 by 4-plex ECLIA V.2 by Group and Timepoint – Results are reported as Arbitrary Units (AU/mL)

| | Group 1E [Dosed Janssen, Boost Moderna] (N=53) | Group 2E [Dosed Moderna, Boost Moderna] (N=51) | Group 3E [Dosed Pfizer, Boost Moderna] (N=50) |
|---|---|---|---|
| **Day 1 Visit (Pre-boost)** | | | |
| N (non-missing) | 52 | 50 | 50 |
| Median ($P_{25}$, $P_{75}$), AU/mL | 1957.13 (1356.14-4533.44) | 31524.96 (20022.23-64506.15) | 19390.99 (9419.07-30827.52) |
| Minimum - Maximum, AU/mL | 480.55-37058.45 | 10719.23-763362.65 | 1061.79-149256.77 |
| Geometric Mean (95% CI), AU/mL | 2377.59 (1854.65-3047.99) | 38611.93 (29788.81-50048.35) | 17836.45 (13344.14-23841.09) |
| **Day 15 Visit (14 days post-boost)** | | | |
| N (non-missing) | 52 | 50 | 50 |
| Median ($P_{25}$, $P_{75}$), AU/mL | 173972.91 (92998.32-315135.27) | 311604.33 (253345.94-420943.33) | 338950.54 (175715.92-664499.32) |
| Minimum - Maximum, AU/mL | 11195.05-1116017.71 | 151282.31-1588556.65 | 70588.24-1493816.81 |
| Geometric Mean (95% CI), AU/mL | 164609.25 (126577.33-214068.38) | 335546.16 (289442.52-388993.39) | 351184.95 (281100.91-438742.33) |
| N* (non-missing pre- and post-boost) | 52 | 50 | 50 |
| Participants with ≥ 2-fold rise[2], 95% CI | 100.0% (93.2%-100.0%) | 98.0% (89.4%-99.9%) | 98.0% (89.4%-99.9%) |
| Participants with ≥ 4-fold rise[2], 95% CI | 100.0% (93.2%-100.0%) | 82.0% (68.6%-91.4%) | 98.0% (89.4%-99.9%) |
| Geometric Mean Fold Rise[2], 95% CI | 69.23 (49.95-95.97) | 8.69 (6.77-11.16) | 19.69 (15.41-25.16) |
| **Day 29 Visit (28 days post-boost)** | | | |
| N (non-missing) | 43 | 48 | 43 |
| Median ($P_{25}$, $P_{75}$), AU/mL | 140880.23 (72913.00-216242.32) | 276079.02 (207035.64-402206.56) | 260739.75 (165860.48-526827.38) |
| Minimum - Maximum, AU/mL | 27932.41-595373.80 | 120383.65-985545.37 | 80635.07-1430145.19 |
| Geometric Mean (95% CI), AU/mL | 136677.97 (110286.61-169384.73) | 286200.12 (246077.31-332864.93) | 290695.66 (233400.02-362056.38) |
| N* (non-missing pre- and post-boost) | 42 | 47 | 43 |
| Participants with ≥ 2-fold rise[2], 95% CI | 100.0% (91.6%-100.0%) | 97.9% (88.7%-99.9%) | 97.7% (87.7%-99.9%) |
| Participants with ≥ 4-fold rise[2], 95% CI | 97.6% (87.4%-99.9%) | 78.7% (64.3%-89.3%) | 97.7% (87.7%-99.9%) |
| Geometric Mean Fold Rise[2], 95% CI | 58.16 (42.89-78.87) | 7.18 (5.58-9.23) | 17.20 (13.36-22.15) |

[1] Antibody values below the lower limit of quantification (LLOQ = 10.254 AU/mL) were assigned a value equal to LLOQ/2. Values greater than the upper limit of quantification (ULOQ = 5740875 AU/mL) are taken as reported, or a ceiling value equivalent to the ULOQ is assigned if values are not provided.
[2] Relative to pre-vaccination (Day 1 Visit) levels, among participants with no missing observations at both pre- and post-boost timepoints.

**Supplemental Table 35:** mRNA-1273 IgG Serum Binding Antibody Response to S-2P- B.351 by 4-plex ECLIA V.2 by Group, Age Group, and Timepoint – Results are reported as Arbitrary Units (AU/mL)

| | Group 1E [Dosed Janssen, Boost Moderna] Age 18-55 yo (N=21) | Group 1E [Dosed Janssen, Boost Moderna] Age ≥56 yo (N=32) | Group 2E [Dosed Moderna, Boost Moderna] Age 18-55 yo (N=26) | Group 2E [Dosed Moderna, Boost Moderna] Age ≥56 yo (N=25) | Group 3E [Dosed Pfizer, Boost Moderna] Age 18-55 yo (N=25) | Group 3E [Dosed Pfizer, Boost Moderna] Age ≥56 yo (N=25) |
|---|---|---|---|---|---|---|
| **Day 1 Visit (Pre-boost)** | | | | | | |
| N (non-missing) | 21 | 31 | 26 | 24 | 25 | 25 |
| Median ($P_{25}$, $P_{75}$), AU/mL | 1998.35 (1401.58-3129.67) | 1865.21 (1291.92-5100.42) | 31513.18 (20057.93-62099.75) | 37565.95 (18512.43-74737.02) | 23015.84 (13448.81-36572.07) | 15282.88 (7358.28-24168.26) |
| Minimum - Maximum, AU/mL | 713.15-22286.72 | 480.55-37058.45 | 12735.80-209983.35 | 10719.23-763362.65 | 1061.79-149256.77 | 2565.02-99270.94 |
| Geometric Mean (95% CI), AU/mL | 2404.74 (1606.74-3599.06) | 2359.38 (1689.16-3295.53) | 36005.89 (26331.32-49235.05) | 41648.41 (26685.19-65001.95) | 22930.78 (14820.06-35480.35) | 13873.87 (9422.41-20428.37) |
| **Day 15 Visit (14 days post-boost)** | | | | | | |
| N (non-missing) | 21 | 31 | 26 | 24 | 25 | 25 |
| Median ($P_{25}$, $P_{75}$), AU/mL | 176941.57 (77797.50-282713.03) | 154176.29 (102914.34-333642.39) | 315396.71 (268581.71-373093.62) | 302647.76 (186210.35-476733.27) | 475332.75 (303023.73-802945.04) | 270475.03 (139251.88-406768.32) |
| Minimum - Maximum, AU/mL | 11195.05-727992.53 | 59366.78-1116017.71 | 181765.04-1279337.11 | 151282.31-1588556.65 | 97346.76-1493816.81 | 70588.24-1433707.12 |
| Geometric Mean (95% CI), AU/mL | 137839.88 (81062.85-234383.97) | 185639.02 (140729.06-244880.81) | 336779.24 (286442.31-395961.95) | 334215.43 (255749.00-436756.18) | 450444.95 (338483.78-599439.80) | 273797.87 (196582.76-381342.06) |
| N* (non-missing pre- and post-boost) | 21 | 31 | 26 | 24 | 25 | 25 |
| Participants with ≥ 2-fold rise[2], 95% CI | 100.0% (83.9%-100.0%) | 100.0% (88.8%-100.0%) | 100.0% (86.8%-100.0%) | 95.8% (78.9%-99.9%) | 96.0% (79.6%-99.9%) | 100.0% (86.3%-100.0%) |
| Participants with ≥ 4-fold rise[2], 95% CI | 100.0% (83.9%-100.0%) | 100.0% (88.8%-100.0%) | 84.6% (65.1%-95.6%) | 79.2% (57.8%-92.9%) | 96.0% (79.6%-99.9%) | 100.0% (86.3%-100.0%) |
| Geometric Mean Fold Rise[2], 95% CI | 57.32 (30.28-108.51) | 78.68 (54.66-113.27) | 9.35 (7.02-12.46) | 8.02 (5.16-12.47) | 19.64 (12.76-30.25) | 19.73 (15.07-25.84) |
| **Day 29 Visit (28 days post-boost)** | | | | | | |
| N (non-missing) | 18 | 25 | 25 | 23 | 21 | 22 |
| Median ($P_{25}$, $P_{75}$), AU/mL | 164820.43 (120619.82-216242.32) | 115561.68 (70141.59-203588.67) | 273877.44 (211865.61-361683.43) | 292418.82 (194288.65-433458.39) | 328802.72 (174472.79-556011.28) | 213982.90 (165022.36-368690.07) |
| Minimum - Maximum, AU/mL | 58329.38-376224.81 | 27932.41-595373.80 | 129071.56-985545.37 | 120383.65-705208.99 | 80635.07-1006829.14 | 99056.77-1430145.19 |
| Geometric Mean (95% CI), AU/mL | 156704.79 (121549.73-202027.54) | 123863.18 (88993.56-172395.49) | 295136.63 (239102.66-364302.22) | 276793.16 (219113.46-349656.52) | 326519.70 (237383.29-449126.44) | 260172.77 (188743.46-358634.26) |
| N* (non-missing pre- and post-boost) | 18 | 24 | 25 | 22 | 21 | 22 |
| Participants with ≥ 2-fold rise[2], 95% CI | 100.0% (81.5%-100.0%) | 100.0% (85.8%-100.0%) | 100.0% (86.3%-100.0%) | 95.5% (77.2%-99.9%) | 95.2% (76.2%-99.9%) | 100.0% (84.6%-100.0%) |

[1] Antibody values below the lower limit of quantification (LLOQ = 10.254 AU/mL) were assigned a value equal to LLOQ/2. Values greater than the upper limit of quantification (ULOQ = 5740875 AU/mL) are taken as reported, or a ceiling value equivalent to the ULOQ is assigned if values are not provided.
[2] Relative to pre-vaccination (Day 1 Visit) levels, among participants with no missing observations at both pre- and post-boost timepoints.

**Supplemental Table 36:** Ad26.COV2.S IgG Serum Binding Antibody Response to S-2P-B.351 by 4-plex ECLIA V.2 by Group and Timepoint – Results are reported as Arbitrary Units (AU/mL)

| | Group 4E [Dosed Janssen, Boost Janssen] (N=50) | Group 5E [Dosed Moderna, Boost Janssen] (N=49) | Group 6E [Dosed Pfizer, Boost Janssen] (N=51) |
|---|---|---|---|
| **Day 1 Visit (Pre-boost)** | | | |
| N (non-missing) | 50 | 49 | 51 |
| Median ($P_{25}$, $P_{75}$), AU/mL | 3736.75 (1412.97-5593.49) | 29529.79 (20883.74-51059.97) | 16525.07 (11593.35-27558.47) |
| Minimum - Maximum, AU/mL | 5.12-292072.17 | 4316.03-158701.48 | 1903.03-90620.45 |
| Geometric Mean (95% CI), AU/mL | 2924.81 (1935.04-4420.83) | 28906.08 (23180.03-36046.61) | 17257.52 (13706.93-21727.82) |
| **Day 15 Visit (14 days post-boost)** | | | |
| N (non-missing) | 50 | 49 | 50 |
| Median ($P_{25}$, $P_{75}$), AU/mL | 17892.28 (9242.73-25147.64) | 121496.14 (78706.00-267988.09) | 87172.77 (51372.88-207866.98) |
| Minimum - Maximum, AU/mL | 5.12-198352.75 | 36315.67-1379442.33 | 16945.86-519497.30 |
| Geometric Mean (95% CI), AU/mL | 15031.97 (10075.70-22426.24) | 138257.72 (110105.02-173608.78) | 99536.65 (77764.17-127405.01) |
| N* (non-missing pre- and post-boost) | 50 | 49 | 50 |
| Participants with ≥ 2-fold rise[2], 95% CI | 88.0% (75.7%-95.5%) | 81.6% (68.0%-91.2%) | 92.0% (80.8%-97.8%) |
| Participants with ≥ 4-fold rise[2], 95% CI | 58.0% (43.2%-71.8%) | 53.1% (38.3%-67.5%) | 56.0% (41.3%-70.0%) |
| Geometric Mean Fold Rise[2], 95% CI | 5.14 (3.99-6.63) | 4.78 (3.62-6.33) | 5.96 (4.53-7.84) |
| **Day 29 Visit (28 days post-boost)** | | | |
| N (non-missing) | 48 | 47 | 50 |
| Median ($P_{25}$, $P_{75}$), AU/mL | 17303.82 (10205.86-27264.98) | 152346.11 (101785.43-390262.88) | 123098.32 (61284.43-228209.90) |
| Minimum - Maximum, AU/mL | 1973.31-156007.31 | 34811.45-2580883.46 | 24145.26-548889.05 |
| Geometric Mean (95% CI), AU/mL | 16487.07 (12985.88-20932.23) | 183215.13 (142153.82-236137.05) | 119673.02 (94487.99-151570.91) |
| N* (non-missing pre- and post-boost) | 48 | 47 | 50 |
| Participants with ≥ 2-fold rise[2], 95% CI | 93.8% (82.8%-98.7%) | 85.1% (71.7%-93.8%) | 90.0% (78.2%-96.7%) |
| Participants with ≥ 4-fold rise[2], 95% CI | 58.3% (43.2%-72.4%) | 61.7% (46.4%-75.5%) | 64.0% (49.2%-77.1%) |
| Geometric Mean Fold Rise[2], 95% CI | 5.80 (4.31-7.81) | 6.22 (4.53-8.53) | 6.79 (5.12-8.98) |

[1] Antibody values below the lower limit of quantification (LLOQ = 10.254 AU/mL) were assigned a value equal to LLOQ/2. Values greater than the upper limit of quantification (ULOQ = 5740875 AU/mL) are taken as reported, or a ceiling value equivalent to the ULOQ is assigned if values are not provided.

[2] Relative to pre-vaccination (Day 1 Visit) levels, among participants with no missing observations at both pre- and post-boost timepoints.

**Supplemental Table 37:** Ad26.COV2.S IgG Serum Binding Antibody Response to S-2P-B.351 by 4-plex ECLIA V.2 by Group, Age Group, and Timepoint – Results are reported as Arbitrary Units (AU/mL)

| | Group 4E [Dosed Janssen, Boost Janssen] Age 18-55 yo (N=26) | Group 4E [Dosed Janssen, Boost Janssen] Age ≥56 yo (N=24) | Group 5E [Dosed Moderna, Boost Janssen] Age 18-55 yo (N=24) | Group 5E [Dosed Moderna, Boost Janssen] Age ≥56 yo (N=25) | Group 6E [Dosed Pfizer, Boost Janssen] Age 18-55 yo (N=25) | Group 6E [Dosed Pfizer, Boost Janssen] Age ≥56 yo (N=26) |
|---|---|---|---|---|---|---|
| **Day 1 Visit (Pre-boost)** | | | | | | |
| N (non-missing) | 26 | 24 | 24 | 25 | 25 | 26 |
| Median ($P_{25}$, $P_{75}$), AU/mL | 3578.62 (1412.97-5496.80) | 3856.29 (1380.04-5598.25) | 33268.51 (20848.28-61871.19) | 29529.79 (20883.74-37342.46) | 17490.70 (13823.16-31332.83) | 13766.31 (6707.58-21663.99) |
| Minimum - Maximum, AU/mL | 561.53-292072.17 | 5.12-63092.06 | 6051.53-158701.48 | 4316.03-70235.11 | 6466.42-90620.45 | 1903.03-85304.83 |
| Geometric Mean (95% CI), AU/mL | 3215.42 (1979.43-5223.20) | 2639.54 (1284.07-5425.86) | 34775.92 (24928.43-48513.48) | 24205.35 (17980.27-32585.65) | 21213.06 (15945.35-28220.99) | 14151.43 (9863.11-20304.25) |
| **Day 15 Visit (14 days post-boost)** | | | | | | |
| N (non-missing) | 26 | 24 | 24 | 25 | 24 | 26 |
| Median ($P_{25}$, $P_{75}$), AU/mL | 18303.14 (9242.73-22951.40) | 17376.54 (9741.25-28140.14) | 150537.80 (90529.36-354225.71) | 103658.87 (72999.71-160420.71) | 100079.26 (51532.32-235108.10) | 81180.35 (44949.80-150829.28) |
| Minimum - Maximum, AU/mL | 3022.48-198352.75 | 5.12-73796.63 | 36315.67-1379442.33 | 41526.23-431376.88 | 16945.86-440331.42 | 27123.92-519497.30 |
| Geometric Mean (95% CI), AU/mL | 16877.36 (11693.23-24359.85) | 13259.78 (6162.65-28530.23) | 164462.66 (112851.73-239677.04) | 117037.92 (89134.45-153676.54) | 108523.79 (73468.40-160305.84) | 91902.83 (65874.71-128215.07) |
| N* (non-missing pre- and post-boost) | 26 | 24 | 24 | 25 | 24 | 26 |
| Participants with ≥ 2-fold rise[2], 95% CI | 88.5% (69.8%-97.6%) | 87.5% (67.6%-97.3%) | 87.5% (67.6%-97.3%) | 76.0% (54.9%-90.6%) | 87.5% (67.6%-97.3%) | 96.2% (80.4%-99.9%) |
| Participants with ≥ 4-fold rise[2], 95% CI | 65.4% (44.3%-82.8%) | 50.0% (29.1%-70.9%) | 41.7% (22.1%-63.4%) | 64.0% (42.5%-82.0%) | 50.0% (29.1%-70.9%) | 61.5% (40.6%-79.8%) |
| Geometric Mean Fold Rise[2], 95% CI | 5.25 (3.78-7.30) | 5.02 (3.31-7.64) | 4.73 (3.07-7.29) | 4.84 (3.28-7.14) | 5.43 (3.66-8.05) | 6.49 (4.32-9.77) |
| **Day 29 Visit (28 days post-boost)** | | | | | | |
| N (non-missing) | 24 | 24 | 23 | 24 | 25 | 25 |
| Median ($P_{25}$, $P_{75}$), AU/mL | 17632.84 (8218.90-26443.80) | 17303.82 (12379.85-27327.81) | 161849.83 (125270.49-390870.18) | 146286.50 (91084.35-278454.11) | 116455.09 (69418.68-238297.24) | 128132.77 (47252.77-202959.40) |
| Minimum - Maximum, AU/mL | 4129.22-156007.31 | 1973.31-57220.75 | 50952.67-2580883.46 | 34811.45-706108.31 | 24145.26-548889.05 | 26950.35-479601.92 |
| Geometric Mean (95% CI), AU/mL | 16275.68 (11185.93-23681.33) | 16701.20 (12068.31-23112.61) | 212658.29 (141963.62-318557.33) | 158831.65 (114057.34-221182.55) | 127926.21 (90618.65-180593.23) | 111952.28 (79173.27-158302.34) |
| N* (non-missing pre- and post-boost) | 24 | 24 | 23 | 24 | 25 | 25 |
| Participants with ≥ 2-fold rise[2], 95% CI | 91.7% (73.0%-99.0%) | 95.8% (78.9%-99.9%) | 91.3% (72.0%-98.9%) | 79.2% (57.8%-92.9%) | 88.0% (68.8%-97.5%) | 92.0% (74.0%-99.0%) |

[1] Antibody values below the lower limit of quantification (LLOQ = 10.254 AU/mL) were assigned a value equal to LLOQ/2. Values greater than the upper limit of quantification (ULOQ = 5740875 AU/mL) are taken as reported, or a ceiling value equivalent to the ULOQ is assigned if values are not provided.

[2] Relative to pre-vaccination (Day 1 Visit) levels, among participants with no missing observations at both pre- and post-boost timepoints.

**Supplemental Table 38:** BNT162b2 IgG Serum Binding Antibody Response to S-2P-B.351 by 4-plex ECLIA V.2 by Group and Timepoint – Results are reported as Arbitrary Units (AU/mL)

| | Group 7E [Dosed Janssen, Boost Pfizer] (N=53) | Group 8E [Dosed Moderna, Boost Pfizer] (N=51) | Group 9E [Dosed Pfizer, Boost Pfizer] (N=50) |
|---|---|---|---|
| **Day 1 Visit (Pre-boost)** | | | |
| N (non-missing) | 53 | 51 | 50 |
| Median ($P_{25}$, $P_{75}$), AU/mL | 3501.53 (1605.09-6512.64) | 27416.05 (14512.33-41354.71) | 12124.07 (6884.80-17548.85) |
| Minimum - Maximum, AU/mL | 153.79-233523.48 | 6453.21-70730.37 | 1215.49-77816.98 |
| Geometric Mean (95% CI), AU/mL | 3303.47 (2404.08-4539.32) | 24561.32 (20531.67-29381.85) | 11593.12 (9332.11-14401.92) |
| **Day 15 Visit (14 days post-boost)** | | | |
| N (non-missing) | 50 | 48 | 48 |
| Median ($P_{25}$, $P_{75}$), AU/mL | 138300.52 (94369.20-222477.63) | 260506.87 (171791.35-344510.73) | 186335.19 (116251.94-310363.78) |
| Minimum - Maximum, AU/mL | 25136.17-1375820.54 | 96109.28-1090776.68 | 40455.53-962595.99 |
| Geometric Mean (95% CI), AU/mL | 138860.95 (110544.88-174430.19) | 251139.29 (215464.09-292721.35) | 190199.69 (153906.09-235051.93) |
| N* (non-missing pre- and post-boost) | 50 | 48 | 48 |
| Participants with ≥ 2-fold rise[2], 95% CI | 98.0% (89.4%-99.9%) | 100.0% (92.6%-100.0%) | 100.0% (92.6%-100.0%) |
| Participants with ≥ 4-fold rise[2], 95% CI | 98.0% (89.4%-99.9%) | 95.8% (85.7%-99.5%) | 97.9% (88.9%-99.9%) |
| Geometric Mean Fold Rise[2], 95% CI | 40.45 (30.06-54.43) | 10.15 (8.52-12.08) | 16.12 (12.79-20.32) |

[1] Antibody values below the lower limit of quantification (LLOQ = 10.254 AU/mL) were assigned a value equal to LLOQ/2. Values greater than the upper limit of quantification (ULOQ = 5740875 AU/mL) are taken as reported, or a ceiling value equivalent to the ULOQ is assigned if values are not provided.

[2] Relative to pre-vaccination (Day 1 Visit) levels, among participants with no missing observations at both pre- and post-boost timepoints.

**Supplemental Table 39:** BNT162b2 IgG Serum Binding Antibody Response to S-2P-B.351 by 4-plex ECLIA V.2 by Group, Age Group and Timepoint – Results are reported as Arbitrary Units (AU/mL)

| | **Group 7E [Dosed Janssen, Boost Pfizer] Age 18-55 yo (N=31)** | **Group 7E [Dosed Janssen, Boost Pfizer] Age ≥56 yo (N=22)** | **Group 8E [Dosed Moderna, Boost Pfizer] Age 18-55 yo (N=22)** | **Group 8E [Dosed Moderna, Boost Pfizer] Age ≥56 yo (N=29)** | **Group 9E [Dosed Pfizer, Boost Pfizer] Age 18-55 yo (N=24)** | **Group 9E [Dosed Pfizer, Boost Pfizer] Age ≥56 yo (N=26)** |
|---|---|---|---|---|---|---|
| **Day 1 Visit (Pre-boost)** | | | | | | |
| N (non-missing) | 31 | 22 | 22 | 29 | 24 | 26 |
| Median ($P_{25}$, $P_{75}$), AU/mL | 3515.44 (1783.67-6513.96) | 2334.28 (1507.00-5536.31) | 35588.39 (23893.26-44611.47) | 19394.15 (11152.78-33241.50) | 12705.32 (8996.08-24720.49) | 9331.22 (6647.80-16290.59) |
| Minimum - Maximum, AU/mL | 376.57-233523.48 | 153.79-18718.63 | 12992.99-57568.83 | 6453.21-70730.37 | 4694.31-77816.98 | 1215.49-32624.23 |
| Geometric Mean (95% CI), AU/mL | 3812.10 (2437.96-5960.77) | 2699.81 (1696.83-4295.63) | 32064.07 (26456.18-38860.67) | 20064.50 (15391.88-26155.63) | 14571.76 (10609.14-20014.46) | 9387.02 (7003.45-12581.81) |
| **Day 15 Visit (14 days post-boost)** | | | | | | |
| N (non-missing) | 28 | 22 | 20 | 28 | 23 | 25 |
| Median ($P_{25}$, $P_{75}$), AU/mL | 145235.51 (108303.76-231482.17) | 108136.61 (80678.22-174995.31) | 275866.96 (255456.89-358770.28) | 202901.01 (158362.46-331259.32) | 200328.63 (130896.70-399337.36) | 184620.16 (107618.72-265279.88) |
| Minimum - Maximum, AU/mL | 36872.59-1375820.54 | 25136.17-1222689.23 | 96109.28-1090776.68 | 97789.63-613507.78 | 40455.53-962595.99 | 49885.22-543888.63 |
| Geometric Mean (95% CI), AU/mL | 156512.38 (117001.54-209365.85) | 119244.49 (81532.68-174399.38) | 284210.56 (226108.61-357242.67) | 229900.15 (186095.89-284015.30) | 208610.90 (148526.68-293001.27) | 174699.96 (132046.52-231131.23) |
| N* (non-missing pre- and post-boost) | 28 | 22 | 20 | 28 | 23 | 25 |
| Participants with ≥ 2-fold rise[2], 95% CI | 96.4% (81.7%-99.9%) | 100.0% (84.6%-100.0%) | 100.0% (83.2%-100.0%) | 100.0% (87.7%-100.0%) | 100.0% (85.2%-100.0%) | 100.0% (86.3%-100.0%) |
| Participants with ≥ 4-fold rise[2], 95% CI | 96.4% (81.7%-99.9%) | 100.0% (84.6%-100.0%) | 95.0% (75.1%-99.9%) | 96.4% (81.7%-99.9%) | 95.7% (78.1%-99.9%) | 100.0% (86.3%-100.0%) |
| Geometric Mean Fold Rise[2], 95% CI | 37.74 (24.56-58.02) | 44.17 (28.65-68.09) | 8.10 (6.43-10.21) | 11.92 (9.34-15.20) | 13.97 (10.00-19.53) | 18.38 (13.15-25.69) |

[1] Antibody values below the lower limit of quantification (LLOQ = 10.254 AU/mL) were assigned a value equal to LLOQ/2. Values greater than the upper limit of quantification (ULOQ = 5740875 AU/mL) are taken as reported, or a ceiling value equivalent to the ULOQ is assigned if values are not provided.

[2] Relative to pre-vaccination (Day 1 Visit) levels, among participants with no missing observations at both pre- and post-boost timepoints.

Supplemental Table 40: **SARS-CoV-2 Pseudovirion Neutralization Antibody -** mRNA-1273 Neutralization Antibody Response (International Units/ml for 50% neutralization, IU50/ml) to Pseudovirus D614G[1], by Group and Timepoint

| | Group 1E [Dosed Janssen, Boost Moderna] (N=53) | Group 2E [Dosed Moderna, Boost Moderna] (N=51) | Group 3E [Dosed Pfizer, Boost Moderna] (N=50) |
|---|---|---|---|
| **Day 1 Visit (Pre-boost)** | | | |
| N (non-missing) | 52 | 50 | 50 |
| Median ($P_{25}$, $P_{75}$) | 8.30 (3.85-16.17) | 82.60 (42.65-180.68) | 28.68 (14.55-39.17) |
| Minimum - Maximum | 1.21-1589.04 | 9.89-1174.00 | 1.21-676.16 |
| Geometric Mean (95% CI) | 8.91 (6.19-12.82) | 88.65 (67.78-115.93) | 24.79 (17.96-34.21) |
| **Day 15 Visit (14 days post-boost)** | | | |
| N (non-missing) | 52 | 50 | 50 |
| Median ($P_{25}$, $P_{75}$) | 791.05 (264.84-1214.46) | 867.04 (634.26-1073.84) | 859.27 (439.77-1171.36) |
| Minimum - Maximum | 136.45-7207.20 | 166.13-6587.61 | 118.44-5430.32 |
| Geometric Mean (95% CI) | 676.12 (517.53-883.31) | 901.81 (727.54-1117.83) | 785.76 (596.43-1035.20) |
| **Day 29 Visit (28 days post-boost)** | | | |
| N (non-missing) | 43 | 48 | 43 |
| Median ($P_{25}$, $P_{75}$) | 501.71 (200.10-797.01) | 725.99 (417.50-1041.86) | 525.91 (201.77-1140.77) |
| Minimum - Maximum | 49.33-3189.84 | 119.08-3793.02 | 85.88-2604.75 |
| Geometric Mean (95% CI) | 431.65 (322.59-577.59) | 700.01 (568.58-861.82) | 495.72 (370.39-663.47) |

1 Calibration to the WHO standard International Units (IU50/mL and IU80/mL) was done using a calibration factor of 0.242 for ID50 and a calibration factor of 1.502 for ID80, both specific to the SARS-CoV-2 D614G pseudovirus.

2 Values below the lower limit of detection (LLOD = 2.42) were assigned the value of LLOD/2. Values between the LLOD and the lower limit of quantification (LLOQ = 4.477) are taken as reported, or a value of LLOQ/2 is assigned if observations are reported as <LLOQ but no value is provided. Values greater than the upper limit of quantification (ULOQ = 10919) are taken as reported, or a ceiling value equivalent to the ULOQ is assigned if values are not provided.

Supplemental Table 41: **SARS-CoV-2 Pseudovirion Neutralization Antibody -** mRNA-1273 Neutralization Antibody Response (International Units/ml for 50% neutralization, IU50/ml) to Pseudovirus D614G[1], by Group, Age Group and Timepoint

| | Group 1E [Dosed Janssen, Boost Moderna] Age 18-55 yo (N=21) | Group 1E [Dosed Janssen, Boost Moderna] Age ≥56 yo (N=32) | Group 2E [Dosed Moderna, Boost Moderna] Age 18-55 yo (N=26) | Group 2E [Dosed Moderna, Boost Moderna] Age ≥56 yo (N=25) | Group 3E [Dosed Pfizer, Boost Moderna] Age 18-55 yo (N=25) | Group 3E [Dosed Pfizer, Boost Moderna] Age ≥56 yo (N=25) |
|---|---|---|---|---|---|---|
| **Day 1 Visit (Pre-boost)** | | | | | | |
| N (non-missing) | 21 | 31 | 26 | 24 | 25 | 25 |
| Median ($P_{25}$, $P_{75}$) | 8.28 (3.39-17.07) | 8.46 (3.98-15.80) | 86.52 (42.65-183.44) | 80.68 (38.55-170.64) | 31.76 (20.50-42.45) | 19.44 (10.40-37.90) |
| Minimum - Maximum | 1.21-1589.04 | 1.21-132.66 | 25.25-1174.00 | 9.89-390.63 | 1.21-676.16 | 1.21-264.05 |
| Geometric Mean (95% CI) | 9.33 (4.34-20.07) | 8.63 (5.97-12.49) | 92.40 (64.19-133.02) | 84.75 (55.43-129.57) | 31.42 (19.68-50.14) | 19.56 (12.39-30.88) |
| **Day 15 Visit (14 days post-boost)** | | | | | | |
| N (non-missing) | 21 | 31 | 26 | 24 | 25 | 25 |
| Median ($P_{25}$, $P_{75}$) | 793.03 (375.84-1279.03) | 711.43 (249.58-1199.00) | 867.04 (724.10-1170.13) | 859.07 (580.51-1057.25) | 934.26 (854.75-1209.55) | 577.51 (252.83-957.42) |
| Minimum - Maximum | 136.45-7207.20 | 193.85-4264.81 | 264.81-4702.89 | 166.13-6587.61 | 175.58-5430.32 | 118.44-3752.66 |
| Geometric Mean (95% CI) | 756.60 (475.95-1202.75) | 626.53 (445.79-880.54) | 953.87 (743.66-1223.51) | 848.62 (582.24-1236.86) | 1077.25 (760.11-1526.70) | 573.15 (379.29-866.08) |
| **Day 29 Visit (28 days post-boost)** | | | | | | |
| N (non-missing) | 18 | 25 | 25 | 23 | 21 | 22 |
| Median ($P_{25}$, $P_{75}$) | 599.37 (217.56-785.64) | 446.44 (186.05-797.01) | 766.85 (456.75-1058.74) | 725.75 (379.88-976.35) | 792.37 (282.64-1150.29) | 423.62 (162.09-873.98) |
| Minimum - Maximum | 152.57-3189.84 | 49.33-2875.00 | 119.08-3793.02 | 237.00-3424.50 | 115.59-2237.34 | 85.88-2604.75 |
| Geometric Mean (95% CI) | 509.26 (326.10-795.29) | 383.21 (255.57-574.58) | 691.81 (511.55-935.59) | 709.04 (520.35-966.16) | 614.53 (419.99-899.17) | 403.81 (257.05-634.35) |

[1] Calibration to the WHO standard International Units (IU50/mL and IU80/mL) was done using a calibration factor of 0.242 for ID50 and a calibration factor of 1.502 for ID80, both specific to the SARS-CoV-2 D614G pseudovirus.

[2] Values below the lower limit of detection (LLOD = 2.42) were assigned the value of LLOD/2. Values between the LLOD and the lower limit of quantification (LLOQ = 4.477) are taken as reported, or a value of LLOQ/2 is assigned if observations are reported as <LLOQ but no value is provided. Values greater than the upper limit of quantification (ULOQ = 10919) are taken as reported, or a ceiling value equivalent to the ULOQ is assigned if values are not provided.

Supplemental Table 42: **SARS-CoV-2 Pseudovirion Neutralization Antibody –** Ad26.COV2.S Neutralization Antibody Response (International Units/ml for 50% neutralization, IU50/ml) to Pseudovirus D614G[1], by Group and Timepoint

| | Group 4E [Dosed Janssen, Boost Janssen] (N=50) | Group 5E [Dosed Moderna, Boost Janssen] (N=49) | Group 6E [Dosed Pfizer, Boost Janssen] (N=51) |
|---|---|---|---|
| **Day 1 Visit (Pre-boost)** | | | |
| N (non-missing) | 50 | 49 | 51 |
| Median ($P_{25}$, $P_{75}$) | 7.35 (2.77-13.75) | 59.01 (38.73-108.99) | 19.42 (8.72-34.48) |
| Minimum - Maximum | 1.21-2291.64 | 5.39-621.28 | 1.21-569.93 |
| Geometric Mean (95% CI) | 7.57 (4.87-11.75) | 61.69 (44.98-84.61) | 18.55 (13.37-25.72) |
| **Day 15 Visit (14 days post-boost)** | | | |
| N (non-missing) | 50 | 49 | 50 |
| Median ($P_{25}$, $P_{75}$) | 30.46 (19.32-40.63) | 385.53 (183.86-706.08) | 194.80 (99.01-480.33) |
| Minimum - Maximum | 1.21-2573.17 | 45.30-4177.09 | 29.10-5220.93 |
| Geometric Mean (95% CI) | 31.42 (22.28-44.33) | 382.07 (290.49-502.52) | 216.38 (157.82-296.68) |

1 Calibration to the WHO standard International Units (IU50/mL and IU80/mL) was done using a calibration factor of 0.242 for ID50 and a calibration factor of 1.502 for ID80, both specific to the SARS-CoV-2 D614G pseudovirus.

2 Values below the lower limit of detection (LLOD = 2.42) were assigned the value of LLOD/2. Values between the LLOD and the lower limit of quantification (LLOQ = 4.477) are taken as reported, or a value of LLOQ/2 is assigned if observations are reported as <LLOQ but no value is provided. Values greater than the upper limit of quantification (ULOQ = 10919) are taken as reported, or a ceiling value equivalent to the ULOQ is assigned if values are not provided.

Supplemental Table 43: **SARS-CoV-2 Pseudovirion Neutralization Antibody –** Ad26.COV2.S Neutralization Antibody Response (International Units/ml for 50% neutralization, IU50/ml) to Pseudovirus D614G[1], by Group, Age Group and Timepoint

| | Group 4E [Dosed Janssen, Boost Janssen] Age 18-55 yo (N=26) | Group 4E [Dosed Janssen, Boost Janssen] Age ≥56 yo (N=24) | Group 5E [Dosed Moderna, Boost Janssen] Age 18-55 yo (N=24) | Group 5E [Dosed Moderna, Boost Janssen] Age ≥56 yo (N=25) | Group 6E [Dosed Pfizer, Boost Janssen] Age 18-55 yo (N=25) | Group 6E [Dosed Pfizer, Boost Janssen] Age ≥56 yo (N=26) |
|---|---|---|---|---|---|---|
| **Day 1 Visit (Pre-boost)** | | | | | | |
| N (non-missing) | 26 | 24 | 24 | 25 | 25 | 26 |
| Median ($P_{25}$, $P_{75}$) | 7.11 (2.77-11.24) | 7.58 (2.14-20.49) | 74.88 (41.98-152.82) | 49.87 (35.20-92.15) | 22.74 (15.68-39.03) | 15.01 (7.85-29.82) |
| Minimum - Maximum | 1.21-834.19 | 1.21-2291.64 | 7.79-621.28 | 5.39-408.96 | 3.58-569.93 | 1.21-207.53 |
| Geometric Mean (95% CI) | 6.88 (3.94-11.99) | 8.39 (4.02-17.52) | 75.04 (48.10-117.08) | 51.11 (32.03-81.54) | 24.21 (15.69-37.36) | 14.35 (8.74-23.56) |
| **Day 15 Visit (14 days post-boost)** | | | | | | |
| N (non-missing) | 26 | 24 | 24 | 25 | 24 | 26 |
| Median ($P_{25}$, $P_{75}$) | 31.83 (17.07-41.00) | 29.24 (23.96-39.95) | 459.18 (189.31-941.30) | 273.18 (173.41-687.24) | 340.93 (110.68-531.22) | 165.78 (99.01-439.80) |
| Minimum - Maximum | 6.30-1202.80 | 1.21-2573.17 | 97.07-4177.09 | 45.30-1263.88 | 29.10-5220.93 | 35.05-1048.57 |
| Geometric Mean (95% CI) | 28.96 (18.44-45.48) | 34.33 (19.64-60.01) | 467.53 (305.82-714.75) | 314.76 (219.01-452.39) | 255.23 (150.99-431.43) | 185.79 (125.46-275.13) |

1 Calibration to the WHO standard International Units (IU50/mL and IU80/mL) was done using a calibration factor of 0.242 for ID50 and a calibration factor of 1.502 for ID80, both specific to the SARS-CoV-2 D614G pseudovirus.

2 Values below the lower limit of detection (LLOD = 2.42) were assigned the value of LLOD/2. Values between the LLOD and the lower limit of quantification (LLOQ = 4.477) are taken as reported, or a value of LLOQ/2 is assigned if observations are reported as <LLOQ but no value is provided. Values greater than the upper limit of quantification (ULOQ = 10919) are taken as reported, or a ceiling value equivalent to the ULOQ is assigned if values are not provided.

Supplemental Table 44: **SARS-CoV-2 Pseudovirion Neutralization Antibody –** BNT162b2 Antibody Response (International Units/ml for 50% neutralization, IU50/ml) to Pseudovirus D614G[1], by Group and Timepoint

| | Group 7E [Dosed Janssen, Boost Pfizer] (N=53) | Group 8E [Dosed Moderna, Boost Pfizer] (N=51) | Group 9E [Dosed Pfizer, Boost Pfizer] (N=50) |
|---|---|---|---|
| **Day 1 Visit (Pre-boost)** | | | |
| N (non-missing) | 53 | 51 | 50 |
| Median ($P_{25}$, $P_{75}$) | 8.08 (5.20-14.44) | 54.86 (33.00-121.10) | 23.10 (8.07-38.37) |
| Minimum - Maximum | 1.21-1112.57 | 7.03-236.40 | 1.21-494.63 |
| Geometric Mean (95% CI) | 9.37 (6.44-13.64) | 57.61 (45.04-73.68) | 21.42 (15.29-30.00) |
| **Day 15 Visit (14 days post-boost)** | | | |
| N (non-missing) | 50 | 48 | 48 |
| Median ($P_{25}$, $P_{75}$) | 353.87 (199.76-628.55) | 754.13 (417.19-1109.54) | 465.47 (225.09-861.98) |
| Minimum - Maximum | 1.21-5616.56 | 106.91-1869.00 | 36.91-3057.04 |
| Geometric Mean (95% CI) | 341.34 (239.58-486.31) | 677.85 (559.44-821.31) | 446.65 (340.27-586.28) |

[1] Calibration to the WHO standard International Units (IU50/mL and IU80/mL) was done using a calibration factor of 0.242 for ID50 and a calibration factor of 1.502 for ID80, both specific to the SARS-CoV-2 D614G pseudovirus.

[2] Values below the lower limit of detection (LLOD = 2.42) were assigned the value of LLOD/2. Values between the LLOD and the lower limit of quantification (LLOQ = 4.477) are taken as reported, or a value of LLOQ/2 is assigned if observations are reported as <LLOQ but no value is provided. Values greater than the upper limit of quantification (ULOQ = 10919) are taken as reported, or a ceiling value equivalent to the ULOQ is assigned if values are not provided.

Supplemental Table 45: **SARS-CoV-2 Pseudovirion Neutralization Antibody –** BNT162b2 Neutralization Antibody Response (International Units/ml for 50% neutralization, IU50/ml) to Pseudovirus D614G[1], by Group, Age Group and Timepoint

| | Group 7E [Dosed Janssen, Boost Pfizer] Age 18-55 yo (N=21) | Group 7E [Dosed Janssen, Boost Pfizer] Age ≥56 yo (N=32) | Group 8E [Dosed Moderna, Boost Pfizer] Age 18-55 yo (N=26) | Group 8E [Dosed Moderna, Boost Pfizer] Age ≥56 yo (N=25) | Group 9E [Dosed Pfizer, Boost Pfizer] Age 18-55 yo (N=25) | Group 9E [Dosed Pfizer, Boost Pfizer] Age ≥56 yo (N=25) |
|---|---|---|---|---|---|---|
| **Day 1 Visit (Pre-boost)** | | | | | | |
| N (non-missing) | 31 | 22 | 22 | 29 | 24 | 26 |
| Median ($P_{25}$, $P_{75}$) | 6.69 (3.49-16.21) | 8.36 (6.19-13.75) | 101.62 (50.76-145.76) | 39.86 (25.34-83.81) | 25.49 (14.72-73.93) | 19.45 (6.20-33.30) |
| Minimum - Maximum | 1.21-1112.57 | 1.21-72.88 | 30.83-188.74 | 7.03-236.40 | 3.69-185.27 | 1.21-494.63 |
| Geometric Mean (95% CI) | 8.90 (4.93-16.07) | 10.07 (6.66-15.24) | 84.48 (64.87-110.03) | 43.08 (30.04-61.79) | 29.56 (18.73-46.65) | 15.91 (9.71-26.06) |
| **Day 15 Visit (14 days post-boost)** | | | | | | |
| N (non-missing) | 28 | 22 | 20 | 28 | 23 | 25 |
| Median ($P_{25}$, $P_{75}$) | 327.07 (199.85-721.20) | 353.87 (198.90-628.55) | 991.39 (602.63-1287.31) | 639.79 (328.35-1004.40) | 710.67 (172.40-899.16) | 365.79 (244.93-785.69) |
| Minimum - Maximum | 1.21-5616.56 | 40.12-4645.35 | 274.10-1869.00 | 106.91-1738.45 | 99.60-3057.04 | 36.91-1489.52 |
| Geometric Mean (95% CI) | 327.47 (189.36-566.29) | 359.85 (228.49-566.70) | 864.68 (663.21-1127.35) | 569.66 (437.97-740.94) | 502.24 (320.17-787.84) | 400.95 (283.87-566.31) |

[1] Calibration to the WHO standard International Units (IU50/mL and IU80/mL) was done using a calibration factor of 0.242 for ID50 and a calibration factor of 1.502 for ID80, both specific to the SARS-CoV-2 D614G pseudovirus.

[2] Values below the lower limit of detection (LLOD = 2.42) were assigned the value of LLOD/2. Values between the LLOD and the lower limit of quantification (LLOQ = 4.477) are taken as reported, or a value of LLOQ/2 is assigned if observations are reported as <LLOQ but no value is provided. Values greater than the upper limit of quantification (ULOQ = 10919) are taken as reported, or a ceiling value equivalent to the ULOQ is assigned if values are not provided.

Supplemental Table 46: **SARS-CoV-2 Pseudovirion Neutralization Antibody –** mRNA-1273 Antibody Response (International Units/ml for 80% neutralization, IU80/ml) to Pseudovirus D614G[1], by Group and Timepoint

| | Group 1E [Dosed Janssen, Boost Moderna] (N=53) | Group 2E [Dosed Moderna, Boost Moderna] (N=51) | Group 3E [Dosed Pfizer, Boost Moderna] (N=50) |
|---|---|---|---|
| **Day 1 Visit (Pre-boost)** | | | |
| N (non-missing) | 52 | 50 | 50 |
| Median ($P_{25}$, $P_{75}$) | 20.28 (7.51-29.79) | 160.84 (99.03-417.93) | 72.49 (26.44-101.48) |
| Minimum - Maximum | 7.51-1726.84 | 26.04-2246.50 | 7.51-968.95 |
| Geometric Mean (95% CI) | 20.01 (14.69-27.26) | 182.40 (140.07-237.52) | 54.23 (39.64-74.17) |
| **Day 15 Visit (14 days post-boost)** | | | |
| N (non-missing) | 52 | 50 | 50 |
| Median ($P_{25}$, $P_{75}$) | 2160.21 (640.75-2788.27) | 2399.61 (1356.91-2821.59) | 2124.84 (779.63-3099.58) |
| Minimum - Maximum | 360.32-10761.37 | 499.36-14106.93 | 245.26-14060.20 |
| Geometric Mean (95% CI) | 1541.33 (1195.32-1987.50) | 2132.32 (1725.70-2634.75) | 1794.74 (1357.69-2372.49) |
| **Day 29 Visit (28 days post-boost)** | | | |
| N (non-missing) | 43 | 48 | 43 |
| Median ($P_{25}$, $P_{75}$) | 858.37 (527.40-1509.17) | 1520.75 (814.82-2454.02) | 887.38 (547.37-2454.82) |
| Minimum - Maximum | 127.78-4598.67 | 394.01-5735.07 | 165.15-5225.61 |
| Geometric Mean (95% CI) | 872.16 (672.45-1131.19) | 1443.57 (1181.33-1764.01) | 1097.65 (817.62-1473.57) |

[1] Calibration to the WHO standard International Units (IU50/mL and IU80/mL) was done using a calibration factor of 0.242 for ID50 and a calibration factor of 1.502 for ID80, both specific to the SARS-CoV-2 D614G pseudovirus.

[2] Values below the lower limit of detection (LLOD = 2.42) were assigned the value of LLOD/2. Values between the LLOD and the lower limit of quantification (LLOQ = 4.477) are taken as reported, or a value of LLOQ/2 is assigned if observations are reported as <LLOQ but no value is provided. Values greater than the upper limit of quantification (ULOQ = 10919) are taken as reported, or a ceiling value equivalent to the ULOQ is assigned if values are not provided.

Supplemental Table 47: **SARS-CoV-2 Pseudovirion Neutralization Antibody –** mRNA-1273 Neutralization Antibody Response (International Units/ml for 80% neutralization, IU80/ml) to Pseudovirus D614G[1], by Group, Age Group and Timepoint

| | **Group 1E [Dosed Janssen, Boost Moderna] Age 18-55 yo (N=21)** | **Group 1E [Dosed Janssen, Boost Moderna] Age ≥56 yo (N=32)** | **Group 2E [Dosed Moderna, Boost Moderna] Age 18-55 yo (N=26)** | **Group 2E [Dosed Moderna, Boost Moderna] Age ≥56 yo (N=25)** | **Group 3E [Dosed Pfizer, Boost Moderna] Age 18-55 yo (N=25)** | **Group 3E [Dosed Pfizer, Boost Moderna] Age ≥56 yo (N=25)** |
|---|---|---|---|---|---|---|
| **Day 1 Visit (Pre-boost)** | | | | | | |
| N (non-missing) | 21 | 31 | 26 | 24 | 25 | 25 |
| Median ($P_{25}$, $P_{75}$) | 21.01 (7.51-30.93) | 19.76 (7.51-28.66) | 156.59 (112.32-417.93) | 163.80 (88.77-380.73) | 79.68 (39.88-97.16) | 36.81 (21.83-101.48) |
| Minimum - Maximum | 7.51-1726.84 | 7.51-192.19 | 60.56-2246.50 | 26.04-687.74 | 7.51-968.95 | 7.51-524.31 |
| Geometric Mean (95% CI) | 24.01 (13.23-43.58) | 17.69 (12.46-25.11) | 196.13 (137.56-279.64) | 168.61 (110.80-256.57) | 73.28 (48.57-110.56) | 40.13 (25.07-64.23) |
| **Day 15 Visit (14 days post-boost)** | | | | | | |
| N (non-missing) | 21 | 31 | 26 | 24 | 25 | 25 |
| Median ($P_{25}$, $P_{75}$) | 2272.42 (717.06-2763.79) | 1410.51 (606.43-2812.76) | 2461.49 (1695.05-2948.50) | 2194.94 (1091.73-2648.45) | 2568.71 (1943.14-3221.35) | 985.20 (606.99-2519.10) |
| Minimum - Maximum | 360.32-10529.95 | 380.64-10761.37 | 584.63-7706.82 | 499.36-14106.93 | 459.45-14060.20 | 245.26-11122.18 |
| Geometric Mean (95% CI) | 1718.31 (1125.89-2622.44) | 1431.92 (1025.41-1999.57) | 2236.43 (1771.59-2823.23) | 2025.00 (1383.56-2963.81) | 2558.66 (1791.23-3654.87) | 1258.91 (840.03-1886.65) |
| **Day 29 Visit (28 days post-boost)** | | | | | | |
| N (non-missing) | 18 | 25 | 25 | 23 | 21 | 22 |
| Median ($P_{25}$, $P_{75}$) | 1031.81 (566.16-1316.97) | 814.34 (398.17-1558.23) | 1491.87 (910.45-2620.19) | 1564.48 (711.76-2077.43) | 1991.70 (684.58-2682.83) | 756.47 (381.91-2097.39) |
| Minimum - Maximum | 219.21-4598.67 | 127.78-3840.32 | 394.01-5735.07 | 487.20-5538.36 | 232.91-5091.38 | 165.15-5225.61 |
| Geometric Mean (95% CI) | 1001.45 (680.40-1473.98) | 789.54 (546.18-1141.34) | 1486.82 (1125.16-1964.74) | 1397.98 (1023.81-1908.88) | 1426.08 (975.94-2083.83) | 854.97 (544.38-1342.75) |

[1] Calibration to the WHO standard International Units (IU50/mL and IU80/mL) was done using a calibration factor of 0.242 for ID50 and a calibration factor of 1.502 for ID80, both specific to the SARS-CoV-2 D614G pseudovirus.

[2] Values below the lower limit of detection (LLOD = 2.42) were assigned the value of LLOD/2. Values between the LLOD and the lower limit of quantification (LLOQ = 4.477) are taken as reported, or a value of LLOQ/2 is assigned if observations are reported as <LLOQ but no value is provided. Values greater than the upper limit of quantification (ULOQ = 10919) are taken as reported, or a ceiling value equivalent to the ULOQ is assigned if values are not provided.

Supplemental Table 48: **SARS-CoV-2 Pseudovirion Neutralization Antibody –** Ad26.COV.2 Neutralization Antibody Response (International Units/ml for 80% neutralization, IU80/ml) to Pseudovirus D614G[1], by Group and Timepoint

| | Group 4E [Dosed Janssen, Boost Janssen] (N=50) | Group 5E [Dosed Moderna, Boost Janssen] (N=49) | Group 6E [Dosed Pfizer, Boost Janssen] (N=51) |
|---|---|---|---|
| **Day 1 Visit (Pre-boost)** | | | |
| N (non-missing) | 50 | 49 | 51 |
| Median ($P_{25}$, $P_{75}$) | 20.81 (7.51-26.97) | 120.73 (85.19-177.23) | 36.97 (23.16-80.65) |
| Minimum - Maximum | 7.51-2950.97 | 7.51-909.19 | 7.51-939.44 |
| Geometric Mean (95% CI) | 20.01 (14.24-28.10) | 118.28 (89.09-157.03) | 40.72 (30.29-54.74) |
| **Day 15 Visit (14 days post-boost)** | | | |
| N (non-missing) | 50 | 49 | 50 |
| Median ($P_{25}$, $P_{75}$) | 83.66 (37.63-111.34) | 722.80 (476.33-1318.36) | 504.34 (169.79-901.64) |
| Minimum - Maximum | 7.51-1647.81 | 113.02-9595.98 | 93.59-8290.50 |
| Geometric Mean (95% CI) | 77.87 (58.32-103.98) | 795.37 (609.54-1037.87) | 487.03 (360.66-657.68) |

1 Calibration to the WHO standard International Units (IU50/mL and IU80/mL) was done using a calibration factor of 0.242 for ID50 and a calibration factor of 1.502 for ID80, both specific to the SARS-CoV-2 D614G pseudovirus.

2 Values below the lower limit of detection (LLOD = 2.42) were assigned the value of LLOD/2. Values between the LLOD and the lower limit of quantification (LLOQ = 4.477) are taken as reported, or a value of LLOQ/2 is assigned if observations are reported as <LLOQ but no value is provided. Values greater than the upper limit of quantification (ULOQ = 10919) are taken as reported, or a ceiling value equivalent to the ULOQ is assigned if values are not provided.

Supplemental Table 49: **SARS-CoV-2 Pseudovirion Neutralization Antibody –** Ad26.COV.2 Neutralization Antibody Response (International Units/ml for 80% neutralization, IU80/ml) to Pseudovirus D614G[1], by Group and Timepoint

| | Group 4E [Dosed Janssen, Boost Janssen] Age 18-55 yo (N=26) | Group 4E [Dosed Janssen, Boost Janssen] Age ≥56 yo (N=24) | Group 5E [Dosed Moderna, Boost Janssen] Age 18-55 yo (N=24) | Group 5E [Dosed Moderna, Boost Janssen] Age ≥56 yo (N=25) | Group 6E [Dosed Pfizer, Boost Janssen] Age 18-55 yo (N=25) | Group 6E [Dosed Pfizer, Boost Janssen] Age ≥56 yo (N=26) |
|---|---|---|---|---|---|---|
| **Day 1 Visit (Pre-boost)** | | | | | | |
| N (non-missing) | 26 | 24 | 24 | 25 | 25 | 26 |
| Median ($P_{25}$, $P_{75}$) | 19.54 (7.51-25.07) | 22.01 (7.51-35.59) | 133.91 (91.63-236.66) | 120.73 (75.54-153.57) | 40.33 (30.20-103.12) | 31.23 (18.00-58.93) |
| Minimum - Maximum | 7.51-961.67 | 7.51-2950.97 | 22.91-909.19 | 7.51-629.06 | 7.51-939.44 | 7.51-341.09 |
| Geometric Mean (95% CI) | 18.30 (11.96-27.99) | 22.04 (12.46-38.98) | 142.49 (96.45-210.50) | 98.92 (64.66-151.32) | 54.27 (35.92-81.98) | 30.89 (20.25-47.13) |
| **Day 15 Visit (14 days post-boost)** | | | | | | |
| N (non-missing) | 26 | 24 | 24 | 25 | 24 | 26 |
| Median ($P_{25}$, $P_{75}$) | 87.39 (34.59-117.76) | 83.66 (40.26-110.54) | 833.63 (523.94-2295.74) | 608.30 (475.45-971.66) | 688.48 (239.44-940.72) | 445.85 (169.79-766.18) |
| Minimum - Maximum | 20.83-1596.75 | 7.51-1647.81 | 190.71-9595.98 | 113.02-3080.22 | 100.66-8290.50 | 93.59-2709.98 |
| Geometric Mean (95% CI) | 75.33 (50.20-113.05) | 80.73 (51.80-125.81) | 985.35 (648.64-1496.83) | 647.55 (459.90-911.76) | 568.66 (352.56-917.20) | 422.12 (283.79-627.86) |

[1] Calibration to the WHO standard International Units (IU50/mL and IU80/mL) was done using a calibration factor of 0.242 for ID50 and a calibration factor of 1.502 for ID80, both specific to the SARS-CoV-2 D614G pseudovirus.

[2] Values below the lower limit of detection (LLOD = 2.42) were assigned the value of LLOD/2. Values between the LLOD and the lower limit of quantification (LLOQ = 4.477) are taken as reported, or a value of LLOQ/2 is assigned if observations are reported as <LLOQ but no value is provided. Values greater than the upper limit of quantification (ULOQ = 10919) are taken as reported, or a ceiling value equivalent to the ULOQ is assigned if values are not provided.

Supplemental Table 50: **SARS-CoV-2 Pseudovirion Neutralization Antibody –** BNT162b2 Neutralization Antibody Response (International Units/ml for 80% neutralization, IU80/ml) to Pseudovirus D614G[1], by Group and Timepoint

| | Group 7E [Dosed Janssen, Boost Pfizer] (N=53) | Group 8E [Dosed Moderna, Boost Pfizer] (N=51) | Group 9E [Dosed Pfizer, Boost Pfizer] (N=50) |
|---|---|---|---|
| **Day 1 Visit (Pre-boost)** | | | |
| N (non-missing) | 53 | 51 | 50 |
| Median ($P_{25}$, $P_{75}$) | 22.07 (15.52-29.43) | 131.10 (83.58-202.68) | 41.71 (23.44-101.08) |
| Minimum - Maximum | 7.51-2790.94 | 22.05-487.93 | 7.51-841.30 |
| Geometric Mean (95% CI) | 24.38 (18.00-33.03) | 115.35 (91.40-145.58) | 47.46 (35.11-64.15) |
| **Day 15 Visit (14 days post-boost)** | | | |
| N (non-missing) | 50 | 48 | 48 |
| Median ($P_{25}$, $P_{75}$) | 724.05 (524.76-1130.98) | 1777.29 (754.09-2655.07) | 799.05 (575.19-2239.57) |
| Minimum - Maximum | 7.51-9758.22 | 243.43-3251.20 | 112.42-5212.69 |
| Geometric Mean (95% CI) | 766.06 (560.86-1046.34) | 1453.65 (1194.75-1768.65) | 1012.31 (786.66-1302.69) |

1 Calibration to the WHO standard International Units (IU50/mL and IU80/mL) was done using a calibration factor of 0.242 for ID50 and a calibration factor of 1.502 for ID80, both specific to the SARS-CoV-2 D614G pseudovirus.

2 Values below the lower limit of detection (LLOD = 2.42) were assigned the value of LLOD/2. Values between the LLOD and the lower limit of quantification (LLOQ = 4.477) are taken as reported, or a value of LLOQ/2 is assigned if observations are reported as <LLOQ but no value is provided. Values greater than the upper limit of quantification (ULOQ = 10919) are taken as reported, or a ceiling value equivalent to the ULOQ is assigned if values are not provided.

Supplemental Table 51: **SARS-CoV-2 Pseudovirion Neutralization Antibody –** BNT162b2 Neutralization Antibody Response (International Units/ml for 80% neutralization, IU80/ml) to Pseudovirus D614G[1], by Group, Age Group and Timepoint

| | Group 7E [Dosed Janssen, Boost Pfizer] Age 18-55 yo (N=21) | Group 7E [Dosed Janssen, Boost Pfizer] Age ≥56 yo (N=32) | Group 8E [Dosed Moderna, Boost Pfizer] Age 18-55 yo (N=26) | Group 8E [Dosed Moderna, Boost Pfizer] Age ≥56 yo (N=25) | Group 9E [Dosed Pfizer, Boost Pfizer] Age 18-55 yo (N=25) | Group 9E [Dosed Pfizer, Boost Pfizer] Age ≥56 yo (N=25) |
|---|---|---|---|---|---|---|
| **Day 1 Visit (Pre-boost)** | | | | | | |
| N (non-missing) | 31 | 22 | 22 | 29 | 24 | 26 |
| Median ($P_{25}$, $P_{75}$) | 22.07 (7.51-29.57) | 21.83 (15.95-29.43) | 178.53 (115.91-226.78) | 99.17 (39.94-145.47) | 53.07 (30.83-132.14) | 33.27 (18.83-93.22) |
| Minimum - Maximum | 7.51-2790.94 | 7.51-141.79 | 83.58-455.82 | 22.05-487.93 | 7.51-406.71 | 7.51-841.30 |
| Geometric Mean (95% CI) | 25.16 (15.68-40.36) | 23.33 (16.43-33.13) | 175.04 (140.65-217.84) | 84.06 (59.82-118.13) | 61.89 (41.06-93.30) | 37.15 (23.84-57.87) |
| **Day 15 Visit (14 days post-boost)** | | | | | | |
| N (non-missing) | 28 | 22 | 20 | 28 | 23 | 25 |
| Median ($P_{25}$, $P_{75}$) | 690.24 (560.55-1617.99) | 732.77 (524.76-1130.98) | 2433.38 (1130.87-2702.94) | 1061.31 (675.47-2557.84) | 1310.23 (513.16-2602.46) | 749.83 (605.36-1530.39) |
| Minimum - Maximum | 7.51-9758.22 | 111.34-8612.65 | 623.00-3251.20 | 243.43-3204.07 | 208.63-5212.69 | 112.42-3294.62 |
| Geometric Mean (95% CI) | 782.17 (483.99-1264.06) | 746.03 (497.29-1119.21) | 1852.06 (1426.11-2405.22) | 1222.70 (929.41-1608.56) | 1166.90 (772.06-1763.66) | 888.24 (644.51-1224.14) |

1 Calibration to the WHO standard International Units (IU50/mL and IU80/mL) was done using a calibration factor of 0.242 for ID50 and a calibration factor of 1.502 for ID80, both specific to the SARS-CoV-2 D614G pseudovirus.

2 Values below the lower limit of detection (LLOD = 2.42) were assigned the value of LLOD/2. Values between the LLOD and the lower limit of quantification (LLOQ = 4.477) are taken as reported, or a value of LLOQ/2 is assigned if observations are reported as <LLOQ but no value is provided. Values greater than the upper limit of quantification (ULOQ = 10919) are taken as reported, or a ceiling value equivalent to the ULOQ is assigned if values are not provided.

Supplemental Table 52: **SARS-CoV-2 Pseudovirion Neutralization Antibody –** mRNA-1273 Neutralization Antibodies Titer (ID50) to Pseudovirus D614G[1], by Group and Timepoint

| | Group 1E [Dosed Janssen, Boost Moderna] (N=53) | Group 2E [Dosed Moderna, Boost Moderna] (N=51) | Group 3E [Dosed Pfizer, Boost Moderna] (N=50) |
|---|---|---|---|
| **Day 1 Visit (Pre-boost)** | | | |
| N (non-missing) | 52 | 50 | 50 |
| Median ($P_{25}$, $P_{75}$) | 34.30 (15.89-66.83) | 341.31 (176.25-746.61) | 118.50 (60.11-161.87) |
| Minimum - Maximum | 5.00-6566.30 | 40.88-4851.22 | 5.00-2794.06 |
| Geometric Mean (95% CI) | 36.81 (25.58-52.96) | 366.31 (280.09-479.06) | 102.44 (74.23-141.38) |
| **Day 15 Visit (14 days post-boost)** | | | |
| N (non-missing) | 52 | 50 | 50 |
| Median ($P_{25}$, $P_{75}$) | 3268.82 (1094.40-5018.44) | 3582.81 (2620.92-4437.34) | 3550.70 (1817.21-4840.33) |
| Minimum - Maximum | 563.83-29781.84 | 686.48-27221.54 | 489.42-22439.32 |
| Geometric Mean (95% CI) | 2793.90 (2138.57-3650.03) | 3726.50 (3006.38-4619.12) | 3246.95 (2464.58-4277.67) |
| N* (non-missing pre- and post-boost) | 52 | 50 | 50 |
| Participants with ≥ 2-fold rise[2], 95% CI | 100.0% (93.2%-100.0%) | 96.0% (86.3%-99.5%) | 100.0% (92.9%-100.0%) |
| Participants with ≥ 4-fold rise[2], 95% CI | 100.0% (93.2%-100.0%) | 86.0% (73.3%-94.2%) | 100.0% (92.9%-100.0%) |
| Geometric Mean Fold Rise[2], 95% CI | 75.91 (54.99-104.78) | 10.17 (8.05-12.85) | 31.69 (23.80-42.21) |
| **Day 29 Visit (28 days post-boost)** | | | |
| N (non-missing) | 43 | 48 | 43 |
| Median ($P_{25}$, $P_{75}$) | 2073.19 (826.88-3293.42) | 2999.98 (1725.22-4305.19) | 2173.20 (833.78-4713.91) |
| Minimum - Maximum | 203.86-13181.15 | 492.07-15673.64 | 354.88-10763.42 |
| Geometric Mean (95% CI) | 1783.68 (1333.00-2386.74) | 2892.62 (2349.52-3561.26) | 2048.44 (1530.54-2741.59) |
| N* (non-missing pre- and post-boost) | 42 | 47 | 43 |
| Participants with ≥ 2-fold rise[2], 95% CI | 97.6% (87.4%-99.9%) | 95.7% (85.5%-99.5%) | 100.0% (91.8%-100.0%) |
| Participants with ≥ 4-fold rise[2], 95% CI | 92.9% (80.5%-98.5%) | 80.9% (66.7%-90.9%) | 100.0% (91.8%-100.0%) |
| Geometric Mean Fold Rise[2], 95% CI | 45.06 (29.36-69.14) | 7.46 (5.92-9.39) | 20.94 (15.36-28.55) |

[1] Values below the lower limit of detection (LLOD = 10) were assigned the value of LLOD/2. Values between the LLOD and the lower limit of quantification (LLOQ = 18.5) are taken as reported, or a value of LLOQ/2 is assigned if observations are reported as <LLOQ but no value is provided. Values greater than the upper limit of quantification (ULOQ = 45118) are taken as reported, or a ceiling value equivalent to the ULOQ is assigned if values are not provided.

Supplemental Table 53: **SARS-CoV-2 Pseudovirion Neutralization Antibody –** mRNA-1273 Neutralization Antibodies Titer (ID50) to Pseudovirus D614G[1], by Group, Age Group and Timepoint

| | **Group 1E [Dosed Janssen, Boost Moderna] Age 18-55 yo (N=21)** | **Group 1E [Dosed Janssen, Boost Moderna] Age ≥56 yo (N=32)** | **Group 2E [Dosed Moderna, Boost Moderna] Age 18-55 yo (N=26)** | **Group 2E [Dosed Moderna, Boost Moderna] Age ≥56 yo (N=25)** | **Group 3E [Dosed Pfizer, Boost Moderna] Age 18-55 yo (N=25)** | **Group 3E [Dosed Pfizer, Boost Moderna] Age ≥56 yo (N=25)** |
|---|---|---|---|---|---|---|
| **Day 1 Visit (Pre-boost)** | | | | | | |
| N (non-missing) | 21 | 31 | 26 | 24 | 25 | 25 |
| Median ($P_{25}$, $P_{75}$) | 34.21 (14.01-70.53) | 34.94 (16.44-65.31) | 357.54 (176.25-758.00) | 333.39 (159.29-705.12) | 131.25 (84.72-175.40) | 80.34 (42.99-156.61) |
| Minimum - Maximum | 5.00-6566.30 | 5.00-548.18 | 104.33-4851.22 | 40.88-1614.17 | 5.00-2794.06 | 5.00-1091.12 |
| Geometric Mean (95% CI) | 38.55 (17.92-82.94) | 35.67 (24.66-51.60) | 381.82 (265.23-549.68) | 350.21 (229.06-535.43) | 129.82 (81.33-207.21) | 80.84 (51.22-127.60) |
| **Day 15 Visit (14 days post-boost)** | | | | | | |
| N (non-missing) | 21 | 31 | 26 | 24 | 25 | 25 |
| Median ($P_{25}$, $P_{75}$) | 3277.00 (1553.04-5285.23) | 2939.81 (1031.32-4954.53) | 3582.81 (2992.16-4835.23) | 3549.89 (2398.81-4368.78) | 3860.59 (3532.00-4998.14) | 2386.42 (1044.76-3956.27) |
| Minimum - Maximum | 563.83-29781.84 | 801.02-17623.19 | 1094.25-19433.45 | 686.48-27221.54 | 725.53-22439.32 | 489.42-15506.86 |
| Geometric Mean (95% CI) | 3126.46 (1966.74-4970.03) | 2588.95 (1842.09-3638.61) | 3941.63 (3072.98-5055.83) | 3506.68 (2405.95-5111.00) | 4451.43 (3140.94-6308.68) | 2368.38 (1567.32-3578.86) |
| N* (non-missing pre- and post-boost) | 21 | 31 | 26 | 24 | 25 | 25 |
| Participants with ≥ 2-fold rise[2], 95% CI | 100.0% (83.9%-100.0%) | 100.0% (88.8%-100.0%) | 100.0% (86.8%-100.0%) | 91.7% (73.0%-99.0%) | 100.0% (86.3%-100.0%) | 100.0% (86.3%-100.0%) |
| Participants with ≥4-fold rise[2], 95% CI | 100.0% (83.9%-100.0%) | 100.0% (88.8%-100.0%) | 92.3% (74.9%-99.1%) | 79.2% (57.8%-92.9%) | 100.0% (86.3%-100.0%) | 100.0% (86.3%-100.0%) |
| Geometric Mean Fold Rise[2], 95% CI | 81.11 (42.41-155.14) | 72.57 (51.04-103.19) | 10.32 (7.79-13.68) | 10.01 (6.69-14.99) | 34.29 (22.57-52.11) | 29.30 (19.25-44.58) |
| **Day 29 Visit (28 days post-boost)** | | | | | | |
| N (non-missing) | 18 | 25 | 25 | 23 | 21 | 22 |
| Median ($P_{25}$, $P_{75}$) | 2476.73 (899.00-3246.43) | 1844.80 (768.80-3293.42) | 3168.81 (1887.40-4374.95) | 2998.95 (1569.76-4034.49) | 3274.25 (1167.93-4753.28) | 1750.48 (669.79-3611.48) |
| Minimum - Maximum | 630.45-13181.15 | 203.86-11880.17 | 492.07-15673.64 | 979.32-14150.83 | 477.62-9245.19 | 354.88-10763.42 |
| Geometric Mean (95% CI) | 2104.38 (1347.53-3286.31) | 1583.49 (1056.09-2374.28) | 2858.71 (2113.84-3866.06) | 2929.94 (2150.22-3992.40) | 2539.37 (1735.52-3715.57) | 1668.64 (1062.20-2621.30) |
| N* (non-missing pre- and post-boost) | 18 | 24 | 25 | 22 | 21 | 22 |

| | Group 1E [Dosed Janssen, Boost Moderna] Age 18-55 yo (N=21) | Group 1E [Dosed Janssen, Boost Moderna] Age ≥56 yo (N=32) | Group 2E [Dosed Moderna, Boost Moderna] Age 18-55 yo (N=26) | Group 2E [Dosed Moderna, Boost Moderna] Age ≥56 yo (N=25) | Group 3E [Dosed Pfizer, Boost Moderna] Age 18-55 yo (N=25) | Group 3E [Dosed Pfizer, Boost Moderna] Age ≥56 yo (N=25) |
|---|---|---|---|---|---|---|
| Participants with ≥ 2-fold rise[2], 95% CI | 100.0% (81.5%-100.0%) | 95.8% (78.9%-99.9%) | 96.0% (79.6%-99.9%) | 95.5% (77.2%-99.9%) | 100.0% (83.9%-100.0%) | 100.0% (84.6%-100.0%) |
| Participants with ≥4-fold rise[2], 95% CI | 88.9% (65.3%-98.6%) | 95.8% (78.9%-99.9%) | 80.0% (59.3%-93.2%) | 81.8% (59.7%-94.8%) | 100.0% (83.9%-100.0%) | 100.0% (84.6%-100.0%) |
| Geometric Mean Fold Rise[2], 95% CI | 48.04 (22.35-103.23) | 42.94 (25.12-73.40) | 7.16 (5.30-9.67) | 7.81 (5.35-11.42) | 21.74 (12.80-36.92) | 20.21 (13.82-29.55) |

[1] Values below the lower limit of detection (LLOD = 10) were assigned the value of LLOD/2. Values between the LLOD and the lower limit of quantification (LLOQ = 18.5) are taken as reported, or a value of LLOQ/2 is assigned if observations are reported as <LLOQ but no value is provided. Values greater than the upper limit of quantification (ULOQ = 45118) are taken as reported, or a ceiling value equivalent to the ULOQ is assigned if values are not provided.

Supplemental Table 54: **SARS-CoV-2 Pseudovirion Neutralization Antibody –** Ad26.COV2.S Neutralization Antibodies Titer (ID50) to Pseudovirus D614G[1], by Group and Timepoint

| | Group 4E [Dosed Janssen, Boost Janssen] (N=50) | Group 5E [Dosed Moderna, Boost Janssen] (N=49) | Group 6E [Dosed Pfizer, Boost Janssen] (N=51) |
|---|---|---|---|
| **Day 1 Visit (Pre-boost)** | | | |
| N (non-missing) | 50 | 49 | 51 |
| Positive Response(%)[2] | 39 ( 78.0%) | 49 (100.0%) | 49 ( 96.1%) |
| Median ($P_{25}$, $P_{75}$) | 30.37 (11.47-56.81) | 243.83 (160.05-450.37) | 80.23 (36.03-142.48) |
| Minimum - Maximum | 5.00-9469.57 | 22.27-2567.26 | 5.00-2355.09 |
| Geometric Mean (95% CI) | 31.27 (20.13-48.57) | 254.91 (185.86-349.62) | 76.63 (55.25-106.28) |
| **Day 15 Visit (14 days post-boost)** | | | |
| N (non-missing) | 50 | 49 | 50 |
| Positive Response(%)[2] | 49 ( 98.0%) | 49 (100.0%) | 50 (100.0%) |
| Median ($P_{25}$, $P_{75}$) | 125.87 (79.84-167.91) | 1593.09 (759.77-2917.69) | 804.96 (409.14-1984.84) |
| Minimum - Maximum | 5.00-10632.92 | 187.18-17260.72 | 120.25-21574.07 |
| Geometric Mean (95% CI) | 129.85 (92.05-183.17) | 1578.80 (1200.37-2076.54) | 894.14 (652.14-1225.94) |
| N* (non-missing pre- and post-boost) | 50 | 49 | 50 |
| Participants with ≥ 2-fold rise[3], 95% CI | 72.0% (57.5%-83.8%) | 81.6% (68.0%-91.2%) | 94.0% (83.5%-98.7%) |
| Participants with ≥ 4-fold rise[3], 95% CI | 50.0% (35.5%-64.5%) | 61.2% (46.2%-74.8%) | 82.0% (68.6%-91.4%) |
| Geometric Mean Fold Rise[3], 95% CI | 4.15 (2.97-5.80) | 6.19 (4.49-8.54) | 12.50 (8.74-17.87) |

[1] Values below the lower limit of detection (LLOD = 10) were assigned the value of LLOD/2. Values between the LLOD and the lower limit of quantification (LLOQ = 18.5) are taken as reported, or a value of LLOQ/2 is assigned if observations are reported as <LLOQ but no value is provided. Values greater than the upper limit of quantification (ULOQ = 45118) are taken as reported, or a ceiling value equivalent to the ULOQ is assigned if values are not provided.

[2] Positive response is defined as an ID titer above the LLOD

[3] Relative to pre-vaccination (Day 1 Visit) levels, among participants with no-missing observations at both pre- and post-boost timepoints.

**Supplemental Table 55:** **SARS-CoV-2 Pseudovirion Neutralization Antibody** – Ad26.COV2.S Neutralization Antibodies Titer (ID50) to Pseudovirus D614G[1], by Group, Age Group and Timepoint

| | Group 4E [Dosed Janssen, Boost Janssen] Age 18-55 yo (N=26) | Group 4E [Dosed Janssen, Boost Janssen] Age ≥56 yo (N=24) | Group 5E [Dosed Moderna, Boost Janssen] Age 18-55 yo (N=24) | Group 5E [Dosed Moderna, Boost Janssen] Age ≥56 yo (N=25) | Group 6E [Dosed Pfizer, Boost Janssen] Age 18-55 yo (N=25) | Group 6E [Dosed Pfizer, Boost Janssen] Age ≥56 yo (N=26) |
|---|---|---|---|---|---|---|
| **Day 1 Visit (Pre-boost)** | | | | | | |
| N (non-missing) | 26 | 24 | 24 | 25 | 25 | 26 |
| Positive Response(%)[2] | 21 ( 80.8%) | 18 ( 75.0%) | 24 (100.0%) | 25 (100.0%) | 25 (100.0%) | 24 ( 92.3%) |
| Median ($P_{25}$, $P_{75}$) | 29.37 (11.47-46.44) | 31.32 (8.86-84.68) | 309.43 (173.48-631.48) | 206.06 (145.45-380.80) | 93.99 (64.79-161.28) | 62.04 (32.42-123.20) |
| Minimum - Maximum | 5.00-3447.06 | 5.00-9469.57 | 32.18-2567.26 | 22.27-1689.90 | 14.79-2355.09 | 5.00-857.55 |
| Geometric Mean (95% CI) | 28.42 (16.30-49.55) | 34.67 (16.61-72.38) | 310.10 (198.76-483.80) | 211.19 (132.37-336.96) | 100.05 (64.85-154.37) | 59.30 (36.12-97.34) |
| **Day 15 Visit (14 days post-boost)** | | | | | | |
| N (non-missing) | 26 | 24 | 24 | 25 | 24 | 26 |
| Positive Response(%)[2] | 26 (100.0%) | 23 ( 95.8%) | 24 (100.0%) | 25 (100.0%) | 24 (100.0%) | 26 (100.0%) |
| Median ($P_{25}$, $P_{75}$) | 131.51 (70.54-169.44) | 120.82 (99.02-165.09) | 1897.44 (782.28-3889.67) | 1128.83 (716.55-2839.82) | 1408.78 (457.37-2195.11) | 685.02 (409.14-1817.36) |
| Minimum - Maximum | 26.05-4970.24 | 5.00-10632.92 | 401.12-17260.72 | 187.18-5222.63 | 120.25-21574.07 | 144.83-4332.95 |
| Geometric Mean (95% CI) | 119.66 (76.19-187.92) | 141.87 (81.17-247.99) | 1931.94 (1263.71-2953.53) | 1300.68 (904.98-1869.38) | 1054.66 (623.92-1782.77) | 767.74 (518.44-1136.91) |
| N* (non-missing pre- and post-boost) | 26 | 24 | 24 | 25 | 24 | 26 |
| Participants with ≥ 2-fold rise[3], 95% CI | 76.9% (56.4%-91.0%) | 66.7% (44.7%-84.4%) | 87.5% (67.6%-97.3%) | 76.0% (54.9%-90.6%) | 95.8% (78.9%-99.9%) | 92.3% (74.9%-99.1%) |
| Participants with ≥4-fold rise[3], 95% CI | 61.5% (40.6%-79.8%) | 37.5% (18.8%-59.4%) | 58.3% (36.6%-77.9%) | 64.0% (42.5%-82.0%) | 83.3% (62.6%-95.3%) | 80.8% (60.6%-93.4%) |
| Geometric Mean Fold Rise[3], 95% CI | 4.21 (2.88-6.16) | 4.09 (2.26-7.41) | 6.23 (3.76-10.33) | 6.16 (3.98-9.54) | 12.02 (7.14-20.24) | 12.95 (7.64-21.93) |

[1] Values below the lower limit of detection (LLOD = 10) were assigned the value of LLOD/2. Values between the LLOD and the lower limit of quantification (LLOQ = 18.5) are taken as reported, or a value of LLOQ/2 is assigned if observations are reported as <LLOQ but no value is provided. Values greater than the upper limit of quantification (ULOQ = 45118) are taken as reported, or a ceiling value equivalent to the ULOQ is assigned if values are not provided.

[2] Positive response is defined as an ID titer above the LLOD

[3] Relative to pre-vaccination (Day 1 Visit) levels, among participants with no-missing observations at both pre- and post-boost timepoints.

Supplemental Table 56: **SARS-CoV-2 Pseudovirion Neutralization Antibody –** BNT162b2 Neutralization Antibodies Titer (ID50) to Pseudovirus D614G[1], by Group and Timepoint

| | Group 7E [Dosed Janssen, Boost Pfizer] (N=53) | Group 8E [Dosed Moderna, Boost Pfizer] (N=51) | Group 9E [Dosed Pfizer, Boost Pfizer] (N=50) |
|---|---|---|---|
| **Day 1 Visit (Pre-boost)** | | | |
| N (non-missing) | 53 | 51 | 50 |
| Positive Response(%)[2] | 45 ( 84.9%) | 51 (100.0%) | 49 ( 98.0%) |
| Median ($P_{25}$, $P_{75}$) | 33.39 (21.49-59.68) | 226.71 (136.36-500.42) | 95.47 (33.33-158.55) |
| Minimum - Maximum | 5.00-4597.42 | 29.04-976.88 | 5.00-2043.94 |
| Geometric Mean (95% CI) | 38.72 (26.61-56.35) | 238.04 (186.10-304.48) | 88.50 (63.17-123.98) |
| **Day 15 Visit (14 days post-boost)** | | | |
| N (non-missing) | 50 | 48 | 48 |
| Positive Response(%)[2] | 49 ( 98.0%) | 48 (100.0%) | 48 (100.0%) |
| Median ($P_{25}$, $P_{75}$) | 1462.26 (825.44-2597.30) | 3116.24 (1723.93-4584.87) | 1923.43 (930.12-3561.90) |
| Minimum - Maximum | 5.00-23208.91 | 441.76-7723.13 | 152.53-12632.42 |
| Geometric Mean (95% CI) | 1410.49 (990.01-2009.55) | 2801.01 (2311.73-3393.85) | 1845.64 (1406.07-2422.64) |
| N* (non-missing pre- and post-boost) | 50 | 48 | 48 |
| Participants with ≥ 2-fold rise[3], 95% CI | 98.0% (89.4%-99.9%) | 100.0% (92.6%-100.0%) | 100.0% (92.6%-100.0%) |
| Participants with ≥ 4-fold rise[3], 95% CI | 98.0% (89.4%-99.9%) | 93.8% (82.8%-98.7%) | 97.9% (88.9%-99.9%) |
| Geometric Mean Fold Rise[3], 95% CI | 35.08 (23.86-51.57) | 11.52 (8.96-14.82) | 19.98 (14.55-27.44) |

[1] Values below the lower limit of detection (LLOD = 10) were assigned the value of LLOD/2. Values between the LLOD and the lower limit of quantification (LLOQ = 18.5) are taken as reported, or a value of LLOQ/2 is assigned if observations are reported as <LLOQ but no value is provided. Values greater than the upper limit of quantification (ULOQ = 45118) are taken as reported, or a ceiling value equivalent to the ULOQ is assigned if values are not provided.
[2] Positive response is defined as an ID titer above the LLOD
[3] Relative to pre-vaccination (Day 1 Visit) levels, among participants with no-missing observations at both pre- and post-boost timepoints.

Supplemental Table 57: **SARS-CoV-2 Pseudovirion Neutralization Antibody** – BNT162b2 Neutralization Antibodies Titer (ID50) to Pseudovirus D614G[1], by Group, Age Group and Timepoint

| | Group 7E [Dosed Janssen, Boost Pfizer] Age 18-55 yo (N=21) | Group 7E [Dosed Janssen, Boost Pfizer] Age ≥56 yo (N=32) | Group 8E [Dosed Moderna, Boost Pfizer] Age 18-55 yo (N=26) | Group 8E [Dosed Moderna, Boost Pfizer] Age ≥56 yo (N=25) | Group 9E [Dosed Pfizer, Boost Pfizer] Age 18-55 yo (N=25) | Group 9E [Dosed Pfizer, Boost Pfizer] Age ≥56 yo (N=25) |
|---|---|---|---|---|---|---|
| **Day 1 Visit (Pre-boost)** | | | | | | |
| N (non-missing) | 31 | 22 | 22 | 29 | 24 | 26 |
| Positive Response(%)[2] | 24 ( 77.4%) | 21 ( 95.5%) | 22 (100.0%) | 29 (100.0%) | 24 (100.0%) | 25 ( 96.2%) |
| Median ($P_{25}$, $P_{75}$) | 27.64 (14.44-66.99) | 34.53 (25.56-56.81) | 419.90 (209.77-602.32) | 164.70 (104.70-346.33) | 105.32 (60.82-305.50) | 80.38 (25.60-137.60) |
| Minimum - Maximum | 5.00-4597.42 | 5.00-301.14 | 127.40-779.92 | 29.04-976.88 | 15.26-765.60 | 5.00-2043.94 |
| Geometric Mean (95% CI) | 36.79 (20.38-66.41) | 41.62 (27.50-62.98) | 349.09 (268.04-454.66) | 178.03 (124.12-255.34) | 122.14 (77.40-192.75) | 65.73 (40.13-107.67) |
| **Day 15 Visit (14 days post-boost)** | | | | | | |
| N (non-missing) | 28 | 22 | 20 | 28 | 23 | 25 |
| Positive Response(%)[2] | 27 ( 96.4%) | 22 (100.0%) | 20 (100.0%) | 28 (100.0%) | 23 (100.0%) | 25 (100.0%) |
| Median ($P_{25}$, $P_{75}$) | 1351.52 (825.85-2980.18) | 1462.26 (821.91-2597.30) | 4096.66 (2490.19-5319.46) | 2643.74 (1356.82-4150.40) | 2936.64 (712.41-3715.54) | 1511.55 (1012.10-3246.64) |
| Minimum - Maximum | 5.00-23208.91 | 165.77-19195.68 | 1132.64-7723.13 | 441.76-7183.68 | 411.58-12632.42 | 152.53-6155.03 |
| Geometric Mean (95% CI) | 1353.17 (782.49-2340.05) | 1486.97 (944.19-2341.75) | 3573.05 (2740.53-4658.47) | 2353.96 (1809.79-3061.74) | 2075.37 (1323.03-3255.54) | 1656.82 (1173.04-2340.13) |
| N* (non-missing pre- and post-boost) | 28 | 22 | 20 | 28 | 23 | 25 |
| Participants with ≥ 2-fold rise[3], 95% CI | 96.4% (81.7%-99.9%) | 100.0% (84.6%-100.0%) | 100.0% (83.2%-100.0%) | 100.0% (87.7%-100.0%) | 100.0% (85.2%-100.0%) | 100.0% (86.3%-100.0%) |
| Participants with ≥4-fold rise[3], 95% CI | 96.4% (81.7%-99.9%) | 100.0% (84.6%-100.0%) | 90.0% (68.3%-98.8%) | 96.4% (81.7%-99.9%) | 100.0% (85.2%-100.0%) | 96.0% (79.6%-99.9%) |
| Geometric Mean Fold Rise[3], 95% CI | 34.58 (18.47-64.74) | 35.73 (23.33-54.72) | 9.76 (7.17-13.29) | 12.97 (8.86-19.00) | 16.21 (10.01-26.23) | 24.22 (15.64-37.48) |

[1] Values below the lower limit of detection (LLOD = 10) were assigned the value of LLOD/2. Values between the LLOD and the lower limit of quantification (LLOQ = 18.5) are taken as reported, or a value of LLOQ/2 is assigned if observations are reported as <LLOQ but no value is provided. Values greater than the upper limit of quantification (ULOQ = 45118) are taken as reported, or a ceiling value equivalent to the ULOQ is assigned if values are not provided.

[2] Positive response is defined as an ID titer above the LLOD

[3] Relative to pre-vaccination (Day 1 Visit) levels, among participants with no-missing observations at both pre- and post-boost timepoints.

Supplemental Table 58: **SARS-CoV-2 Pseudovirion Neutralization Antibody –** mRNA-1273 Neutralization Antibodies Titer (ID80) to Pseudovirus D614G[1], by Group and Timepoint

| | Group 1E [Dosed Janssen, Boost Moderna] (N=53) | Group 2E [Dosed Moderna, Boost Moderna] (N=51) | Group 3E [Dosed Pfizer, Boost Moderna] (N=50) |
|---|---|---|---|
| **Day 1 Visit (Pre-boost)** | | | |
| N (non-missing) | 52 | 50 | 50 |
| Median ($P_{25}$, $P_{75}$) | 13.50 (5.00-19.84) | 107.09 (65.93-278.25) | 48.26 (17.60-67.56) |
| Minimum - Maximum | 5.00-1149.69 | 17.33-1495.67 | 5.00-645.11 |
| Geometric Mean (95% CI) | 13.32 (9.78-18.15) | 121.44 (93.26-158.14) | 36.10 (26.39-49.38) |
| **Day 15 Visit (14 days post-boost)** | | | |
| N (non-missing) | 52 | 50 | 50 |
| Median ($P_{25}$, $P_{75}$) | 1438.22 (426.60-1856.37) | 1597.61 (903.40-1878.56) | 1414.67 (519.06-2063.64) |
| Minimum - Maximum | 239.89-7164.69 | 332.46-9392.10 | 163.29-9360.99 |
| Geometric Mean (95% CI) | 1026.18 (795.82-1323.24) | 1419.65 (1148.93-1754.16) | 1194.90 (903.92-1579.56) |
| N* (non-missing pre- and post-boost) | 52 | 50 | 50 |
| Participants with ≥ 2-fold rise[2], 95% CI | 100.0% (93.2%-100.0%) | 98.0% (89.4%-99.9%) | 100.0% (92.9%-100.0%) |
| Participants with ≥ 4-fold rise[2], 95% CI | 100.0% (93.2%-100.0%) | 94.0% (83.5%-98.7%) | 100.0% (92.9%-100.0%) |
| Geometric Mean Fold Rise[2], 95% CI | 77.02 (57.05-104.00) | 11.69 (9.31-14.67) | 33.10 (25.50-42.96) |
| **Day 29 Visit (28 days post-boost)** | | | |
| N (non-missing) | 43 | 48 | 43 |
| Median ($P_{25}$, $P_{75}$) | 571.48 (351.13-1004.77) | 1012.49 (542.49-1633.83) | 590.80 (364.43-1634.37) |
| Minimum - Maximum | 85.07-3061.70 | 262.33-3818.29 | 109.95-3479.10 |
| Geometric Mean (95% CI) | 580.67 (447.70-753.13) | 961.10 (786.51-1174.44) | 730.79 (544.36-981.07) |
| N* (non-missing pre- and post-boost) | 42 | 47 | 43 |
| Participants with ≥ 2-fold rise[2], 95% CI | 97.6% (87.4%-99.9%) | 95.7% (85.5%-99.5%) | 100.0% (91.8%-100.0%) |
| Participants with ≥ 4-fold rise[2], 95% CI | 95.2% (83.8%-99.4%) | 80.9% (66.7%-90.9%) | 100.0% (91.8%-100.0%) |
| Geometric Mean Fold Rise[2], 95% CI | 41.27 (28.81-59.12) | 7.58 (6.08-9.44) | 21.29 (15.90-28.49) |

[1] Values below the lower limit of detection (LLOD = 10) were assigned the value of LLOD/2. Values between the LLOD and the lower limit of quantification (LLOQ = 14.3) are taken as reported, or a value of LLOQ/2 is assigned if observations are reported as <LLOQ but no value is provided. Values greater than the upper limit of quantification (ULOQ = 10232) are taken as reported, or a ceiling value equivalent to the ULOQ is assigned if values are not provided.

**Supplemental Table 59: SARS-CoV-2 Pseudovirion Neutralization Antibody** – mRNA-1273 Neutralization Antibodies Titer (ID80) to Pseudovirus D614G[1], by Group, Age Group and Timepoint

| | Group 1E [Dosed Janssen, Boost Moderna] Age 18-55 yo (N=21) | Group 1E [Dosed Janssen, Boost Moderna] Age ≥56 yo (N=32) | Group 2E [Dosed Moderna, Boost Moderna] Age 18-55 yo (N=26) | Group 2E [Dosed Moderna, Boost Moderna] Age ≥56 yo (N=25) | Group 3E [Dosed Pfizer, Boost Moderna] Age 18-55 yo (N=25) | Group 3E [Dosed Pfizer, Boost Moderna] Age ≥56 yo (N=25) |
|---|---|---|---|---|---|---|
| **Day 1 Visit (Pre-boost)** | | | | | | |
| N (non-missing) | 21 | 31 | 26 | 24 | 25 | 25 |
| Median ($P_{25}$, $P_{75}$) | 13.99 (5.00-20.59) | 13.15 (5.00-19.08) | 104.25 (74.78-278.25) | 109.05 (59.10-253.48) | 53.05 (26.55-64.68) | 24.51 (14.53-67.56) |
| Minimum - Maximum | 5.00-1149.69 | 5.00-127.95 | 40.32-1495.67 | 17.33-457.89 | 5.00-645.11 | 5.00-349.07 |
| Geometric Mean (95% CI) | 15.99 (8.81-29.01) | 11.77 (8.29-16.72) | 130.58 (91.58-186.18) | 112.26 (73.77-170.82) | 48.79 (32.34-73.61) | 26.72 (16.69-42.76) |
| **Day 15 Visit (14 days post-boost)** | | | | | | |
| N (non-missing) | 21 | 31 | 26 | 24 | 25 | 25 |
| Median ($P_{25}$, $P_{75}$) | 1512.93 (477.40-1840.07) | 939.09 (403.75-1872.68) | 1638.81 (1128.53-1963.05) | 1461.35 (726.85-1763.28) | 1710.19 (1293.70-2144.71) | 655.93 (404.12-1677.17) |
| Minimum - Maximum | 239.89-7010.62 | 253.42-7164.69 | 389.23-5131.04 | 332.46-9392.10 | 305.89-9360.99 | 163.29-7404.92 |
| Geometric Mean (95% CI) | 1144.01 (749.60-1745.96) | 953.34 (682.70-1331.27) | 1488.97 (1179.49-1879.64) | 1348.20 (921.14-1973.25) | 1703.50 (1192.56-2433.34) | 838.15 (559.27-1256.09) |
| N* (non-missing pre- and post-boost) | 21 | 31 | 26 | 24 | 25 | 25 |
| Participants with ≥ 2-fold rise[2], 95% CI | 100.0% (83.9%-100.0%) | 100.0% (88.8%-100.0%) | 100.0% (86.8%-100.0%) | 95.8% (78.9%-99.9%) | 100.0% (86.3%-100.0%) | 100.0% (86.3%-100.0%) |
| Participants with ≥ 4-fold rise[2], 95% CI | 100.0% (83.9%-100.0%) | 100.0% (88.8%-100.0%) | 96.2% (80.4%-99.9%) | 91.7% (73.0%-99.0%) | 100.0% (86.3%-100.0%) | 100.0% (86.3%-100.0%) |
| Geometric Mean Fold Rise[2], 95% CI | 71.56 (42.03-121.83) | 80.96 (55.50-118.11) | 11.40 (8.57-15.17) | 12.01 (8.20-17.59) | 34.92 (24.40-49.96) | 31.37 (20.93-47.02) |
| **Day 29 Visit (28 days post-boost)** | | | | | | |
| N (non-missing) | 18 | 25 | 25 | 23 | 21 | 22 |
| Median ($P_{25}$, $P_{75}$) | 686.96 (376.94-876.81) | 542.17 (265.09-1037.43) | 993.25 (606.16-1744.46) | 1041.60 (473.88-1383.11) | 1326.03 (455.78-1786.17) | 503.64 (254.27-1396.40) |
| Minimum - Maximum | 145.95-3061.70 | 85.07-2556.80 | 262.33-3818.29 | 324.37-3687.32 | 155.07-3389.74 | 109.95-3479.10 |
| Geometric Mean (95% CI) | 666.74 (453.00-981.35) | 525.66 (363.64-759.88) | 989.89 (749.10-1308.08) | 930.74 (681.63-1270.89) | 949.45 (649.76-1387.37) | 569.22 (362.44-893.98) |
| N* (non-missing pre- and post-boost) | 18 | 24 | 25 | 22 | 21 | 22 |
| Participants with ≥ 2-fold rise[2], 95% CI | 100.0% (81.5%-100.0%) | 95.8% (78.9%-99.9%) | 96.0% (79.6%-99.9%) | 95.5% (77.2%-99.9%) | 100.0% (83.9%-100.0%) | 100.0% (84.6%-100.0%) |

| | Group 1E [Dosed Janssen, Boost Moderna] Age 18-55 yo (N=21) | Group 1E [Dosed Janssen, Boost Moderna] Age ≥56 yo (N=32) | Group 2E [Dosed Moderna, Boost Moderna] Age 18-55 yo (N=26) | Group 2E [Dosed Moderna, Boost Moderna] Age ≥56 yo (N=25) | Group 3E [Dosed Pfizer, Boost Moderna] Age 18-55 yo (N=25) | Group 3E [Dosed Pfizer, Boost Moderna] Age ≥56 yo (N=25) |
|---|---|---|---|---|---|---|
| Participants with ≥ 4-fold rise[2], 95% CI | 94.4% (72.7%-99.9%) | 95.8% (78.9%-99.9%) | 84.0% (63.9%-95.5%) | 77.3% (54.6%-92.2%) | 100.0% (83.9%-100.0%) | 100.0% (84.6%-100.0%) |
| Geometric Mean Fold Rise[2], 95% CI | 38.37 (21.15-69.60) | 43.59 (26.91-70.61) | 7.32 (5.47-9.78) | 7.88 (5.49-11.31) | 21.21 (13.27-33.90) | 21.36 (14.43-31.61) |

[1] Values below the lower limit of detection (LLOD = 10) were assigned the value of LLOD/2. Values between the LLOD and the lower limit of quantification (LLOQ = 14.3) are taken as reported, or a value of LLOQ/2 is assigned if observations are reported as <LLOQ but no value is provided. Values greater than the upper limit of quantification (ULOQ = 10232) are taken as reported, or a ceiling value equivalent to the ULOQ is assigned if values are not provided.

Supplemental Table 60: **SARS-CoV-2 Pseudovirion Neutralization Antibody –** Ad26.COV2.S Neutralization Antibodies Titer (ID80) to Pseudovirus D614G[1], by Group and Timepoint

| | Group 4E [Dosed Janssen, Boost Janssen] (N=50) | Group 5E [Dosed Moderna, Boost Janssen] (N=49) | Group 6E [Dosed Pfizer, Boost Janssen] (N=51) |
|---|---|---|---|
| **Day 1 Visit (Pre-boost)** | | | |
| N (non-missing) | 50 | 49 | 51 |
| Positive Response(%)[2] | 30 ( 60.0%) | 48 ( 98.0%) | 44 ( 86.3%) |
| Median ($P_{25}$, $P_{75}$) | 13.85 (5.00-17.96) | 80.38 (56.72-118.00) | 24.62 (15.42-53.70) |
| Minimum - Maximum | 5.00-1964.69 | 5.00-605.32 | 5.00-625.46 |
| Geometric Mean (95% CI) | 13.32 (9.48-18.71) | 78.75 (59.31-104.55) | 27.11 (20.16-36.44) |
| **Day 15 Visit (14 days post-boost)** | | | |
| N (non-missing) | 50 | 49 | 50 |
| Positive Response(%)[2] | 49 ( 98.0%) | 49 (100.0%) | 50 (100.0%) |
| Median ($P_{25}$, $P_{75}$) | 55.70 (25.06-74.13) | 481.23 (317.13-877.74) | 335.78 (113.05-600.30) |
| Minimum - Maximum | 5.00-1097.07 | 75.25-6388.80 | 62.31-5519.64 |
| Geometric Mean (95% CI) | 51.85 (38.83-69.23) | 529.54 (405.82-690.99) | 324.25 (240.12-437.87) |
| N* (non-missing pre- and post-boost) | 50 | 49 | 50 |
| Participants with ≥ 2-fold rise[3], 95% CI | 78.0% (64.0%-88.5%) | 85.7% (72.8%-94.1%) | 96.0% (86.3%-99.5%) |
| Participants with ≥ 4-fold rise[3], 95% CI | 48.0% (33.7%-62.6%) | 69.4% (54.6%-81.7%) | 90.0% (78.2%-96.7%) |
| Geometric Mean Fold Rise[3], 95% CI | 3.89 (2.99-5.06) | 6.72 (4.94-9.16) | 12.74 (9.28-17.47) |

[1] Values below the lower limit of detection (LLOD = 10) were assigned the value of LLOD/2. Values between the LLOD and the lower limit of quantification (LLOQ = 14.3) are taken as reported, or a value of LLOQ/2 is assigned if observations are reported as <LLOQ but no value is provided. Values greater than the upper limit of quantification (ULOQ = 10232) are taken as reported, or a ceiling value equivalent to the ULOQ is assigned if values are not provided.

Supplemental Table 61: **SARS-CoV-2 Pseudovirion Neutralization Antibody -** Ad26.COV2.S Neutralization Antibodies Titer (ID80) to Pseudovirus D614G[1], by Group, Age Group and Timepoint

| | Group 4E [Dosed Janssen, Boost Janssen] Age 18-55 yo (N=26) | Group 4E [Dosed Janssen, Boost Janssen] Age ≥56 yo (N=24) | Group 5E [Dosed Moderna, Boost Janssen] Age 18-55 yo (N=24) | Group 5E [Dosed Moderna, Boost Janssen] Age ≥56 yo (N=25) | Group 6E [Dosed Pfizer, Boost Janssen] Age 18-55 yo (N=25) | Group 6E [Dosed Pfizer, Boost Janssen] Age ≥56 yo (N=26) |
|---|---|---|---|---|---|---|
| **Day 1 Visit (Pre-boost)** | | | | | | |
| N (non-missing) | 26 | 24 | 24 | 25 | 25 | 26 |
| Positive Response(%)[2] | 16 ( 61.5%) | 14 ( 58.3%) | 24 (100.0%) | 24 ( 96.0%) | 24 ( 96.0%) | 20 ( 76.9%) |
| Median ($P_{25}$, $P_{75}$) | 13.01 (5.00-16.69) | 14.65 (5.00-23.69) | 89.15 (61.01-157.56) | 80.38 (50.29-102.24) | 26.85 (20.11-68.65) | 20.79 (11.98-39.23) |
| Minimum - Maximum | 5.00-640.26 | 5.00-1964.69 | 15.25-605.32 | 5.00-418.82 | 5.00-625.46 | 5.00-227.09 |
| Geometric Mean (95% CI) | 12.18 (7.96-18.64) | 14.67 (8.30-25.95) | 94.86 (64.21-140.14) | 65.86 (43.05-100.75) | 36.13 (23.92-54.58) | 20.57 (13.48-31.37) |
| **Day 15 Visit (14 days post-boost)** | | | | | | |
| N (non-missing) | 26 | 24 | 24 | 25 | 24 | 26 |
| Positive Response(%)[2] | 26 (100.0%) | 23 ( 95.8%) | 24 (100.0%) | 25 (100.0%) | 24 (100.0%) | 26 (100.0%) |
| Median ($P_{25}$, $P_{75}$) | 58.18 (23.03-78.41) | 55.70 (26.81-73.60) | 555.01 (348.83-1528.45) | 405.00 (316.54-646.91) | 458.37 (159.42-626.31) | 296.84 (113.05-510.11) |
| Minimum - Maximum | 13.87-1063.08 | 5.00-1097.07 | 126.97-6388.80 | 75.25-2050.74 | 67.01-5519.64 | 62.31-1804.25 |
| Geometric Mean (95% CI) | 50.15 (33.42-75.27) | 53.75 (34.49-83.76) | 656.02 (431.85-996.56) | 431.13 (306.19-607.03) | 378.60 (234.73-610.65) | 281.04 (188.94-418.02) |
| N* (non-missing pre- and post-boost) | 26 | 24 | 24 | 25 | 24 | 26 |
| Participants with ≥ 2-fold rise[3], 95% CI | 84.6% (65.1%-95.6%) | 70.8% (48.9%-87.4%) | 91.7% (73.0%-99.0%) | 80.0% (59.3%-93.2%) | 95.8% (78.9%-99.9%) | 96.2% (80.4%-99.9%) |
| Participants with ≥ 4-fold rise[3], 95% CI | 46.2% (26.6%-66.6%) | 50.0% (29.1%-70.9%) | 66.7% (44.7%-84.4%) | 72.0% (50.6%-87.9%) | 91.7% (73.0%-99.0%) | 88.5% (69.8%-97.6%) |
| Geometric Mean Fold Rise[3], 95% CI | 4.12 (3.04-5.58) | 3.66 (2.31-5.81) | 6.92 (4.32-11.08) | 6.55 (4.23-10.13) | 11.80 (7.36-18.93) | 13.66 (8.68-21.52) |

[1] Values below the lower limit of detection (LLOD = 10) were assigned the value of LLOD/2. Values between the LLOD and the lower limit of quantification (LLOQ = 14.3) are taken as reported, or a value of LLOQ/2 is assigned if observations are reported as <LLOQ but no value is provided. Values greater than the upper limit of quantification (ULOQ = 10232) are taken as reported, or a ceiling value equivalent to the ULOQ is assigned if values are not provided.

Supplemental Table 62: **SARS-CoV-2 Pseudovirion Neutralization Antibody –** BNT162b2 Neutralization Antibodies Titer (ID80) to Pseudovirus D614G[1], by Group and Timepoint

| | **Group 7E [Dosed Janssen, Boost Pfizer] (N=53)** | **Group 8E [Dosed Moderna, Boost Pfizer] (N=51)** | **Group 9E [Dosed Pfizer, Boost Pfizer] (N=50)** |
|---|---|---|---|
| **Day 1 Visit (Pre-boost)** | | | |
| N (non-missing) | 53 | 51 | 50 |
| Positive Response(%)[2] | 40 ( 75.5%) | 51 (100.0%) | 45 ( 90.0%) |
| Median ($P_{25}$, $P_{75}$) | 14.69 (10.33-19.59) | 87.28 (55.65-134.94) | 27.77 (15.61-67.29) |
| Minimum - Maximum | 5.00-1858.15 | 14.68-324.85 | 5.00-560.12 |
| Geometric Mean (95% CI) | 16.23 (11.99-21.99) | 76.80 (60.85-96.92) | 31.60 (23.38-42.71) |
| **Day 15 Visit (14 days post-boost)** | | | |
| N (non-missing) | 50 | 48 | 48 |
| Positive Response(%)[2] | 49 ( 98.0%) | 48 (100.0%) | 48 (100.0%) |
| Median ($P_{25}$, $P_{75}$) | 482.06 (349.37-752.98) | 1183.28 (502.05-1767.69) | 531.99 (382.95-1491.06) |
| Minimum - Maximum | 5.00-6496.82 | 162.07-2164.58 | 74.85-3470.50 |
| Geometric Mean (95% CI) | 510.03 (373.41-696.63) | 967.81 (795.44-1177.53) | 673.97 (523.74-867.30) |
| N* (non-missing pre- and post-boost) | 50 | 48 | 48 |
| Participants with ≥ 2-fold rise[3], 95% CI | 98.0% (89.4%-99.9%) | 100.0% (92.6%-100.0%) | 100.0% (92.6%-100.0%) |
| Participants with ≥ 4-fold rise[3], 95% CI | 96.0% (86.3%-99.5%) | 93.8% (82.8%-98.7%) | 97.9% (88.9%-99.9%) |
| Geometric Mean Fold Rise[3], 95% CI | 30.02 (21.81-41.33) | 12.36 (9.95-15.35) | 20.63 (15.69-27.14) |

[1] Values below the lower limit of detection (LLOD = 10) were assigned the value of LLOD/2. Values between the LLOD and the lower limit of quantification (LLOQ = 14.3) are taken as reported, or a value of LLOQ/2 is assigned if observations are reported as <LLOQ but no value is provided. Values greater than the upper limit of quantification (ULOQ = 10232) are taken as reported, or a ceiling value equivalent to the ULOQ is assigned if values are not provided.

**Supplemental Table 63: SARS-CoV-2 Pseudovirion Neutralization Antibody** – BNT162b2 Neutralization Antibodies Titer (ID80) to Pseudovirus D614G[1], by Group, Age Group and Timepoint

| | Group 7E [Dosed Janssen, Boost Pfizer] Age 18-55 yo (N=21) | Group 7E [Dosed Janssen, Boost Pfizer] Age ≥56 yo (N=32) | Group 8E [Dosed Moderna, Boost Pfizer] Age 18-55 yo (N=26) | Group 8E [Dosed Moderna, Boost Pfizer] Age ≥56 yo (N=25) | Group 9E [Dosed Pfizer, Boost Pfizer] Age 18-55 yo (N=25) | Group 9E [Dosed Pfizer, Boost Pfizer] Age ≥56 yo (N=25) |
|---|---|---|---|---|---|---|
| **Day 1 Visit (Pre-boost)** | | | | | | |
| N (non-missing) | 31 | 22 | 22 | 29 | 24 | 26 |
| Positive Response(%)[2] | 22 ( 71.0%) | 18 ( 81.8%) | 22 (100.0%) | 29 (100.0%) | 23 ( 95.8%) | 22 ( 84.6%) |
| Median ($P_{25}$, $P_{75}$) | 14.69 (5.00-19.69) | 14.54 (10.62-19.59) | 118.86 (77.17-150.99) | 66.02 (26.59-96.85) | 35.33 (20.53-87.97) | 22.15 (12.54-62.07) |
| Minimum - Maximum | 5.00-1858.15 | 5.00-94.40 | 55.65-303.48 | 14.68-324.85 | 5.00-270.78 | 5.00-560.12 |
| Geometric Mean (95% CI) | 16.75 (10.44-26.87) | 15.53 (10.94-22.06) | 116.54 (93.64-145.03) | 55.97 (39.83-78.65) | 41.21 (27.34-62.12) | 24.73 (15.88-38.53) |
| **Day 15 Visit (14 days post-boost)** | | | | | | |
| N (non-missing) | 28 | 22 | 20 | 28 | 23 | 25 |
| Positive Response(%)[2] | 27 ( 96.4%) | 22 (100.0%) | 20 (100.0%) | 28 (100.0%) | 23 (100.0%) | 25 (100.0%) |
| Median ($P_{25}$, $P_{75}$) | 459.55 (373.20-1077.23) | 487.87 (349.37-752.98) | 1620.10 (752.91-1799.56) | 706.60 (449.71-1702.96) | 872.32 (341.65-1732.66) | 499.22 (403.03-1018.90) |
| Minimum - Maximum | 5.00-6496.82 | 74.13-5734.12 | 414.78-2164.58 | 162.07-2133.20 | 138.90-3470.50 | 74.85-2193.49 |
| Geometric Mean (95% CI) | 520.75 (322.23-841.58) | 496.69 (331.08-745.14) | 1233.06 (949.47-1601.35) | 814.05 (618.78-1070.94) | 776.90 (514.02-1174.21) | 591.37 (429.10-815.01) |
| N* (non-missing pre- and post-boost) | 28 | 22 | 20 | 28 | 23 | 25 |
| Participants with ≥ 2-fold rise[3], 95% CI | 96.4% (81.7%-99.9%) | 100.0% (84.6%-100.0%) | 100.0% (83.2%-100.0%) | 100.0% (87.7%-100.0%) | 100.0% (85.2%-100.0%) | 100.0% (86.3%-100.0%) |
| Participants with ≥ 4-fold rise[3], 95% CI | 92.9% (76.5%-99.1%) | 100.0% (84.6%-100.0%) | 85.0% (62.1%-96.8%) | 100.0% (87.7%-100.0%) | 100.0% (85.2%-100.0%) | 96.0% (79.6%-99.9%) |
| Geometric Mean Fold Rise[3], 95% CI | 28.57 (17.35-47.04) | 31.98 (21.41-47.75) | 10.15 (7.60-13.57) | 14.22 (10.41-19.44) | 18.14 (11.93-27.58) | 23.23 (15.87-34.02) |

[1] Values below the lower limit of detection (LLOD = 10) were assigned the value of LLOD/2. Values between the LLOD and the lower limit of quantification (LLOQ = 14.3) are taken as reported, or a value of LLOQ/2 is assigned if observations are reported as <LLOQ but no value is provided. Values greater than the upper limit of quantification (ULOQ = 10232) are taken as reported, or a ceiling value equivalent to the ULOQ is assigned if values are not provided.

Supplemental Table 64: **SARS-CoV-2 Pseudovirion Neutralization Antibody –** mRNA-1273 Neutralization Antibodies Titer (ID50) to Pseudovirus B.1.617.2 (Delta)[1], by Group and Timepoint

| | Group 1E [Dosed Janssen, Boost Moderna] (N=53) | Group 2E [Dosed Moderna, Boost Moderna] (N=51) | Group 3E [Dosed Pfizer, Boost Moderna] (N=50) |
|---|---|---|---|
| **Day 1 Visit (Pre-boost)** | | | |
| N (non-missing) | 20 | 20 | 20 |
| Median ($P_{25}$, $P_{75}$) | 10.31 (5.00-14.94) | 119.57 (44.00-179.43) | 32.34 (15.79-44.09) |
| Minimum - Maximum | 5.00-5475.92 | 12.18-671.41 | 5.00-375.35 |
| Geometric Mean (95% CI) | 13.80 (6.20-30.71) | 98.26 (59.15-163.22) | 27.34 (16.31-45.85) |
| **Day 15 Visit (14 days post-boost)** | | | |
| N (non-missing) | 20 | 20 | 20 |
| Median ($P_{25}$, $P_{75}$) | 846.27 (533.97-1363.28) | 1057.97 (564.98-1923.82) | 920.70 (425.96-2519.23) |
| Minimum - Maximum | 113.44-10563.34 | 292.72-4890.59 | 151.45-5081.50 |
| Geometric Mean (95% CI) | 789.54 (477.99-1304.15) | 1085.42 (768.10-1533.83) | 946.29 (567.17-1578.84) |
| N* (non-missing pre- and post-boost) | 20 | 20 | 20 |
| Participants with ≥ 2-fold rise[2], 95% CI | 90.0% (68.3%-98.8%) | 90.0% (68.3%-98.8%) | 100.0% (83.2%-100.0%) |
| Participants with ≥ 4-fold rise[2], 95% CI | 90.0% (68.3%-98.8%) | 80.0% (56.3%-94.3%) | 100.0% (83.2%-100.0%) |
| Geometric Mean Fold Rise[2], 95% CI | 57.23 (27.27-120.10) | 11.05 (6.70-18.21) | 34.61 (23.05-51.96) |
| **Day 29 Visit (28 days post-boost)** | | | |
| N (non-missing) | 18 | 20 | 19 |
| Median ($P_{25}$, $P_{75}$) | 587.90 (382.48-793.70) | 691.54 (463.35-1177.51) | 600.90 (240.87-1696.30) |
| Minimum - Maximum | 79.99-3723.96 | 163.11-3296.81 | 101.33-8113.99 |
| Geometric Mean (95% CI) | 542.79 (326.62-902.04) | 735.02 (507.00-1065.60) | 654.46 (370.54-1155.94) |
| N* (non-missing pre- and post-boost) | 18 | 20 | 19 |
| Participants with ≥ 2-fold rise[2], 95% CI | 88.9% (65.3%-98.6%) | 95.0% (75.1%-99.9%) | 100.0% (82.4%-100.0%) |
| Participants with ≥ 4-fold rise[2], 95% CI | 88.9% (65.3%-98.6%) | 70.0% (45.7%-88.1%) | 100.0% (82.4%-100.0%) |
| Geometric Mean Fold Rise[2], 95% CI | 35.15 (14.69-84.06) | 7.48 (4.90-11.43) | 24.40 (15.29-38.94) |

[1] Values below the lower limit of detection (LLOD = 10) were assigned the value of LLOD/2

[2] Relative to pre-vaccination (Day 1 Visit) levels, among participants with no-missing observations at both pre- and post-boost timepoints.

Supplemental Table 65: **SARS-CoV-2 Pseudovirion Neutralization Antibody –** mRNA-1273 Neutralization Antibodies Titer (ID50) to Pseudovirus B.1.617.2 (Delta)[1], by Group, Age Group and Timepoint

| | Group 1E [Dosed Janssen, Boost Moderna] Age 18-55 yo (N=21) | Group 1E [Dosed Janssen, Boost Moderna] Age ≥56 yo (N=32) | Group 2E [Dosed Moderna, Boost Moderna] Age 18-55 yo (N=26) | Group 2E [Dosed Moderna, Boost Moderna] Age ≥56 yo (N=25) | Group 3E [Dosed Pfizer, Boost Moderna] Age 18-55 yo (N=25) | Group 3E [Dosed Pfizer, Boost Moderna] Age ≥56 yo (N=25) |
|---|---|---|---|---|---|---|
| **Day 1 Visit (Pre-boost)** | | | | | | |
| N (non-missing) | 10 | 10 | 10 | 10 | 10 | 10 |
| Median ($P_{25}$, $P_{75}$) | 10.85 (5.00-16.03) | 7.56 (5.00-13.84) | 109.79 (46.23-154.39) | 156.98 (29.70-205.87) | 32.34 (28.13-54.52) | 27.15 (5.00-43.55) |
| Minimum - Maximum | 5.00-5475.92 | 5.00-286.40 | 12.18-671.41 | 18.77-671.32 | 12.19-375.35 | 5.00-105.68 |
| Geometric Mean (95% CI) | 17.65 (3.88-80.34) | 10.78 (4.43-26.27) | 96.26 (45.47-203.82) | 100.29 (43.26-232.53) | 41.40 (21.78-78.71) | 18.06 (7.78-41.92) |
| **Day 15 Visit (14 days post-boost)** | | | | | | |
| N (non-missing) | 10 | 10 | 10 | 10 | 10 | 10 |
| Median ($P_{25}$, $P_{75}$) | 978.86 (850.29-1704.42) | 704.65 (191.86-842.25) | 773.57 (571.97-1263.33) | 1499.07 (543.96-1950.24) | 1832.92 (1046.55-4002.16) | 425.96 (246.09-794.85) |
| Minimum - Maximum | 113.44-10563.34 | 155.56-2555.61 | 452.44-2669.53 | 292.72-4890.59 | 702.90-5081.50 | 151.45-2635.40 |
| Geometric Mean (95% CI) | 1153.14 (517.16-2571.20) | 540.59 (279.32-1046.26) | 945.66 (606.16-1475.31) | 1245.83 (677.87-2289.65) | 1893.80 (1100.41-3259.22) | 472.84 (241.71-924.99) |
| N* (non-missing pre- and post-boost) | 10 | 10 | 10 | 10 | 10 | 10 |
| Participants with ≥ 2-fold rise[2], 95% CI | 90.0% (55.5%-99.7%) | 90.0% (55.5%-99.7%) | 90.0% (55.5%-99.7%) | 90.0% (55.5%-99.7%) | 100.0% (69.2%-100.0%) | 100.0% (69.2%-100.0%) |
| Participants with ≥4-fold rise[2], 95% CI | 90.0% (55.5%-99.7%) | 90.0% (55.5%-99.7%) | 80.0% (44.4%-97.5%) | 80.0% (44.4%-97.5%) | 100.0% (69.2%-100.0%) | 100.0% (69.2%-100.0%) |
| Geometric Mean Fold Rise[2], 95% CI | 65.33 (22.52-189.55) | 50.13 (14.40-174.51) | 9.82 (4.42-21.82) | 12.42 (5.80-26.62) | 45.74 (24.08-86.88) | 26.19 (14.95-45.87) |
| **Day 29 Visit (28 days post-boost)** | | | | | | |
| N (non-missing) | 9 | 9 | 10 | 10 | 9 | 10 |
| Median ($P_{25}$, $P_{75}$) | 734.60 (509.46-955.80) | 459.77 (382.48-700.72) | 691.54 (601.01-957.26) | 701.61 (336.11-1388.36) | 1147.44 (647.20-2424.42) | 286.01 (173.96-595.16) |
| Minimum - Maximum | 89.10-3723.96 | 79.99-2568.75 | 226.40-2668.49 | 163.11-3296.81 | 532.21-8113.99 | 101.33-1696.30 |
| Geometric Mean (95% CI) | 667.63 (302.37-1474.13) | 441.29 (200.10-973.22) | 735.14 (465.54-1160.85) | 734.91 (369.82-1460.42) | 1445.30 (695.73-3002.42) | 320.79 (169.92-605.63) |
| N* (non-missing pre- and post-boost) | 9 | 9 | 10 | 10 | 9 | 10 |
| Participants with ≥ 2-fold rise[2], 95% CI | 88.9% (51.8%-99.7%) | 88.9% (51.8%-99.7%) | 90.0% (55.5%-99.7%) | 100.0% (69.2%-100.0%) | 100.0% (66.4%-100.0%) | 100.0% (69.2%-100.0%) |
| Participants with ≥4-fold rise[2], 95% CI | 88.9% (51.8%-99.7%) | 88.9% (51.8%-99.7%) | 70.0% (34.8%-93.3%) | 70.0% (34.8%-93.3%) | 100.0% (66.4%-100.0%) | 100.0% (69.2%-100.0%) |
| Geometric Mean Fold Rise[2], 95% CI | 32.88 (9.80-110.28) | 37.57 (8.01-176.31) | 7.64 (3.57-16.35) | 7.33 (4.21-12.75) | 34.71 (14.94-80.63) | 17.77 (10.32-30.57) |

[1] Values below the lower limit of detection (LLOD = 10) were assigned the value of LLOD/2

[2] Relative to pre-vaccination (Day 1 Visit) levels, among participants with no-missing observations at both pre- and post-boost timepoints.

Supplemental Table 66: **SARS-CoV-2 Pseudovirion Neutralization Antibody –** mRNA-1273 Neutralization Antibodies Titer (ID80) to Pseudovirus B.1.617.2 (Delta)[1], by Group and Timepoint

| | Group 1E [Dosed Janssen, Boost Moderna] (N=53) | Group 2E [Dosed Moderna, Boost Moderna] (N=51) | Group 3E [Dosed Pfizer, Boost Moderna] (N=50) |
|---|---|---|---|
| **Day 1 Visit (Pre-boost)** | | | |
| N (non-missing) | 20 | 20 | 20 |
| Median ($P_{25}$, $P_{75}$) | 5.00 (5.00-5.00) | 31.99 (16.65-70.54) | 15.54 (5.00-16.09) |
| Minimum - Maximum | 5.00-430.95 | 10.18-246.45 | 5.00-92.89 |
| Geometric Mean (95% CI) | 7.03 (4.32-11.45) | 34.58 (22.82-52.38) | 12.24 (8.70-17.22) |
| **Day 15 Visit (14 days post-boost)** | | | |
| N (non-missing) | 20 | 20 | 20 |
| Median ($P_{25}$, $P_{75}$) | 385.49 (192.79-459.97) | 383.40 (222.03-551.80) | 378.45 (140.12-722.76) |
| Minimum - Maximum | 67.73-1151.46 | 98.54-1941.47 | 58.67-1954.70 |
| Geometric Mean (95% CI) | 293.43 (200.29-429.89) | 362.49 (263.16-499.33) | 346.42 (210.56-569.95) |
| N* (non-missing pre- and post-boost) | 20 | 20 | 20 |
| Participants with ≥ 2-fold rise[2], 95% CI | 100.0% (83.2%-100.0%) | 95.0% (75.1%-99.9%) | 100.0% (83.2%-100.0%) |
| Participants with ≥ 4-fold rise[2], 95% CI | 90.0% (68.3%-98.8%) | 90.0% (68.3%-98.8%) | 100.0% (83.2%-100.0%) |
| Geometric Mean Fold Rise[2], 95% CI | 41.72 (24.43-71.22) | 10.48 (7.24-15.19) | 28.30 (20.42-39.21) |
| **Day 29 Visit (28 days post-boost)** | | | |
| N (non-missing) | 18 | 20 | 19 |
| Median ($P_{25}$, $P_{75}$) | 287.44 (127.74-370.77) | 306.60 (112.48-442.28) | 275.92 (89.90-525.23) |
| Minimum - Maximum | 23.59-597.39 | 79.66-1412.89 | 28.55-2020.17 |
| Geometric Mean (95% CI) | 211.56 (136.84-327.07) | 266.93 (183.54-388.20) | 222.35 (128.35-385.18) |
| N* (non-missing pre- and post-boost) | 18 | 20 | 19 |
| Participants with ≥ 2-fold rise[2], 95% CI | 88.9% (65.3%-98.6%) | 95.0% (75.1%-99.9%) | 100.0% (82.4%-100.0%) |
| Participants with ≥ 4-fold rise[2], 95% CI | 88.9% (65.3%-98.6%) | 90.0% (68.3%-98.8%) | 100.0% (82.4%-100.0%) |
| Geometric Mean Fold Rise[2], 95% CI | 28.96 (14.68-57.12) | 7.72 (5.45-10.94) | 18.41 (12.84-26.40) |

[1] Values below the lower limit of detection (LLOD = 10) were assigned the value of LLOD/2

[2] Relative to pre-vaccination (Day 1 Visit) levels, among participants with no-missing observations at both pre- and post-boost timepoints.

Supplemental Table 67: **SARS-CoV-2 Pseudovirion Neutralization Antibody –** mRNA-1273 Neutralization Antibodies Titer (ID50) to Pseudovirus B.1.617.2 (Delta)[1], by Group, Age Group and Timepoint

| | Group 1E [Dosed Janssen, Boost Moderna] Age 18-55 yo (N=21) | Group 1E [Dosed Janssen, Boost Moderna] Age ≥56 yo (N=32) | Group 2E [Dosed Moderna, Boost Moderna] Age 18-55 yo (N=26) | Group 2E [Dosed Moderna, Boost Moderna] Age ≥56 yo (N=25) | Group 3E [Dosed Pfizer, Boost Moderna] Age 18-55 yo (N=25) | Group 3E [Dosed Pfizer, Boost Moderna] Age ≥56 yo (N=25) |
|---|---|---|---|---|---|---|
| **Day 1 Visit (Pre-boost)** | | | | | | |
| N (non-missing) | 10 | 10 | 10 | 10 | 10 | 10 |
| Median ($P_{25}$, $P_{75}$) | 5.00 (5.00-5.00) | 5.00 (5.00-5.00) | 27.54 (16.99-39.19) | 41.99 (13.72-79.12) | 15.54 (12.65-19.02) | 10.36 (5.00-15.92) |
| Minimum - Maximum | 5.00-430.95 | 5.00-23.23 | 10.18-246.45 | 11.03-147.82 | 5.00-92.89 | 5.00-20.11 |
| Geometric Mean (95% CI) | 8.49 (3.11-23.18) | 5.83 (4.12-8.25) | 33.08 (17.29-63.30) | 36.14 (18.79-69.51) | 16.40 (9.77-27.53) | 9.14 (5.78-14.44) |
| **Day 15 Visit (14 days post-boost)** | | | | | | |
| N (non-missing) | 10 | 10 | 10 | 10 | 10 | 10 |
| Median ($P_{25}$, $P_{75}$) | 423.00 (371.54-566.07) | 295.61 (84.20-390.89) | 374.10 (268.74-412.62) | 459.14 (169.23-569.39) | 512.28 (395.90-1184.69) | 140.12 (93.47-342.47) |
| Minimum - Maximum | 67.73-1151.46 | 75.73-699.75 | 148.22-692.76 | 98.54-1941.47 | 351.36-1954.70 | 58.67-1278.71 |
| Geometric Mean (95% CI) | 393.37 (233.95-661.43) | 218.89 (120.98-396.02) | 347.15 (244.21-493.50) | 378.51 (204.35-701.13) | 676.81 (416.51-1099.78) | 177.31 (88.88-353.72) |
| N* (non-missing pre- and post-boost) | 10 | 10 | 10 | 10 | 10 | 10 |
| Participants with ≥ 2-fold rise[2], 95% CI | 100.0% (69.2%-100.0%) | 100.0% (69.2%-100.0%) | 90.0% (55.5%-99.7%) | 100.0% (69.2%-100.0%) | 100.0% (69.2%-100.0%) | 100.0% (69.2%-100.0%) |
| Participants with ≥ 4-fold rise[2], 95% CI | 90.0% (55.5%-99.7%) | 90.0% (55.5%-99.7%) | 90.0% (55.5%-99.7%) | 90.0% (55.5%-99.7%) | 100.0% (69.2%-100.0%) | 100.0% (69.2%-100.0%) |
| Geometric Mean Fold Rise[2], 95% CI | 46.35 (19.60-109.62) | 37.54 (16.69-84.48) | 10.49 (5.61-19.62) | 10.47 (6.14-17.88) | 41.27 (26.51-64.26) | 19.40 (12.90-29.19) |
| **Day 29 Visit (28 days post-boost)** | | | | | | |
| N (non-missing) | 9 | 9 | 10 | 10 | 9 | 10 |
| Median ($P_{25}$, $P_{75}$) | 303.55 (253.17-418.99) | 217.34 (116.41-345.26) | 304.18 (209.60-361.56) | 327.10 (97.82-482.69) | 359.08 (293.29-595.73) | 96.68 (64.51-275.92) |
| Minimum - Maximum | 55.58-597.39 | 23.59-515.71 | 85.55-635.72 | 79.66-1412.89 | 128.75-2020.17 | 28.55-525.23 |
| Geometric Mean (95% CI) | 271.31 (152.91-481.42) | 164.96 (78.44-346.90) | 261.35 (168.74-404.79) | 272.63 (134.43-552.91) | 464.59 (228.77-943.48) | 114.55 (60.11-218.29) |
| N* (non-missing pre- and post-boost) | 9 | 9 | 10 | 10 | 9 | 10 |
| Participants with ≥ 2-fold rise[2], 95% CI | 88.9% (51.8%-99.7%) | 88.9% (51.8%-99.7%) | 90.0% (55.5%-99.7%) | 100.0% (69.2%-100.0%) | 100.0% (66.4%-100.0%) | 100.0% (69.2%-100.0%) |
| Participants with ≥ 4-fold rise[2], 95% CI | 88.9% (51.8%-99.7%) | 88.9% (51.8%-99.7%) | 90.0% (55.5%-99.7%) | 90.0% (55.5%-99.7%) | 100.0% (66.4%-100.0%) | 100.0% (69.2%-100.0%) |
| Geometric Mean Fold Rise[2], 95% CI | 30.15 (10.12-89.81) | 27.82 (9.52-81.24) | 7.90 (4.22-14.79) | 7.54 (4.80-11.86) | 28.22 (16.24-49.05) | 12.54 (8.41-18.69) |

[1] Values below the lower limit of detection (LLOD = 10) were assigned the value of LLOD/2

[2] Relative to pre-vaccination (Day 1 Visit) levels, among participants with no-missing observations at both pre- and post-boost timepoints.

Supplemental Table 68: **SARS-CoV-2 Pseudovirion Neutralization Antibody –** mRNA-1273 Neutralization Antibodies Titer (ID50) to Pseudovirus B.1.351 (Beta)[1], by Group and Timepoint

| | Group 1E [Dosed Janssen, Boost Moderna] (N=53) | Group 2E [Dosed Moderna, Boost Moderna] (N=51) | Group 3E [Dosed Pfizer, Boost Moderna] (N=50) |
|---|---|---|---|
| **Day 1 Visit (Pre-boost)** | | | |
| N (non-missing) | 20 | 20 | 20 |
| Median ($P_{25}$, $P_{75}$) | 5.00 (5.00-13.33) | 32.44 (14.81-58.54) | 17.35 (13.16-23.07) |
| Minimum - Maximum | 5.00-246.66 | 5.00-213.06 | 5.00-96.78 |
| Geometric Mean (95% CI) | 8.66 (5.50-13.63) | 29.74 (17.80-49.69) | 17.81 (13.14-24.15) |
| **Day 15 Visit (14 days post-boost)** | | | |
| N (non-missing) | 20 | 20 | 20 |
| Median ($P_{25}$, $P_{75}$) | 541.64 (181.66-683.88) | 611.76 (249.00-1127.14) | 661.71 (198.58-1600.13) |
| Minimum - Maximum | 63.72-1311.06 | 82.36-4665.41 | 81.52-5627.48 |
| Geometric Mean (95% CI) | 407.53 (276.66-600.31) | 572.08 (358.19-913.69) | 582.60 (336.76-1007.90) |
| N* (non-missing pre- and post-boost) | 20 | 20 | 20 |
| Participants with ≥ 2-fold rise[2], 95% CI | 95.0% (75.1%-99.9%) | 100.0% (83.2%-100.0%) | 100.0% (83.2%-100.0%) |
| Participants with ≥ 4-fold rise[2], 95% CI | 95.0% (75.1%-99.9%) | 95.0% (75.1%-99.9%) | 100.0% (83.2%-100.0%) |
| Geometric Mean Fold Rise[2], 95% CI | 47.06 (25.86-85.64) | 19.23 (12.38-29.87) | 32.70 (22.42-47.71) |
| **Day 29 Visit (28 days post-boost)** | | | |
| N (non-missing) | 18 | 20 | 19 |
| Median ($P_{25}$, $P_{75}$) | 331.99 (191.06-484.13) | 569.92 (212.59-860.34) | 374.93 (146.23-906.55) |
| Minimum - Maximum | 38.12-1137.58 | 63.14-2009.86 | 61.96-3908.71 |
| Geometric Mean (95% CI) | 281.66 (188.71-420.39) | 439.79 (283.81-681.50) | 405.60 (233.58-704.32) |
| N* (non-missing pre- and post-boost) | 18 | 20 | 19 |
| Participants with ≥ 2-fold rise[2], 95% CI | 94.4% (72.7%-99.9%) | 100.0% (83.2%-100.0%) | 100.0% (82.4%-100.0%) |
| Participants with ≥ 4-fold rise[2], 95% CI | 94.4% (72.7%-99.9%) | 95.0% (75.1%-99.9%) | 100.0% (82.4%-100.0%) |
| Geometric Mean Fold Rise[2], 95% CI | 32.34 (16.71-62.60) | 14.79 (10.41-20.99) | 23.26 (15.69-34.47) |

[1] Values below the lower limit of detection (LLOD = 10) were assigned the value of LLOD/2

[2] Relative to pre-vaccination (Day 1 Visit) levels, among participants with no-missing observations at both pre- and post-boost timepoints.

Supplemental Table 69: **SARS-CoV-2 Pseudovirion Neutralization Antibody –** mRNA-1273 Neutralization Antibodies Titer (ID50) to Pseudovirus B.1.351 (Beta)[1], by Group, Age Group and Timepoint

| | Group 1E [Dosed Janssen, Boost Moderna] Age 18-55 yo (N=21) | Group 1E [Dosed Janssen, Boost Moderna] Age ≥56 yo (N=32) | Group 2E [Dosed Moderna, Boost Moderna] Age 18-55 yo (N=26) | Group 2E [Dosed Moderna, Boost Moderna] Age ≥56 yo (N=25) | Group 3E [Dosed Pfizer, Boost Moderna] Age 18-55 yo (N=25) | Group 3E [Dosed Pfizer, Boost Moderna] Age ≥56 yo (N=25) |
|---|---|---|---|---|---|---|
| **Day 1 Visit (Pre-boost)** | | | | | | |
| N (non-missing) | 10 | 10 | 10 | 10 | 10 | 10 |
| Median ($P_{25}$, $P_{75}$) | 5.00 (5.00-13.53) | 5.00 (5.00-13.12) | 32.44 (20.84-39.33) | 33.32 (14.79-65.27) | 22.61 (14.72-33.17) | 14.16 (11.81-17.56) |
| Minimum - Maximum | 5.00-22.38 | 5.00-246.66 | 5.00-213.06 | 5.00-202.24 | 13.15-96.78 | 5.00-23.43 |
| Geometric Mean (95% CI) | 7.21 (4.69-11.07) | 10.41 (4.27-25.36) | 29.09 (12.71-66.58) | 30.42 (13.90-66.55) | 24.84 (16.21-38.07) | 12.77 (8.69-18.78) |
| **Day 15 Visit (14 days post-boost)** | | | | | | |
| N (non-missing) | 10 | 10 | 10 | 10 | 10 | 10 |
| Median ($P_{25}$, $P_{75}$) | 544.66 (464.29-632.68) | 364.83 (143.87-801.02) | 611.76 (213.50-970.80) | 671.54 (284.50-1197.28) | 1251.82 (747.00-1682.16) | 230.04 (133.48-512.86) |
| Minimum - Maximum | 63.72-1311.06 | 123.51-1013.32 | 130.77-4665.41 | 82.36-2246.85 | 213.34-5627.48 | 81.52-2018.46 |
| Geometric Mean (95% CI) | 479.11 (276.06-831.54) | 346.64 (183.27-655.65) | 603.13 (286.71-1268.77) | 542.62 (262.92-1119.91) | 1199.67 (636.27-2261.93) | 282.93 (140.86-568.27) |
| N* (non-missing pre- and post-boost) | 10 | 10 | 10 | 10 | 10 | 10 |
| Participants with ≥ 2-fold rise[2], 95% CI | 100.0% (69.2%-100.0%) | 90.0% (55.5%-99.7%) | 100.0% (69.2%-100.0%) | 100.0% (69.2%-100.0%) | 100.0% (69.2%-100.0%) | 100.0% (69.2%-100.0%) |
| Participants with ≥4-fold rise[2], 95% CI | 100.0% (69.2%-100.0%) | 90.0% (55.5%-99.7%) | 90.0% (55.5%-99.7%) | 100.0% (69.2%-100.0%) | 100.0% (69.2%-100.0%) | 100.0% (69.2%-100.0%) |
| Geometric Mean Fold Rise[2], 95% CI | 66.49 (33.92-130.33) | 33.31 (11.25-98.66) | 20.74 (9.20-46.71) | 17.84 (10.41-30.56) | 48.29 (31.18-74.80) | 22.15 (12.37-39.64) |
| **Day 29 Visit (28 days post-boost)** | | | | | | |
| N (non-missing) | 9 | 9 | 10 | 10 | 9 | 10 |
| Median ($P_{25}$, $P_{75}$) | 428.27 (235.32-514.67) | 212.66 (132.05-472.21) | 640.38 (188.83-727.23) | 389.00 (236.02-965.87) | 714.91 (374.93-980.71) | 191.80 (104.49-422.80) |
| Minimum - Maximum | 38.12-651.51 | 108.25-1137.58 | 121.44-1558.39 | 63.14-2009.86 | 267.52-3908.71 | 61.96-2234.84 |
| Geometric Mean (95% CI) | 303.54 (155.34-593.12) | 261.36 (143.66-475.49) | 468.05 (247.97-883.46) | 413.23 (198.41-860.62) | 774.25 (387.41-1547.38) | 226.67 (106.81-481.05) |
| N* (non-missing pre- and post-boost) | 9 | 9 | 10 | 10 | 9 | 10 |
| Participants with ≥ 2-fold rise[2], 95% CI | 100.0% (66.4%-100.0%) | 88.9% (51.8%-99.7%) | 100.0% (69.2%-100.0%) | 100.0% (69.2%-100.0%) | 100.0% (66.4%-100.0%) | 100.0% (69.2%-100.0%) |

| | Group 1E [Dosed Janssen, Boost Moderna] Age 18-55 yo (N=21) | Group 1E [Dosed Janssen, Boost Moderna] Age ≥56 yo (N=32) | Group 2E [Dosed Moderna, Boost Moderna] Age 18-55 yo (N=26) | Group 2E [Dosed Moderna, Boost Moderna] Age ≥56 yo (N=25) | Group 3E [Dosed Pfizer, Boost Moderna] Age 18-55 yo (N=25) | Group 3E [Dosed Pfizer, Boost Moderna] Age ≥56 yo (N=25) |
|---|---|---|---|---|---|---|
| Participants with ≥4-fold rise[2], 95% CI | 100.0% (66.4%-100.0%) | 88.9% (51.8%-99.7%) | 90.0% (55.5%-99.7%) | 100.0% (69.2%-100.0%) | 100.0% (66.4%-100.0%) | 100.0% (69.2%-100.0%) |
| Geometric Mean Fold Rise[2], 95% CI | 45.18 (20.74-98.42) | 23.15 (6.92-77.48) | 16.09 (8.43-30.73) | 13.59 (8.89-20.75) | 31.42 (18.50-53.36) | 17.74 (9.60-32.81) |

[1] Values below the lower limit of detection (LLOD = 10) were assigned the value of LLOD/2

[2] Relative to pre-vaccination (Day 1 Visit) levels, among participants with no-missing observations at both pre- and post-boost timepoints.

Supplemental Table 70: **SARS-CoV-2 Pseudovirion Neutralization Antibody –** mRNA-1273 Neutralization Antibodies Titer (ID80) to Pseudovirus B.1.351 (Beta)[1], by Group and Timepoint

| | **Group 1E [Dosed Janssen, Boost Moderna] (N=53)** | **Group 2E [Dosed Moderna, Boost Moderna] (N=51)** | **Group 3E [Dosed Pfizer, Boost Moderna] (N=50)** |
|---|---|---|---|
| **Day 1 Visit (Pre-boost)** | | | |
| N (non-missing) | 20 | 20 | 20 |
| Median ($P_{25}$, $P_{75}$) | 5.00 (5.00-5.00) | 15.28 (5.00-19.31) | 5.00 (5.00-11.46) |
| Minimum - Maximum | 5.00-53.19 | 5.00-84.98 | 5.00-28.14 |
| Geometric Mean (95% CI) | 5.88 (4.54-7.63) | 12.95 (8.60-19.50) | 7.17 (5.57-9.24) |
| **Day 15 Visit (14 days post-boost)** | | | |
| N (non-missing) | 20 | 20 | 20 |
| Median ($P_{25}$, $P_{75}$) | 254.38 (81.27-321.81) | 293.42 (88.11-434.43) | 265.37 (79.93-474.26) |
| Minimum - Maximum | 23.57-419.13 | 28.01-1374.77 | 38.09-1931.82 |
| Geometric Mean (95% CI) | 175.31 (120.54-254.96) | 217.74 (139.31-340.33) | 213.16 (130.73-347.57) |
| N* (non-missing pre- and post-boost) | 20 | 20 | 20 |
| Participants with ≥ 2-fold rise[2], 95% CI | 95.0% (75.1%-99.9%) | 100.0% (83.2%-100.0%) | 100.0% (83.2%-100.0%) |
| Participants with ≥ 4-fold rise[2], 95% CI | 95.0% (75.1%-99.9%) | 95.0% (75.1%-99.9%) | 100.0% (83.2%-100.0%) |
| Geometric Mean Fold Rise[2], 95% CI | 29.79 (18.20-48.76) | 16.81 (11.53-24.50) | 29.71 (20.65-42.76) |
| **Day 29 Visit (28 days post-boost)** | | | |
| N (non-missing) | 18 | 20 | 19 |
| Median ($P_{25}$, $P_{75}$) | 109.24 (83.55-210.97) | 168.05 (80.44-375.39) | 120.70 (59.47-366.57) |
| Minimum - Maximum | 17.08-413.37 | 20.61-716.85 | 20.37-1544.45 |
| Geometric Mean (95% CI) | 119.08 (80.29-176.59) | 166.83 (108.05-257.59) | 139.63 (81.25-239.97) |
| N* (non-missing pre- and post-boost) | 18 | 20 | 19 |
| Participants with ≥ 2-fold rise[2], 95% CI | 94.4% (72.7%-99.9%) | 100.0% (83.2%-100.0%) | 100.0% (82.4%-100.0%) |
| Participants with ≥ 4-fold rise[2], 95% CI | 88.9% (65.3%-98.6%) | 95.0% (75.1%-99.9%) | 100.0% (82.4%-100.0%) |
| Geometric Mean Fold Rise[2], 95% CI | 19.87 (11.44-34.51) | 12.88 (9.35-17.74) | 19.93 (13.10-30.33) |

[1] Values below the lower limit of detection (LLOD = 10) were assigned the value of LLOD/2

[2] Relative to pre-vaccination (Day 1 Visit) levels, among participants with no-missing observations at both pre- and post-boost timepoints.

Supplemental Table 71: **SARS-CoV-2 Pseudovirion Neutralization Antibody –** mRNA-1273 Neutralization Antibodies Titer (ID80) to Pseudovirus B.1.351 (Beta)[1], by Group, Age Group and Timepoint

| | Group 1E [Dosed Janssen, Boost Moderna] Age 18-55 yo (N=21) | Group 1E [Dosed Janssen, Boost Moderna] Age ≥56 yo (N=32) | Group 2E [Dosed Moderna, Boost Moderna] Age 18-55 yo (N=26) | Group 2E [Dosed Moderna, Boost Moderna] Age ≥56 yo (N=25) | Group 3E [Dosed Pfizer, Boost Moderna] Age 18-55 yo (N=25) | Group 3E [Dosed Pfizer, Boost Moderna] Age ≥56 yo (N=25) |
|---|---|---|---|---|---|---|
| **Day 1 Visit (Pre-boost)** | | | | | | |
| N (non-missing) | 10 | 10 | 10 | 10 | 10 | 10 |
| Median ($P_{25}$, $P_{75}$) | 5.00 (5.00-5.00) | 5.00 (5.00-5.00) | 15.28 (12.32-16.27) | 11.01 (5.00-19.90) | 11.46 (5.00-13.61) | 5.00 (5.00-5.00) |
| Minimum - Maximum | 5.00-12.22 | 5.00-53.19 | 5.00-84.00 | 5.00-84.98 | 5.00-28.14 | 5.00-5.00 |
| Geometric Mean (95% CI) | 5.47 (4.47-6.69) | 6.33 (3.71-10.81) | 14.73 (8.38-25.91) | 11.39 (5.67-22.89) | 10.29 (6.83-15.51) | 5.00 (5.00-5.00) |
| **Day 15 Visit (14 days post-boost)** | | | | | | |
| N (non-missing) | 10 | 10 | 10 | 10 | 10 | 10 |
| Median ($P_{25}$, $P_{75}$) | 272.36 (133.08-302.90) | 164.02 (73.70-340.71) | 293.42 (88.40-358.15) | 305.92 (87.82-450.72) | 426.77 (279.31-520.91) | 82.73 (65.97-250.27) |
| Minimum - Maximum | 23.57-419.13 | 66.12-370.54 | 64.82-1374.77 | 28.01-614.16 | 86.79-1931.82 | 38.09-629.28 |
| Geometric Mean (95% CI) | 198.94 (108.61-364.42) | 154.48 (88.73-268.94) | 239.02 (123.22-463.67) | 198.36 (95.47-412.14) | 403.46 (229.56-709.08) | 112.62 (59.92-211.65) |
| N* (non-missing pre- and post-boost) | 10 | 10 | 10 | 10 | 10 | 10 |
| Participants with ≥ 2-fold rise[2], 95% CI | 100.0% (69.2%-100.0%) | 90.0% (55.5%-99.7%) | 100.0% (69.2%-100.0%) | 100.0% (69.2%-100.0%) | 100.0% (69.2%-100.0%) | 100.0% (69.2%-100.0%) |
| Participants with ≥ 4-fold rise[2], 95% CI | 100.0% (69.2%-100.0%) | 90.0% (55.5%-99.7%) | 90.0% (55.5%-99.7%) | 100.0% (69.2%-100.0%) | 100.0% (69.2%-100.0%) | 100.0% (69.2%-100.0%) |
| Geometric Mean Fold Rise[2], 95% CI | 36.39 (19.59-67.59) | 24.39 (10.15-58.61) | 16.23 (8.70-30.26) | 17.42 (9.98-30.40) | 39.20 (26.02-59.05) | 22.52 (11.98-42.33) |
| **Day 29 Visit (28 days post-boost)** | | | | | | |
| N (non-missing) | 9 | 9 | 10 | 10 | 9 | 10 |
| Median ($P_{25}$, $P_{75}$) | 129.27 (92.91-210.97) | 88.84 (72.51-207.21) | 199.05 (79.26-327.25) | 138.68 (81.63-395.94) | 347.36 (120.70-409.81) | 72.61 (52.47-121.89) |
| Minimum - Maximum | 17.08-315.44 | 44.94-413.37 | 50.97-485.52 | 20.61-716.85 | 90.86-1544.45 | 20.37-536.24 |
| Geometric Mean (95% CI) | 123.01 (62.91-240.49) | 115.27 (64.47-206.09) | 177.81 (100.20-315.55) | 156.53 (72.28-338.96) | 268.29 (128.64-559.56) | 77.57 (39.33-152.99) |
| N* (non-missing pre- and post-boost) | 9 | 9 | 10 | 10 | 9 | 10 |
| Participants with ≥ 2-fold rise[2], 95% CI | 100.0% (66.4%-100.0%) | 88.9% (51.8%-99.7%) | 100.0% (69.2%-100.0%) | 100.0% (69.2%-100.0%) | 100.0% (66.4%-100.0%) | 100.0% (69.2%-100.0%) |
| Participants with ≥ 4-fold rise[2], 95% CI | 88.9% (51.8%-99.7%) | 88.9% (51.8%-99.7%) | 90.0% (55.5%-99.7%) | 100.0% (69.2%-100.0%) | 100.0% (66.4%-100.0%) | 100.0% (69.2%-100.0%) |
| Geometric Mean Fold Rise[2], 95% CI | 22.27 (11.02-45.03) | 17.73 (6.41-49.00) | 12.07 (7.25-20.09) | 13.75 (8.42-22.45) | 26.32 (15.06-46.00) | 15.51 (7.87-30.60) |

[1] Values below the lower limit of detection (LLOD = 10) were assigned the value of LLOD/2

[2] Relative to pre-vaccination (Day 1 Visit) levels, among participants with no-missing observations at both pre- and post-boost timepoints

#### Supplemental Figure 1: Consort Diagram for Stage 1 (Groups 1-3)

**166 Assessed for Eligibility**
Eligible but not enrolled (N=0)
Incomplete Screening (N=1)
Ineligible (N=11)
Ineligible and enrolled (N=0)

**154 Enrolled to Receive mRNA-1273 Booster Vaccination**

**Ad26-COV2-S EUA Dosed- 5 x $10^{10}$ vp (N = 53)**

**18-55 years (N = 21)**
- Completed Day 8 Safety Assessment (N = 21)
- Completed Day 15 Visit (N = 21)

**56 years and older (N = 32)**
- Completed Day 8 Safety Assessment (N = 32)
- Completed Day 15 Visit (N = 32)

**Endpoints Analyzed for 53 Vaccine Recipients:**
- **53** Analyzed for vaccine safety
- **52** Analyzed for Binding Antibodies
- **52** Analyzed for PsVNA Assay (D614G)
- **20** Analyzed for PsVNA Assay (B.1.617.2 and B.1.351)

**mRNA-1273 EUA Dosed - 100-mcg (N = 51)**

**18-55 years (N = 26)**
- Completed Day 8 Safety Assessment (N = 26)
- Completed Day 15 Visit (N = 26)

**56 years and older (N = 25)**
- Completed Day 8 Safety Assessment (N = 25)
- Completed Day 15 Visit (N = 25)

**Endpoints Analyzed for 51 Vaccine Recipients:**
- **51** Analyzed for vaccine safety
- **50** Analyzed for Binding Antibodies
- **50** Analyzed for PsVNA Assay (D614G)
- **20** Analyzed for PsVNA Assay (B.1.617.2 and B.1.351)

**BNT162b2 EUA Dosed - 30-mcg (N = 50)**

**18-55 years (N = 25)**
- Completed Day 8 Safety Assessment (N = 25)
- Completed Day 15 Visit (N = 25)

**56 years and older (N = 25)**
- Completed Day 8 Safety Assessment (N = 25)
- Completed Day 15 Visit (N = 25)

**Endpoints Analyzed for 50 Vaccine Recipients:**
- **50** Analyzed for vaccine safety
- **50** Analyzed for Binding Antibodies
- **50** Analyzed for PsVNA Assay (D614G)
- **20** Analyzed for PsVNA Assay (B.1.617.2 and B.1.351)

**Supplementary Figure 1. Stage 1 Enrollment, Booster Vaccinations and Study Endpoints Analyses**

#### Supplemental Figure 2: Consort Diagram for Stage 2 (Groups 4-6)

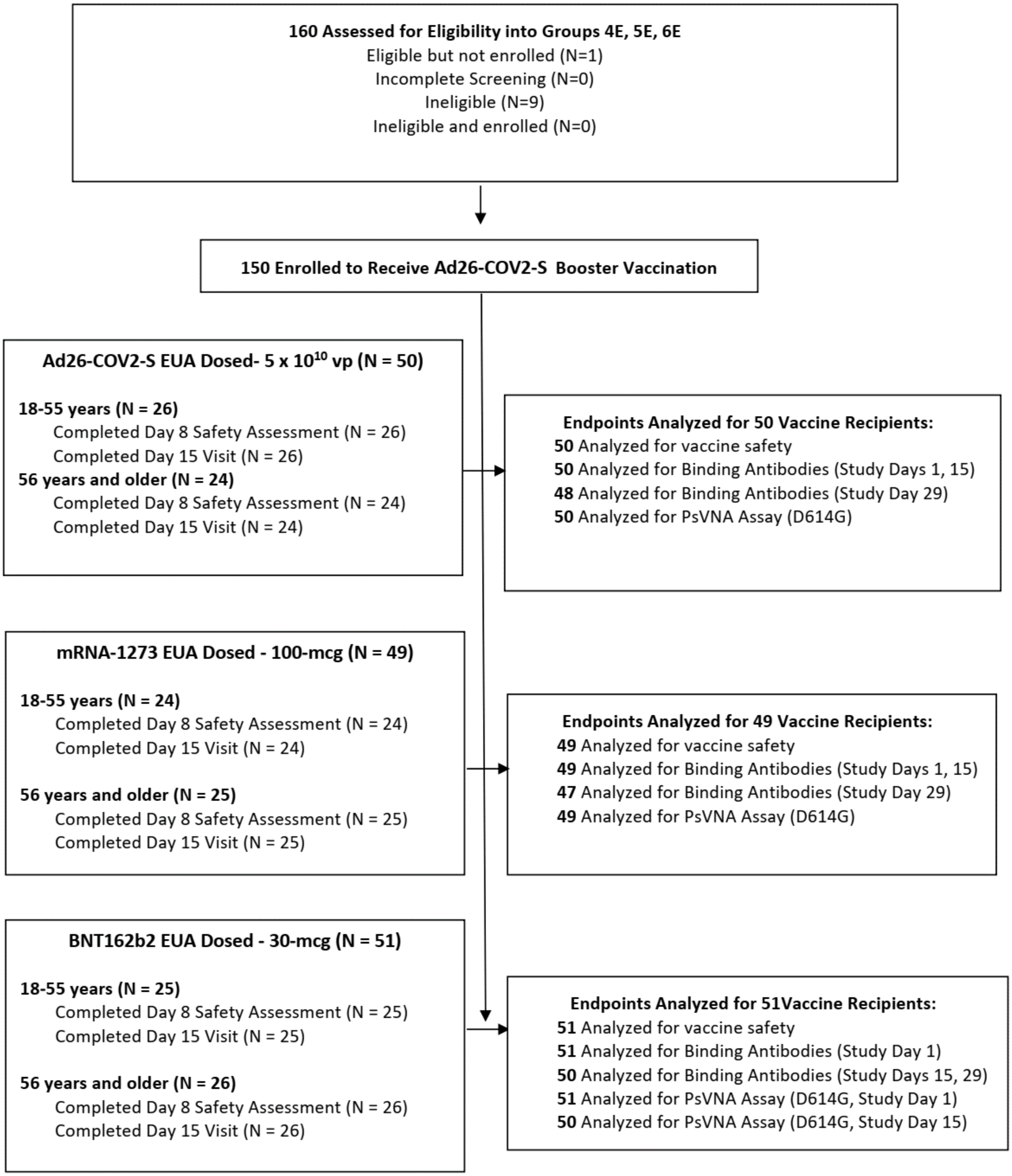

**Supplementary Figure 2.** **Stage 2 Enrollment, Booster Vaccinations and Study Endpoints Analyses**

#### Supplemental Figure 3: Consort Diagram for Stage 3 (Groups 7-9)

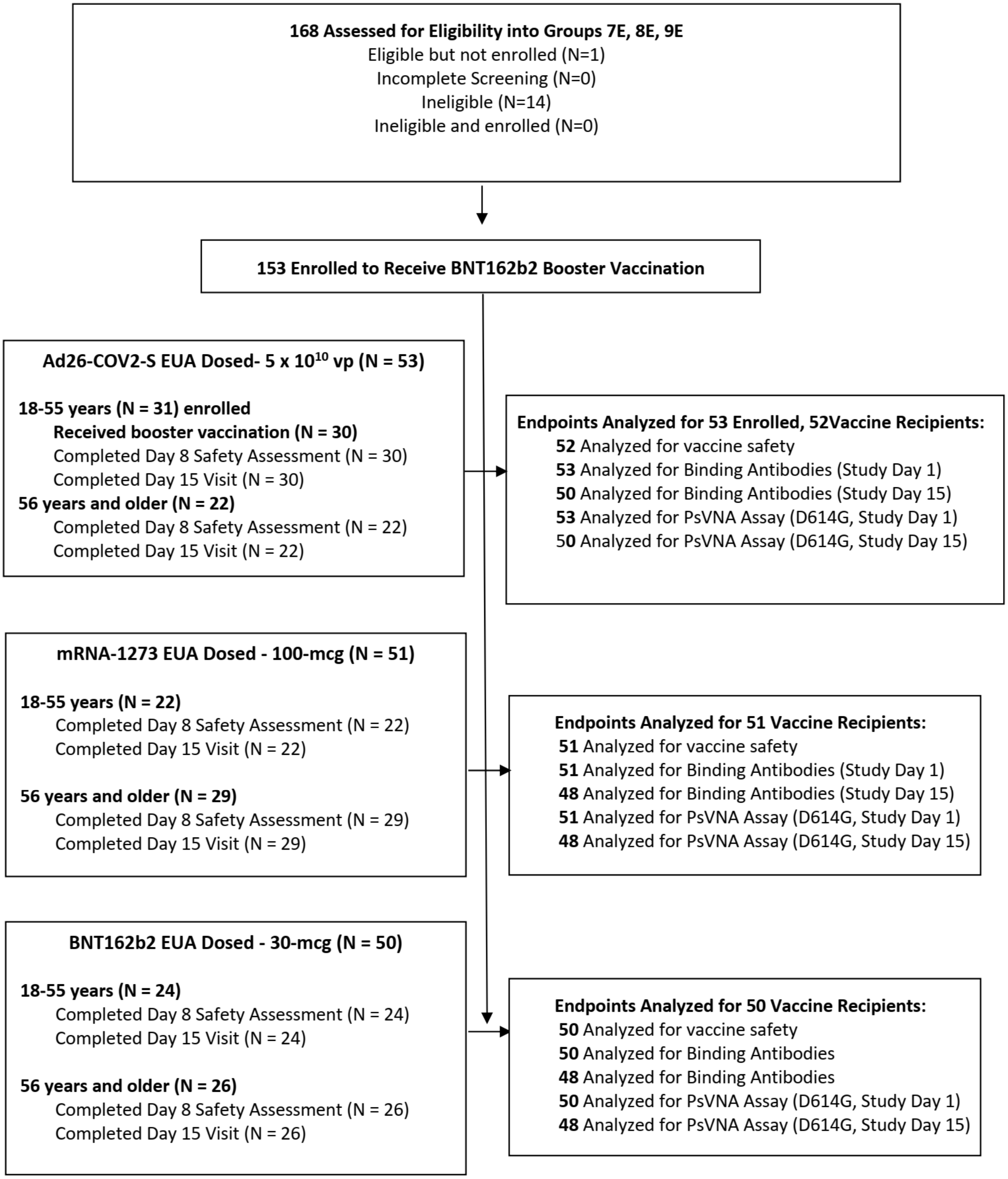

**Supplementary Figure 3.** Stage 3 Enrollment, Booster Vaccinations and Study Endpoints Analyses
